# Spatial transcriptomics reveals mechanism of autoimmunity driven by internalized autoantibodies

**DOI:** 10.64898/2026.02.14.26346329

**Authors:** Iago Pinal-Fernandez, Katherine Pak, Maria Casal-Dominguez, Sandra Muñoz-Braceras, Gustaf Wigerblad, Stefania Dell’Orso, Faiza Naz, Shamima Islam, Gustavo Gutierrez-Cruz, Travis B. Kinder, S. Amara Ogbonnaya-Whittlesey, Andreu Fernández-Codina, Margherita Giannini, Benjamin Ellezam, Gilles Laverny, Valentine Gilbart, Océane Landon-Cardinal, Marie Hudson, Yves Troyanov, Davide Randazzo, Aster Kenea, Ana Matas-Garcia, Gloria Garrabou, Iban Aldecoa, Gabriela Ailen-Caballero, Albert Gil-Vila, Ernesto Trallero, Margherita Milone, Teerin Liewluck, Elie Naddaf, Gerard Espinosa, Carmen Pilar Simeon-Aznar, Alfredo Guillen-Del-Castillo, Corinna Preusse, Felix Kleefeld, Nikolas Bublitz, Werner Stenzel, Alain Meyer, Janet E. Pope, Albert Selva-O’Callaghan, Jose C. Milisenda, Andrew L. Mammen

## Abstract

Autoantibody internalization has been implicated in autoimmune disease pathogenesis, yet its mechanisms, and generality across different diseases, cell types, and affected tissues remain poorly defined. Using bulk RNA sequencing, we identified reproducible, autoantibody-specific transcriptomic signatures consistent with autoantigen dysfunction in muscle biopsies from patients with anti-Mi2 dermatomyositis and anti-PM/Scl scleromyositis across independent cohorts. Electroporation of purified patient IgG into primary cultures of healthy cells was sufficient to induce the corresponding transcriptomic programs *in vitro*. Direct immunofluorescence demonstrated immunoglobulin internalization into subcellular compartments matching the localization of the autoantigen in different affected tissues. Spatial transcriptomic analyses revealed that antibody-secreting cells translocated cytoplasmic material (i.e., immunoglobulin RNA) into adjacent affected cells expressing autoantibody-specific transcripts. The disease-specific transcripts were present not only in muscle fibers, but also in other cells, including macrophages, endothelial cells, and fibroblasts. Autoantibody-induced transcriptomic programs were associated with cell damage and autoantibody-specific reactive inflammatory programs, including activation of type I interferon and TGF-ß1 signaling in anti-Mi2 dermatomyositis and activation of type II interferon in anti-PM/Scl scleromyositis. Antibody internalization was also observed in different tissues from patients with other autoimmune diseases, including anti-U1RNP mixed connective tissue disease, anti-Ku overlap syndrome, and anti-Scl70 systemic sclerosis. Together, these findings establish autoantibody internalization as a shared pathogenic mechanism across diverse autoimmune diseases, providing a unifying framework for conditions driven by autoantibodies against intracellular antigens.

## INTRODUCTION

Autoimmunity has been mechanistically linked to a limited set of well-defined processes, including germline mutations driving autoinflammatory syndromes^1^, somatic mutations such as those causing VEXAS^2^, and autoantibodies directed against cell-surface proteins that perturb receptor or channel function, as in myasthenia gravis or Graves’ disease^3^. Despite major advances in these areas, many prevalent autoimmune diseases in rheumatology (e.g., myositis, systemic sclerosis, vasculitis, Sjögren, systemic lupus erythematosus), endocrinology (e.g., type 1 diabetes, Hashimoto thyroiditis), neurology (e.g. selected autoimmune encephalopathies and neuropathies), gastroenterology (e.g. primary biliary cholangitis, autoimmune hepatitis), ophthalmology (e.g. autoimmune retinitis), and other subspecialties are not explained by these mechanisms^3^. Instead, they are unified by the presence of autoantibodies against intracellular antigens. These conditions are typically described as the consequence of complex interactions between environmental, polygenic, and acquired factors. In the absence of a defined pathogenic mechanism, classification criteria have organized patients into clinically useful but heterogeneous entities (such as myositis or systemic sclerosis).

A parsimonious mechanistic hypothesis for the pathogenesis of these diseases with autoantibodies against intracellular autoantigens is that the autoantibodies themselves are directly pathogenic^3^. Autoantibody internalization into living cells has been proposed for decades^4^, but this concept was long overshadowed by the dogma that immunoglobulins cannot access intracellular compartments *in vivo*, the limited functional effects of immunoglobulin incubation *in vitro* in the absence of forced internalization^5^, and a lack of robust methods to detect and study intracellular antibodies in relevant human cells and tissues. As a result, disruption of autoantigen function by internalized autoantibodies was largely considered an unlikely mechanism. However, recent work has revived this idea by developing *in vitro* approaches to probe the intracellular actions of autoantibodies and by demonstrating autoantibody internalization with secondary autoantigen dysfunction in myositis and scleromyositis *in vivo*^5,6^. Several groups have provided convergent evidence that, in selected settings, internalization of disease-specific autoantibodies is necessary and sufficient to induce tissue-specific transcriptional changes^7,8^. However, key issues remain unresolved: how autoantibodies gain access to intracellular compartments *in vivo*, whether this mechanism is restricted to specific diseases or tissues or is more pervasive across systemic autoimmunity, which cell types are affected, and how antibody internalization leads to distinct inflammatory programs.

Among the conditions in which this mechanism has been most rigorously interrogated, anti-Mi2 dermatomyositis and anti-PM/Scl scleromyositis are particularly informative. Both are characterized by autoantibodies recognizing intracellular targets in well-defined subcellular locations—Mi2-associated antigens in the nucleus and components of the nuclear RNA exosome complex in the nucleolus—and by distinctive gene expression patterns associated to autoantigen dysfunction. In anti-Mi2 dermatomyositis, autoantibodies initially thought to recognize Mi2a and Mi2b of the Nucleosome Remodeling Deacetylase (NuRD) complex^9^, a transcriptomic repressor, were subsequently shown to target the PHD domain of multiple proteins^10,11^, and to be associated with a specific transcriptional program involving derepression of normally repressed gene sets and ectopic expression of tissue-restricted transcripts in muscle^5,6^. In anti-PM/Scl scleromyositis, autoantibodies target the nuclear RNA exosome components responsible for degrading long noncoding RNAs, including divergent and antisense transcripts^12^. Muscle tissue from anti-PM/Scl scleromyositis patients accumulates these exosome substrates, directly linking autoantibody specificity to autoantigen dysfunction^5^. Notably, internalization of purified patient immunoglobulin into healthy cells *in vitro* recapitulates these transcriptomic changes, supporting a causal role for intracellular antibodies. These two diseases thus offer model systems in which the relationships among autoantibody specificity, subcellular localization, and transcriptional consequences can be dissected.

To address the outstanding questions regarding autoantibody internalization as a mechanism of disease, we assembled multicenter cohorts of patients with anti-Mi2 and anti-PM/Scl autoantibodies, together with additional autoantibody-defined systemic autoimmune diseases, and integrated bulk RNA sequencing, direct immunofluorescence, functional assays with purified IgG, and spatial transcriptomics. First, we asked whether autoantibody-specific muscle transcriptomic signatures in anti-Mi2 dermatomyositis and anti-PM/Scl scleromyositis are robust and reproducible in independent patient cohorts. Second, we validated whether purified patient IgG is sufficient to induce these signatures in primary human skeletal muscle cells *in vitro*. Third, we mapped the subcellular distribution of internalized immunoglobulin in muscle and skin, focusing on nuclear and nucleolar compartments in these index diseases. Fourth, using spatial transcriptomics, we sought to identify how autoantibodies gain intracellular access *in vivo*, to define the cell types that exhibit autoantibody-specific transcriptional programs, and to characterize the associated inflammatory pathways. Finally, we asked whether similar patterns of autoantibody internalization occur in other autoimmune diseases characterized by autoantibodies to intracellular antigens, including anti-U1RNP mixed connective tissue disease, anti-Ku overlap syndrome, and anti-Scl70 systemic sclerosis, and whether these patterns extend beyond muscle to other affected tissues.

## RESULTS

To address the outstanding questions regarding autoantibody internalization, we assembled a multicenter, cross-sectional cohort comprising 814 muscle biopsies from individuals with autoantibody-defined myopathies, healthy comparators, genetic myopathies, and individuals with other autoimmune diseases (e.g. systemic sclerosis, systemic lupus erythematosus) (Supplementary Fig.1). Serological groups included those with anti-Mi2, anti-PM/Scl, anti-NXP2, anti-TIF1γ, anti-MDA5, anti-Jo1, anti-HMGCR, anti-SRP, anti-U1RNP, anti-Ku, anti-Scl70, and anti-centromere autoantibodies.

An external validation cohort, processed independently outside the NIH and drawn from distinct geographical regions (Canada and France), was used to validate anti-PM/Scl-associated transcriptomic signatures (Supplementary Fig.1). This cohort included 41 muscle biopsy specimens from patients with anti-PM/Scl autoantibodies, along with healthy and disease comparators.

From the primary cohort, we selected subsets of muscle biopsies—and, when available, paired skin biopsies—for complementary experimental approaches. Serum IgG purified from representative patients was electroporated into primary human skeletal muscle cells to test whether autoantibodies are sufficient to induce previously defined disease-specific transcriptional programs^5^. In parallel, a subset of biopsies underwent spatial transcriptomic profiling using a custom capture panel targeting: (i) disease-associated transcripts previously linked to anti-Mi2 and anti-PM/Scl^5^, (ii) immunoglobulin genes, (iii) muscle and stromal cell markers, and (iv) type I and type II interferon-inducible genes.

### Robust autoantibody-specific muscle transcriptomic signatures in anti-Mi2 and anti-PM/Scl myopathies

Building on prior work, we first asked whether the previously identified autoantibody-specific transcriptional programs associated with anti-Mi2 dermatomyositis (more than 100 genes) and anti-PM/Scl scleromyositis (more than 200 genes)^5^ are robust and generalizable. Bulk RNA sequencing of the expanded cohort of diagnostic muscle biopsies confirmed these two highly distinctive gene programs (Fig.1, Supplementary Fig.2, Supplementary Tbl.1).

**Figure 1.**
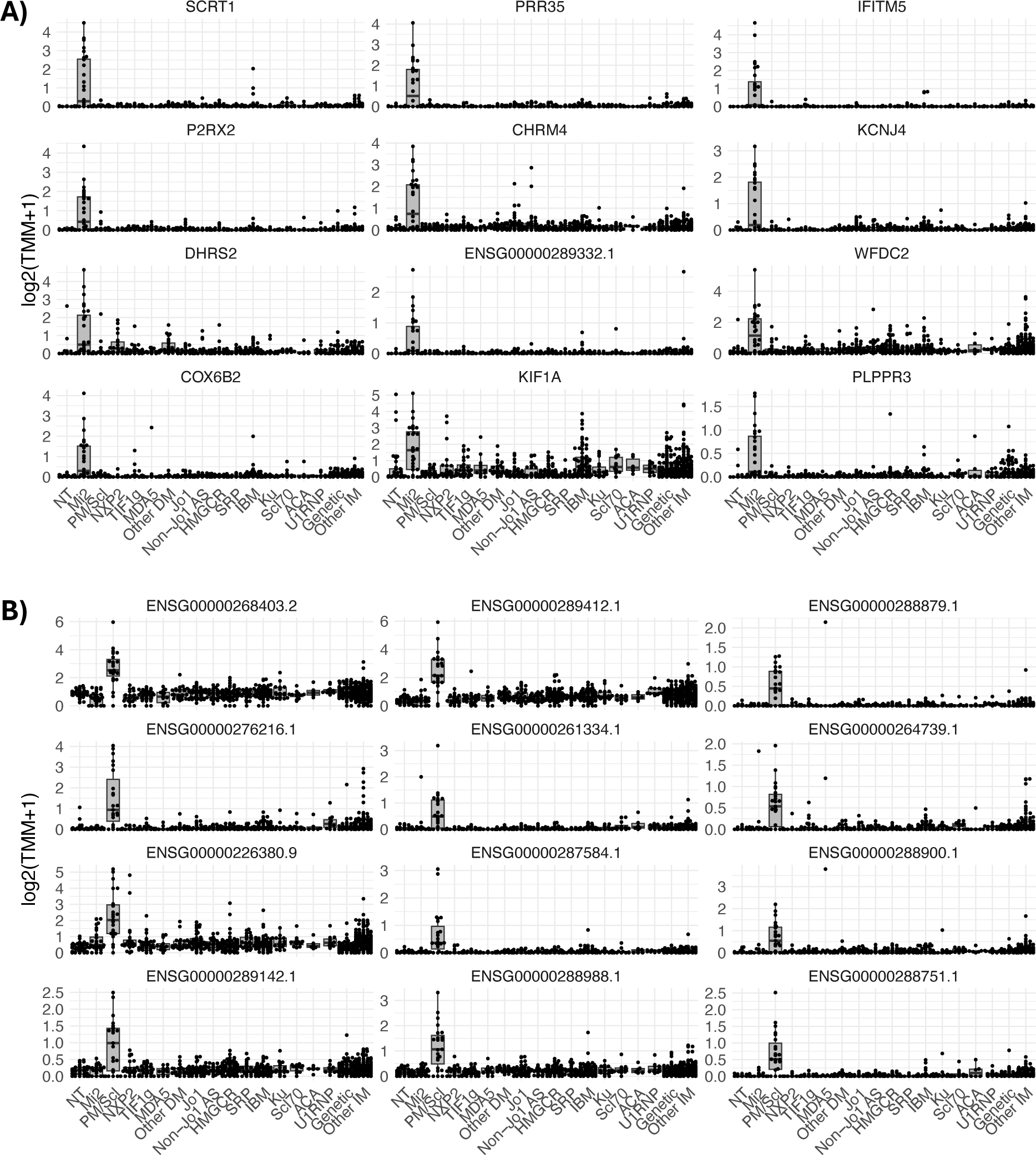
Expression of the anti-Mi2 (A) and anti-PM/Scl (B) specific gene sets in different types of myositis. Specifically overexpressed genes in muscle biopsies from anti-Mi2 (A) and anti-PM/Scl (B) patients compared to all the other groups included in the study. Each dot represents the gene expression value from a single patient. NT: histologically normal muscle biopsies; DM: dermatomyositis; AS: antisynthetase syndrome; IBM: inclusion body myositis; ACA, anti-centromere autoantibodies; IM: inflammatory myopathies.

Muscle biopsies from patients with anti-Mi2 autoantibodies exhibited a consistent pattern of transcriptional derepression, marked by upregulation of gene sets previously defined as Mi2/NuRD-regulated targets and by ectopic expression of normally repressed, tissue-restricted transcripts^5^. This anti-Mi2-specific gene set was strongly enriched in anti-Mi2 biopsies and largely absent from other serological subgroups and disease controls, including samples obtained from multiple independent centers (Fig.1A, Supplementary Fig.2A, Supplementary Tbl.1).

In contrast, muscle from patients with anti-PM/Scl autoantibodies displayed a distinct transcriptomic signature characterized by the accumulation of long noncoding RNAs, including divergent and antisense transcripts, corresponding to known substrates of the nuclear RNA exosome (Fig.1B, Supplementary Fig.2B, Supplementary Tbl.2)^5^. This PM/Scl-specific gene set was highly specific to patients with anti-PM/Scl autoantibodies and was not observed in other inflammatory myopathies or comparator conditions. Importantly, this signature was reproducible across centers, including an external validation cohort comprising biopsies from France and Canada that were processed independently at the University of Strasbourg (Supplementary Fig.3, Supplementary Tbl.3), supporting the robustness of the findings and arguing against site-specific or technical artifacts.

### Patient IgG is sufficient to induce autoantibody-specific transcriptional programs *in vitro*

To determine whether the observed autoantibody-specific transcriptional programs are directly driven by patient immunoglobulin, we electroporated purified IgG from patients with anti-Mi2 and anti-PM/Scl autoantibodies into primary human skeletal muscle cells, expanding upon prior experiments to include additional patient samples and control groups^5^. Transcriptomic profiling at serial time points following IgG internalization demonstrated that intracellular patient IgG was sufficient to recapitulate the corresponding *in vivo* muscle transcriptional signatures (Fig.2; Supplementary Fig.4-7, Supplementary Tbl.4-6).

**Figure 2.**
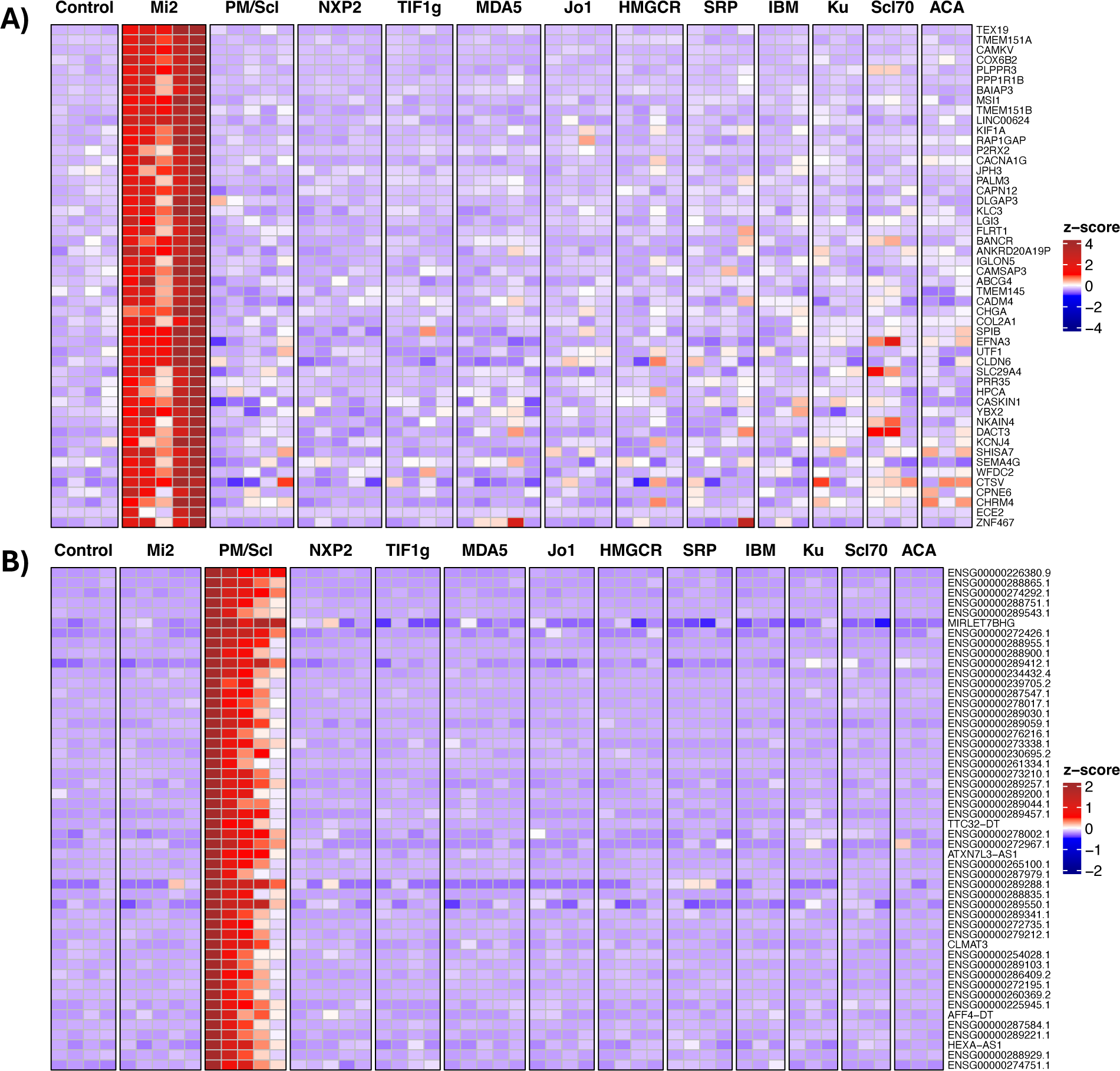
Anti-Mi2- and anti-PM/Scl-specific gene expression in cultured human skeletal muscle cells following IgG electroporation. Heatmaps display standardized gene expression levels (Z-scores) of the top 50 specific genes associated with anti-Mi2 (A) and anti-PM/Scl (B) responses, measured at 24h (A) and 72h (B) after electroporation with purified IgG from myositis patients or healthy controls. Each column represents IgG from an individual donor. Robust and selective induction of anti-Mi2-specific (A) and anti-PM/Scl-specific (B) gene signatures is observed exclusively in cells electroporated with IgG from anti-Mi2-positive and anti-PM/Scl-positive patients, respectively. IBM, inclusion body myositis; ACA, anti-centromere autoantibodies.

Cells electroporated with anti-Mi2 IgG rapidly induced the Mi2-associated transcriptional derepression program, with high expression of Mi2-specific genes—including *SCRT1*, *CAMKV*, and other targets— one day after internalization (Fig.2; Supplementary Fig.4, Supplementary Tbl.4-5). In contrast, cells electroporated with anti-PM/Scl IgG showed progressive accumulation of long noncoding and divergent transcripts over time, with stronger signal of the PM/Scl-specific program occurring three days after internalization (Fig.2; Supplementary Fig.5, Supplementary Tbl.4 and 6).

These distinct temporal dynamics mirror the known biology of the targeted intracellular complexes—rapid chromatin-associated effects for Mi-2-related targets versus slower accumulation of nuclear RNA exosome substrates for PM/Scl—and support mechanistically divergent modes of transcriptional disruption mediated by each autoantibody.

### IgG is internalized into distinct nuclear and nucleolar compartments in muscle and skin

We next asked whether patient immunoglobulin localizes to the subcellular compartments occupied by its cognate autoantigens (Fig.3). Building on our prior observations in muscle^5^, confocal direct immunofluorescence (DIF) of fresh frozen muscle sections from anti-Mi2 patients confirmed robust IgG deposition within myonuclei without discrete nucleolar enrichment (Fig.3A-C). In anti-PM/Scl muscle, IgG accumulated predominantly within the nucleoli of myonuclei, consistent with the nuclear localization of anti-PM/Scl autoantigens (Fig.3G-I).

**Figure 3.**
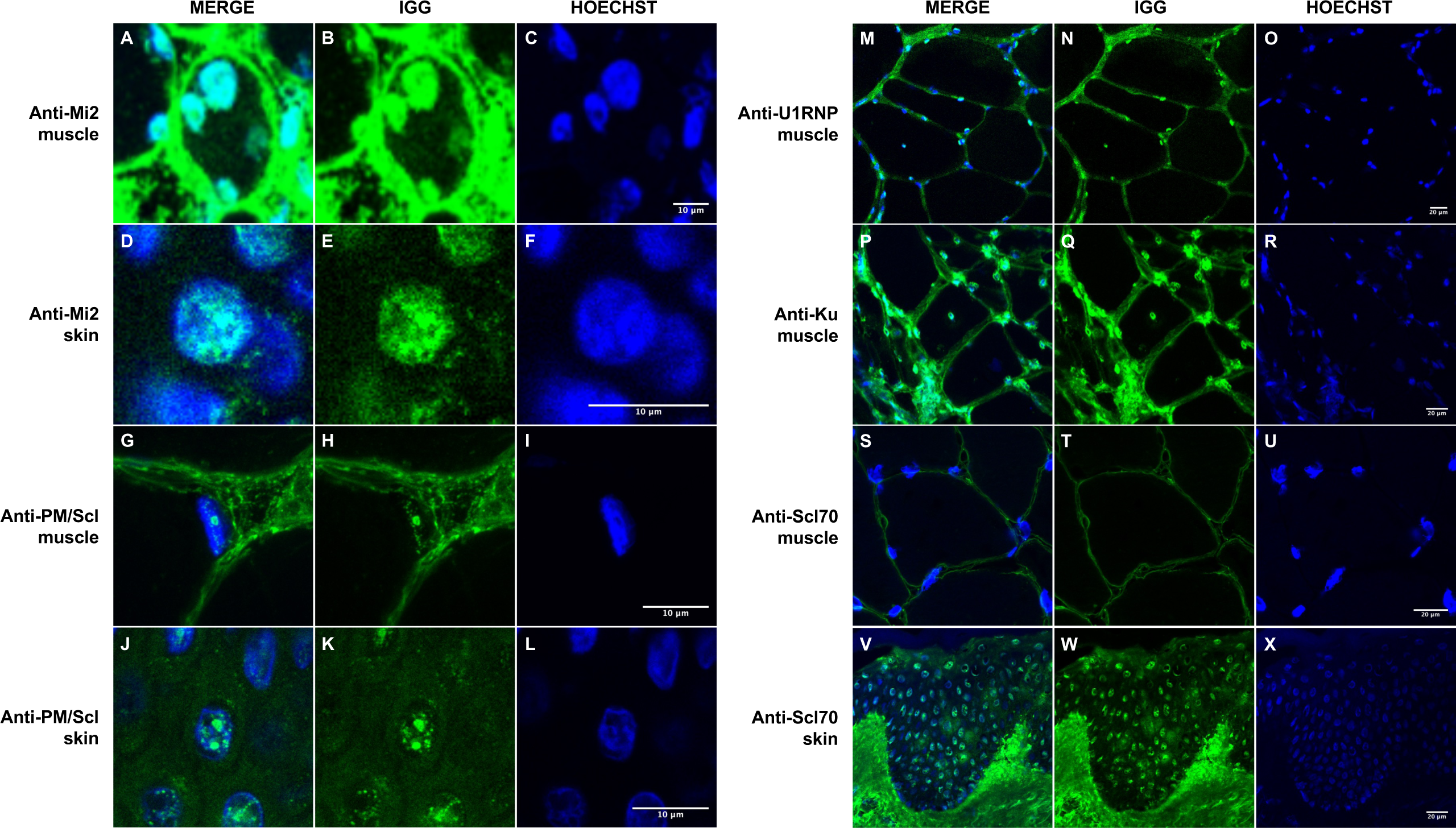
Immunoglobulin localization in muscle and skin from patients with anti-Mi2, anti-PM/Scl, anti-U1RNP, anti-Ku, and anti-Scl70 autoantibodies. Confocal immunofluorescence analysis of human IgG in muscle and skin tissues demonstrates autoantibody-specific patterns of nuclear localization. In patients with anti-Mi2 autoantibodies, IgG deposition is observed within the nuclei of muscle (A-C) and skin cells (D-F). Patients with anti-PM/Scl autoantibodies exhibit predominant nucleolar accumulation of IgG in both muscle (G-I) and skin cells (J-L). In contrast, anti-U1RNP (M-O) and anti-Ku (P-R) autoantibodies are associated with nuclear IgG accumulation in muscle tissue, whereas patients with anti-Scl70 (S-U) autoantibodies show no nuclear IgG accumulation in muscle but display clear nuclear localization in skin tissue (V-X).

We next extended these analyses to skin. Anti-Mi2 skin biopsies showed epidermal IgG accumulation within keratinocyte nuclei without nucleolar enrichment (Fig.3D-F). In contrast, in anti-PM/Scl skin biopsies, keratinocyte nuclei displayed a nucleolar IgG staining pattern that closely mirrored the distribution observed in muscle (Fig.3J-L). Collectively, these findings demonstrate that internalized IgG can access nuclear and nucleolar compartments across various tissues, and that its subnuclear localization faithfully reflects the known intracellular distribution of the targeted autoantigens.

### Internalized autoantibodies induce cell damage, reprograms multiple cell types, and activate autoantibody-specific inflammatory pathways

To define how autoantibody internalization reshapes the tissue microenvironment, we designed a custom spatial transcriptomics capture panel targeting: (i) anti-Mi2-specific transcripts (e.g. *SCRT1*, *CAMKV*)^5^, (ii) anti-PM/Scl-associated long noncoding and divergent transcripts^5^, (iii) immunoglobulin genes, (iv) muscle and stromal cell markers, and (v) genes induced by type I and type II interferon signaling (Supplementary Video 1).

#### Anti-Mi2 muscle: focal gene derepression, type I interferon gradients, cell damage and TGFβ overexpression

In anti-Mi2 dermatomyositis, spatial transcriptomic maps revealed marked heterogeneity in the expression of Mi2-specific transcripts across individual myofibers. A discrete subset of fibers, predominantly perifascicular, exhibited high levels of Mi2-specific genes in both the cytoplasm and nuclei, whereas adjacent fibers showed little or no induction (Fig.4A; Supp Fig.8A, 9A, and 10, Supp Vid.1). This patchwork pattern indicates that autoantibody internalization—and its downstream transcriptional effects—occurs focally rather than uniformly across the tissue, closely paralleling the characteristic distribution of muscle fiber involvement in anti-Mi2 dermatomyositis^13^.

**Figure 4.**
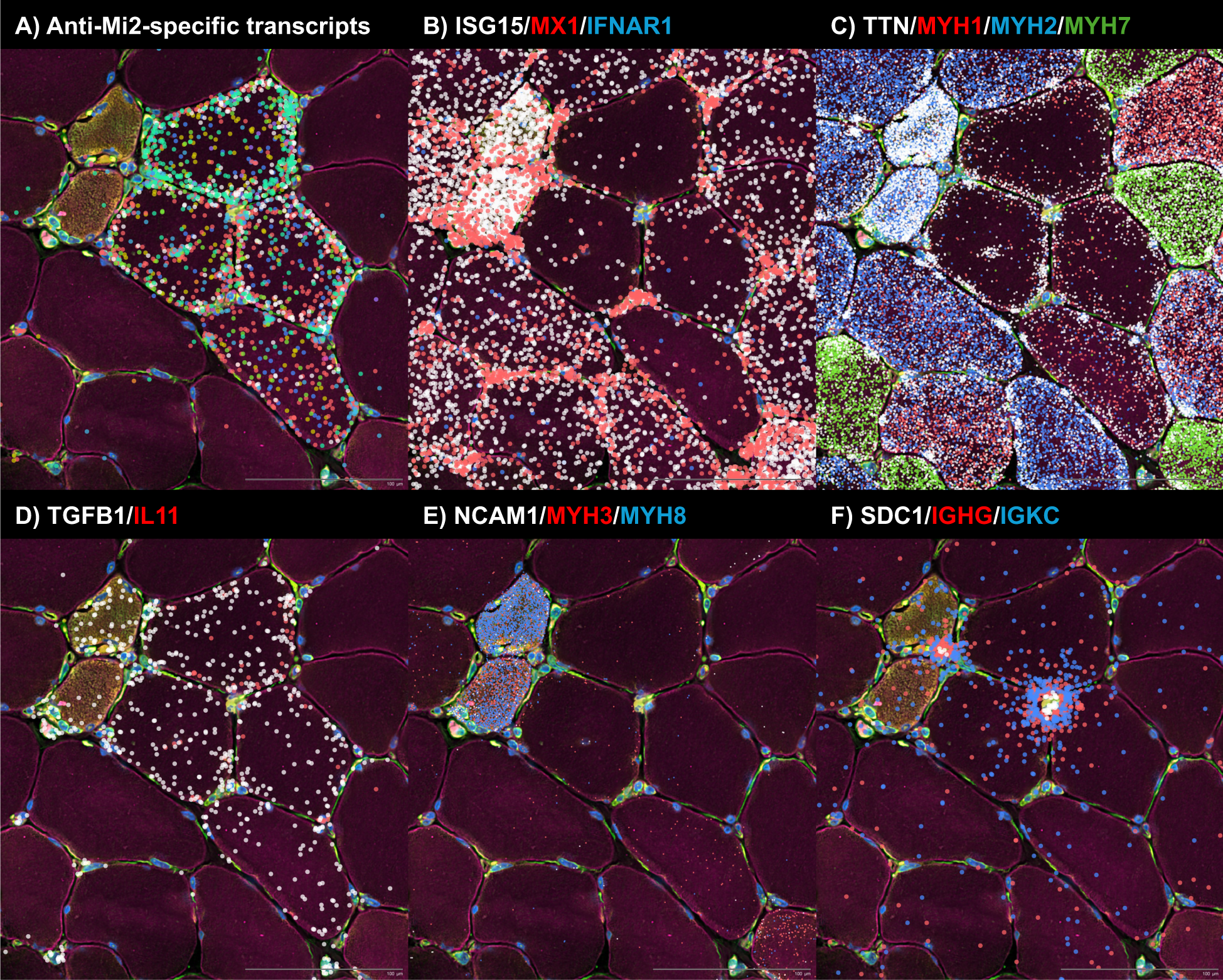
Spatial transcriptomic analysis of a representative biopsy area from an anti-Mi2-positive patient, highlighting molecular signatures consistent with autoantibody internalization, autoantibody internalization-associated tissue damage and inflammation, and IgG-producing antibody-secreting cells exhibiting immunoglobulin RNA externalization. Colored dots indicate the spatial distribution of: (A) anti-Mi2-specific transcripts; (B) type I interferon-inducible genes (*ISG15*, *MX1*) and the interferon receptor *IFNAR1*; (C) transcripts associated with mature muscle fibers (*TTN*) and specific muscle fiber types (*MYH1*, type IIx; *MYH2*, type IIa; *MYH7*, type I); (D) *TGFB1* and *IL11*; (E) markers of muscle regeneration (*NCAM1*, *MYH3*, *MYH8*); and (F) antibody-secreting cell markers (*SDC1*), immunoglobulin G heavy-chain constant region (*IGHG*), and immunoglobulin kappa constant region (*IGKC*).

A paradox in anti-Mi2 dermatomyositis has been the discrepancy between the robust type I interferon (IFN-I) signature detected by bulk RNAseq in patient muscle biopsies and the relatively modest induction of IFN-I-responsive genes following internalization of purified IgG from anti-Mi2 patients *in vitro*. Spatial transcriptomics resolves this discordance. Cells immediately adjacent to those expressing high levels of Mi2-specific transcripts—including fibroblasts and regenerating myoblasts—exhibited strong expression of IFN-I-inducible genes such as *ISG15* and *MX1*. In contrast, the fibers expressing high levels of Mi2-specific transcripts themselves showed minimal IFN-I gene induction, associated with reduced expression of the type I IFN receptor *IFNAR1* (Fig.4B; Supplementary Fig.8B and 9B). These findings support a model in which fibers that internalize anti-Mi2 autoantibodies become relatively refractory to IFN-I signaling due to downregulation of the IFN receptor, while surrounding cells mount a pronounced interferon response.

Another unresolved question has been whether autoantibody internalization—and the transcriptional programs it induces—is directly linked to damage of target cells. Spatial transcriptomic analysis revealed that Mi2-high fibers exhibited reduced expression of canonical markers of mature adult muscle, including *MYH1* (type 2X fibers), *MYH2* (type 2A fibers), *MYH7* (type 1 fibers), and *TTN*. The coordinated loss of these key muscle transcripts suggests that activation of the Mi2-specific gene set is intrinsically deleterious to the fibers in which it occurs (Fig.4C; Supplementary Fig.8C and 9C). Importantly, numerous fibers display clear induction of Mi2-specific transcripts while retaining expression of key muscle genes, indicating that the anti-Mi2 transcriptional signature precedes the development of muscle toxicity rather than arising as a consequence of it (Supplementary Fig.9C).

In addition to regenerating muscle fibers (Fig.4E; Supplementary Fig.8E and 9E), Mi2-high fibers showed selective upregulation of *TGFB1* (Fig.4D; Supplementary Fig.8D and 9D), a cytokine whose sustained expression in muscle is known to promote atrophy, fibrosis, and impaired regeneration.^14^ Consistent with this, *IL11*—a downstream effector of TGFβ signaling and one of the Mi2-specific transcripts previously identified—was co-induced within these fibers^15^.

Together, these data support a sequential model in which intracellular access of anti-Mi2 autoantibodies triggers focal gene derepression within affected myofibers, drives *TGFB1* overexpression and muscle injury, and elicits a secondary interferon response in surrounding cells.

#### Anti-PM/Scl muscle: nuclear accumulation of nuclear RNA exosome substrates and IFNγ-biased inflammation

In muscle from anti-PM/Scl patients, PM/Scl-specific long noncoding and divergent transcripts accumulated in a highly focal, predominantly intranuclear pattern that was distinct from the distributions observed in other myopathies profiled in parallel (Fig.5A and 5E, Supplementary Figures 10 and 11A).

**Figure 5.**
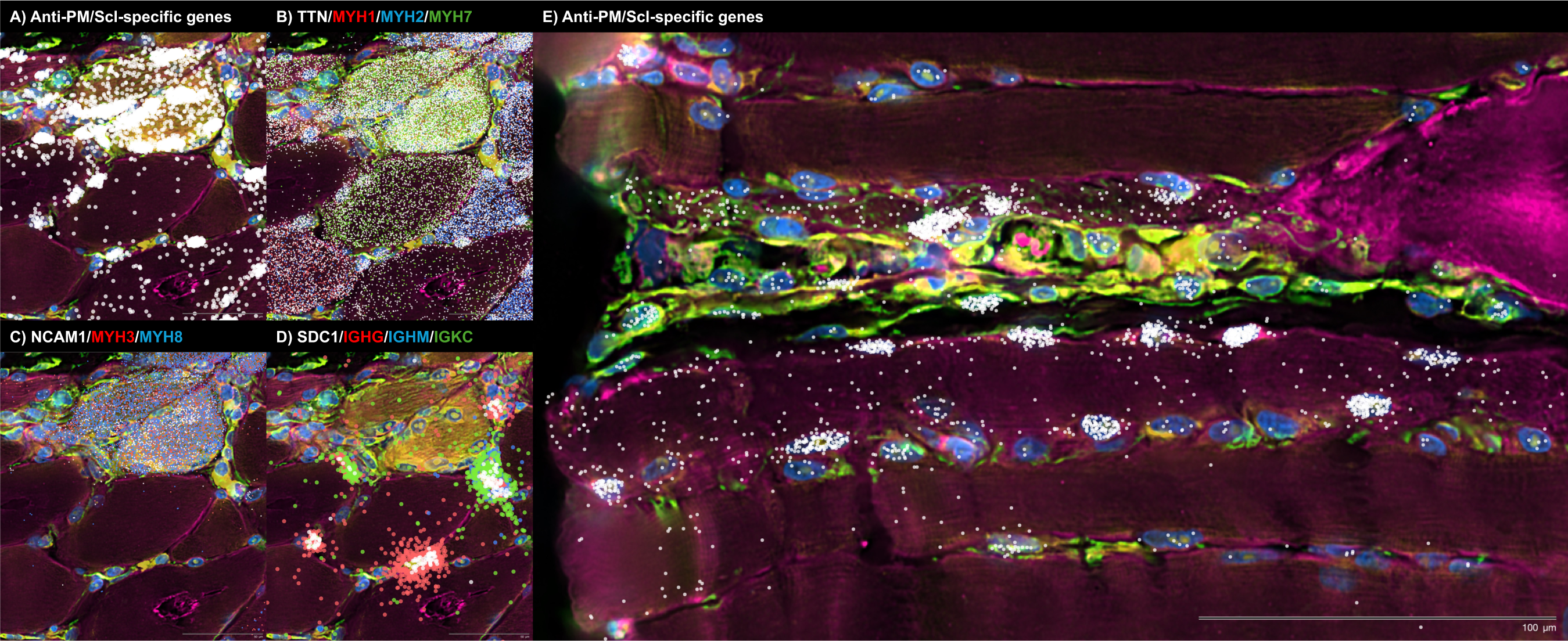
Spatial transcriptomic analysis of two representative biopsy regions from an anti-PM/Scl-positive patient, highlighting molecular signatures consistent with autoantibody internalization, autoantibody internalization-associated tissue damage and immunoglobulin RNA externalization by IgG-producing antibody-secreting cells. Colored dots depict the spatial distribution of: (A, E) anti-PM/Scl-specific transcripts; (B) transcripts associated with mature muscle fibers (*TTN*) and specific fiber types (*MYH1*, type IIx; *MYH2*, type IIa; *MYH7*, type I); (C) markers of muscle regeneration (*NCAM1*, *MYH3*, *MYH8*); and (D) antibody-secreting cell markers (*SDC1*), immunoglobulin G heavy-chain constant region (*IGHG*), immunoglobulin M heavy-chain constant region (*IGHM*), and immunoglobulin kappa constant region (*IGKC*).

As in anti-Mi2^+^ dermatomyositis, a direct link between autoantibody internalization and muscle fiber damage has not yet been established. Using spatial transcriptomics, we found that fibers enriched for PM/Scl-specific transcripts showed reduced expression of canonical markers of mature adult muscle, including *MYH1* (type 2X fibers), *MYH2* (type 2A fibers), *MYH7* (type 1 fibers), and *TTN* (Fig.4B, Supplementary Fig.11B). This coordinated downregulation of key structural and contractile genes suggests that the transcriptional program induced by intracellular anti-PM/Scl autoantibodies is intrinsically deleterious to the affected muscle fibers.

Importantly, and consistent with observations in anti-Mi2 dermatomyositis, many fibers exhibited robust induction of PM/Scl-specific transcripts while maintaining normal expression of essential muscle genes. This suggests that the anti-PM/Scl transcriptional signature precedes muscle toxicity rather than arising as a secondary consequence of fiber damage (Fig.4B, Supplementary Fig.11B). Notably, a subset of transcript-rich fibers also expressed markers of regeneration, including *NCAM1*, *MYH3*, and *MYH8*, consistent with activation of an attempted repair response (Fig.4C).

Spatial hotspots of PM/Scl-specific transcripts coincided with dense inflammatory infiltrates composed of fibroblasts, macrophages, T cells, and pre-class-switched IgM-expressing B cells (Supplementary Fig.11D and E), suggesting that accumulation of these nuclear RNA exosome substrates marks sites of active immune engagement.

Both spatial and bulk transcriptomic analyses converged on a type II interferon (IFNγ)-biased inflammatory milieu in anti-PM/Scl muscle. IFNγ-inducible genes were strongly enriched in immune cells surrounding PM/Scl transcript-rich fibers (Supplementary Fig.11C), and muscle biopsies from anti-PM/Scl patients exhibited higher IFNγ-inducible gene expression than anti-Scl70 and anti-centromere comparators (Supplementary Fig.12). Together, these findings link the intranuclear accumulation of nuclear RNA exosome substrates in muscle fibers to the formation of IFNγ-polarized, B-cell-rich^16^ inflammatory microenvironments characteristic of this condition.

### Autoantibody-induced transcriptomic signatures are also observed in macrophages, fibroblasts, and endothelial cells

In earlier work, we observed internalization of autoantibodies into cells outside of myofibers; however, we were unable to confidently assign these signals to specific cell types and therefore did not pursue this observation further^5^. With spatial transcriptomics, we could now resolve the cellular heterogeneity of this process and compare its distribution among autoantibody specificities. In anti-Mi2 muscle, Mi2-specific transcripts were detected in cells of the macrophage lineage, which notably represented only a small subset of cells with detectable *IFNB1* expression (Supplementary Fig.13). By contrast, in anti-PM/Scl biopsies, PM/Scl-specific transcripts were detected not only in myonuclei but also in the nuclei of endothelial cells and fibroblasts (Supplementary Fig.13).

### Spatial transcriptomics implicates autoantibody secreting cell-to-tissue cell transfer of immunoglobulin as a route of intracellular antibody access

A central unresolved question is how immunoglobulins against intracellular antigens gain access to the cytoplasm and nucleus of tissue cells in vivo. Spatial transcriptomics provided a key clue. In both anti-Mi2 and anti-PM/Scl muscle, we identified antibody-secreting cells adjacent to fibers expressing high levels of the corresponding disease-specific gene programs (Fig.4F, Fig.5D, Supplementary Fig.8F, 9F, and 11F). We detected a spatial gradient of immunoglobulin transcripts radiating from antibody-secreting cell regions into neighboring cells.

Notably, externalization of immunoglobulin RNA was predominantly observed in antibody-secreting cells, with minimal externalization detected in B cells (Fig.6). Importantly, this process exhibited marked sequence specificity. *SDC1* (CD138), *CD38*, and *IGHGP* transcripts were largely retained within antibody-secreting cells, with little signal detected outside the cell membrane. In contrast, Ig heavy- and light-chain transcripts were depleted within antibody-secreting cells and instead enriched in adjacent spatial spots, including within surrounding muscle cells and in nearby intercellular spaces. Collectively, these findings support a unidirectional and specialized transfer of Ig RNA-containing cytoplasmic material from antibody-secreting cells to nearby cells.

**Figure 6.**
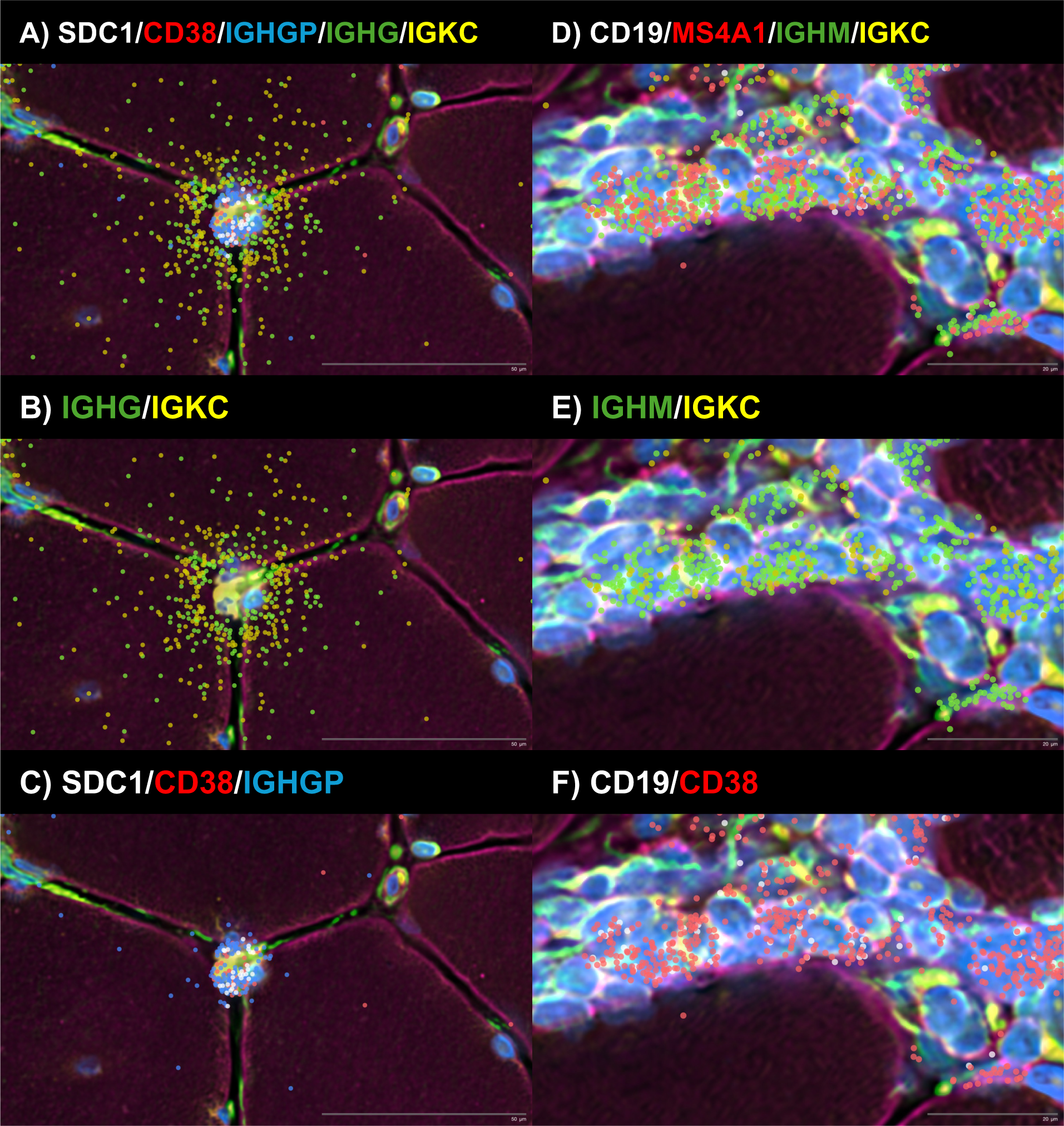
Spatial transcriptomic analysis of representative antibody-secreting cells (A-C) and B cells (D-F) reveals preferential externalization of immunoglobulin RNA, including both heavy-and light-chain transcripts, in antibody-secreting cells, with minimal externalization observed in B cells. In contrast, transcripts encoding canonical autoantibody secreting cell surface markers (*SDC1* and *CD38*) and the immunoglobulin pseudogene *IGHGP* were preferentially not externalized in antibody-secreting cells. These findings indicate that immunoglobulin RNA externalization is a specialized, antibody-secreting cell-acquired process that exhibits sequence specificity.

Technical features of the assay argue against diffusion or misassignment of immunoglobulin transcripts as an artifactual explanation. At the spatial resolution used, transcripts from individual cells, including small cells, remain sharply confined within their morphologically defined boundaries rather than “bleeding” into adjacent cells; this is illustrated in Supplementary Fig. 14, where transcripts corresponding to distinct muscle cell types are contained within their respective cellular boundaries.

### Autoantibody internalization aligns with autoantigen localization across systemic autoimmune diseases and tissues

Finally, we asked whether the patterns observed in anti-Mi2 and anti-PM/Scl muscle biopsies extend to other autoimmune conditions associated with autoantibodies recognizing intracellular targets. We previously demonstrated autoantibody internalization in muscle biopsies from patients with anti-NXP2, anti-TIF1γ, and anti-MDA5 dermatomyositis, anti-Jo1 antisynthetase syndrome, as well as anti-HMGCR and anti-SRP immune-mediated necrotizing myopathy^5^. Here, we expanded our direct immunofluorescence analyses to include muscle biopsies from patients with anti-U1RNP mixed connective tissue disease and anti-Ku overlap syndrome, as well as muscle and skin biopsies from patients with anti-Scl70 systemic sclerosis.

In anti-U1RNP and anti-Ku muscle, confocal DIF revealed nuclear IgG accumulation with subnuclear distributions consistent with the known intracellular localization of the respective ribonucleoprotein targets (Fig.3M-R). In contrast, anti-Scl70 biopsies showed prominent epidermal IgG deposition in skin cells (Fig.3V-X), whereas muscle biopsies exhibited little or no nuclear IgG internalization (Fig.3S-U), consistent with the predominant vascular and cutaneous tropism of anti-Scl70 disease. Across all autoantibody groups examined, IgG deposition in at least one affected tissue aligned with the expected subcellular distribution of the cognate autoantigen.

These findings were independently validated in muscle biopsies from anti-U1RNP- and anti-Ku-positive patients, in which direct immunofluorescence was performed in Canada (Supplementary Fig.15).

## DISCUSSION

Autoantibodies directed against intracellular antigens define many common systemic and tissue-specific autoimmune diseases. Yet their pathogenic role has long been difficult to reconcile with a basic principle of cell biology: immunoglobulins are not expected to cross intact plasma membranes. This conceptual disconnect has favored models in which such autoantibodies are viewed largely as epiphenomena, with tissue injury attributed instead to indirect immune mechanisms. Here, by integrating large-cohort bulk transcriptomics with functional studies of IgG internalization, direct tissue immunofluorescence, and spatial transcriptomics, we provide convergent evidence for a distinct and unifying framework. Our data support a model in which antibody-secreting cells enable intracellular delivery of autoantibodies to compartments containing their cognate targets *in vivo*, where antibody binding directly disrupts autoantigen function. These intracellular effects are sufficient to drive focal tissue-cell injury and to couple target dysfunction to stereotyped, yet distinct, inflammatory microenvironments (Extended Fig. 1).

A central observation is the robustness and specificity of tissue transcriptional programs linked to individual autoantibody specificities. In a multicenter cohort of diagnostic muscle biopsies spanning autoantibody-defined inflammatory myopathies and related systemic autoimmune diseases, anti-Mi2 dermatomyositis and anti-PM/Scl scleromyositis each exhibit highly distinctive transcriptomic signatures that are largely absent from other serological groups and disease controls. Importantly, the anti-PM/Scl-specific gene set replicates in an independently processed external cohort from different geographic regions, arguing against site-specific artifacts and underscoring that these signatures reflect conserved biology rather than technical idiosyncrasy. In this context, the autoantibody is not merely a clinical stratifier but maps onto reproducible molecular pathology within the affected tissue.

To test the sufficiency of internalized autoantibodies to drive disease-specific gene signatures, we delivered purified patient IgG into primary human skeletal muscle cells and asked whether intracellular immunoglobulin could reproduce the *in vivo* programs. IgG from anti-Mi2 dermatomyositis rapidly induces the Mi2-specific gene derepression, whereas IgG from anti-PM/Scl scleromyositis drives a slower, progressive accumulation of long non-coding RNAs consistent with impaired nuclear RNA exosome function. The divergent kinetics are mechanistically coherent with the distinct functions of the targeted complexes—rapid anti-Mi2-induced transcriptional derepression versus gradual accumulation of RNA substrates following anti-PM/Scl-induced exosome dysfunction—and reinforce the interpretation that these signatures represent functional footprints of autoantigen disruption rather than secondary inflammatory effects.

We next asked whether immunoglobulin is present within tissue cells *in vivo* and, if so, whether its subcellular distribution aligns with the location of the targeted antigens. In anti-PM/Scl scleromyositis, IgG accumulates within nucleoli of myofibers and keratinocytes, consistent with the nucleolar localization of nuclear RNA exosome components. In anti-Mi2 dermatomyositis, IgG localizes diffusely to nuclei without nucleolar enrichment, reflecting the distribution of anti-Mi2 autoantigens. Similar patterns of intracellular IgG localization were observed in other autoantibody-defined conditions, including anti-U1RNP mixed connective tissue disease and anti-Ku overlap syndrome, as well as epidermal IgG deposition in anti-Scl70 systemic sclerosis skin, consistent with disease-specific tissue tropism. Together, these findings indicate that intracellular autoantibody internalization is not restricted to a single disease or tissue and that IgG localization tracks with autoantigen distribution, coinciding with prior evidence from other forms of myositis (e.g., anti-MDA5, anti-Jo1, anti-HMGCR)^5,7,17^ and from additional autoimmune diseases^18–20^.

Spatial transcriptomics then resolves how these intracellular events are organized in the tissue and how they relate to injury and inflammation. In anti-Mi2 muscle, expression of the Mi2-specific gene set is strikingly heterogeneous: a subset of myofibers, predominantly perifascicular, shows high induction while adjacent fibers show little or none, generating a patchwork pattern that parallels the focal myofiber injury characteristic of anti-Mi2 dermatomyositis^13^. Fibers expressing the Mi2-specific program also show coordinated loss of canonical adult muscle transcripts, suggesting this transcriptional state is intrinsically deleterious. A similar relationship is observed in anti-PM/Scl scleromyositis: muscle cells expressing the anti-PM/Scl-specific genet set have reduced expression of structural and contractile muscle genes. In both diseases, some fibers display the disease-specific signatures in the absence of overt loss of adult muscle transcripts, suggesting that activation of these transcriptional programs precedes, rather than merely reflects, tissue damage.

The spatial data also provide insight into how autoantibody-specific tissue-cell perturbations could become coupled to distinct inflammatory programs. In anti-Mi2 disease, type I interferon-inducible transcripts are most prominent in cells surrounding fibers with a high anti-Mi2-specific transcriptional signature rather than within those fibers themselves, which exhibit reduced expression of the type I interferon receptor. This pattern is consistent with a paracrine interferon response in the local microenvironment and relative refractoriness of the affected fibers. Notably, a subset of macrophages expressing anti-Mi2-associated transcripts also shows detectable *IFNB1* expression, suggesting that intracellular exposure to these autoantibodies in this cell type may directly induce interferon-β production. In contrast, in anti-PM/Scl scleromyositis, fibers with a high anti-PM/Scl-specific signature are embedded within immune infiltrates enriched for fibroblasts, macrophages, T cells, and B-lineage cells and are associated with a type II interferon (IFNγ)-biased program. Thus, although these diseases converge on a shared proximal event—intracellular autoantibody-associated target dysfunction—they diverge in both the dominant interferon axis and the cellular architecture of the accompanying inflammatory response.

Importantly, the spatial transcriptomic maps provide a clue about how autoantibodies could gain intracellular access *in vivo*. In both anti-Mi2 and anti-PM/Scl muscle, antibody-secreting cells are positioned adjacent to cells expressing disease-specific transcriptional programs, and we detect gradients of immunoglobulin transcripts extending from antibody-secreting cells into neighboring muscle cells. Notably, these cells externalize immunoglobulin RNA but not transcripts encoding other canonical plasma-cell markers, such as *SDC1* (*CD138*) or *CD38*, suggesting a specialized, sequence-selective export mechanism. B-lineage cells have previously been shown to export immunoglobulin-containing cytoplasmic material via extracellular vesicles^21^, a process that could account for the observed sequence specificity. Although this mode of transfer will require direct mechanistic validation, it provides a plausible route for local intracellular delivery and offers a natural explanation for the focality of tissue-cell reprogramming: intracellular access would be most efficient near antibody-secreting cell niches, generating spatially restricted patterns of target dysfunction, injury, and secondary inflammation.

Spatial transcriptomics also reveals differences in the breadth of cell types directly affected by intracellular autoantibodies. In anti-Mi2 dermatomyositis, the disease-specific signature is detected in a subset of macrophages that express high levels of *IFNB1*. In contrast, in anti-PM/Scl scleromyositis, nuclear accumulation of RNA exosome substrates (i.e. the anti-PM/Scl specific gene set) is observed not only in myofibers but also in endothelial cells and fibroblasts, suggesting that autoantibody internalization directly perturbs stromal compartments implicated in vasculopathy and fibrosis. These differences illustrate how a shared pathogenic principle—intracellular autoantibody activity—can give rise to divergent organ- and cell-type-specific phenotypes across systemic autoimmune diseases.

Several limitations define key next steps. First, although IgG delivery using nucleofection in cultured cells establishes sufficiency, it bypasses physiological uptake; defining endogenous entry routes—including contributions from membrane injury, receptor-mediated uptake, vesicular transfer, or other mechanisms—will be essential. Second, while spatial evidence for transfer from antibody-secreting cells to tissue cells is suggestive, direct proof of translation in recipient cells or transfer of immunoglobulin protein or fragments into surrounding parenchyma is lacking. Orthogonal approaches that track labelled autoantibodies, autoantibody fragments, or autoantibody-encoding material *in vivo*, combined with perturbation of candidate pathways, will be required. Third, for additional intracellular specificities (for example, anti-U1RNP or anti-Ku autoantibodies), predicted cellular consequences such as splicing disruption or impaired DNA repair remain to be demonstrated directly in tissue.

In aggregate, our results argue that pathogenic internalization of autoantibodies is a shared driver of pathology across diverse autoimmune diseases defined by autoantibodies recognizing intracellular antigens. By linking autoantibody specificity to subcellular IgG localization, to distinct and reproducible signatures of target dysfunction, and to spatially organized patterns of injury and inflammation, this work provides a unifying mechanistic framework for conditions that have historically been grouped by phenotype rather than by cause.

**Extended Figure 1.**
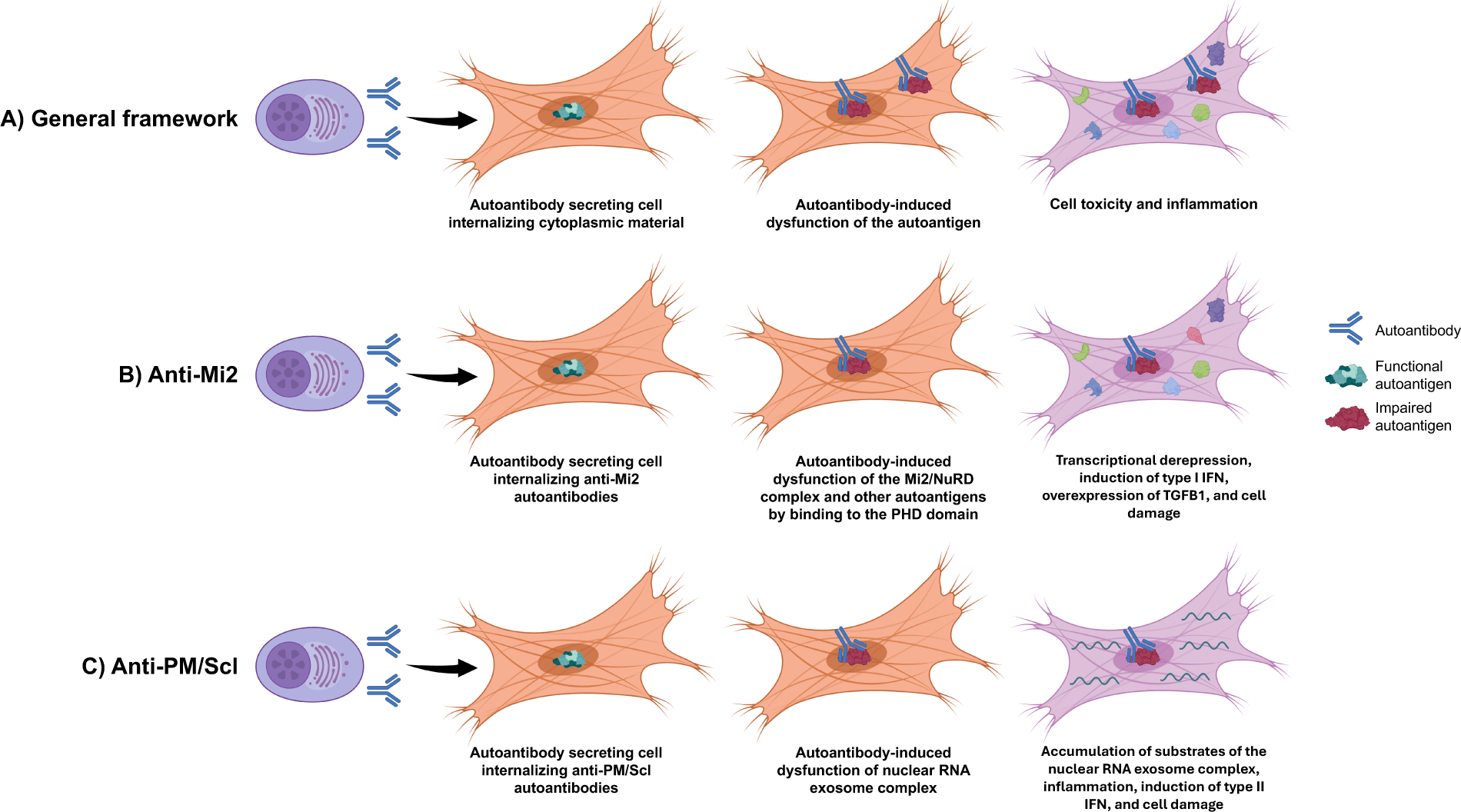
Framework for pathogenic autoantibody internalization in autoimmunity. Autoantibodies secreted by autoreactive B cells can be internalized by neighboring cells, where they interfere with the normal function of their cognate autoantigens, resulting in cellular toxicity and inflammation (A). In anti-Mi2 dermatomyositis, autoantibodies bind the PHD domains of multiple nuclear proteins, leading to transcriptional derepression of target genes, induction of type I interferon signaling in surrounding—but not autoantibody-containing—cells, accompanied by reduced type I interferon receptor expression, and increased *TGFB1* expression, ultimately promoting cellular damage (B). In anti-PM/Scl overlap syndrome, autoantibodies target components of the nuclear RNA exosome complex, impairing its function and causing accumulation of undegraded RNA substrates. This RNA dysregulation drives inflammation, induces type II interferon responses, and results in tissue injury in affected cells (C).

## METHODS

### Study design and participants

We performed a multicenter, cross-sectional analysis of muscle biopsies and, when available, paired skin biopsies from participants enrolled in institutional review board-approved natural history protocols at the National Institutes of Health, Johns Hopkins, Hospital Clinic (Barcelona), Vall d’Hebron Hospital (Barcelona), Mayo Clinic, Strasbourg University Hospitals, Centre hospitalier de l’Université de Montréal (CHUM), and Charité-Universitätsmedizin Berlin. Inclusion required a clinically indicated muscle or skin biopsy and serologic positivity for one of the following autoantibodies: anti-Mi2, anti-NXP2, anti-TIF1γ, anti-MDA5, anti-Jo1, anti-HMGCR, anti-SRP, anti-PM/Scl, anti-U1RNP, anti-Ku, anti-Scl70 (topoisomerase I), or anti-centromere. Autoantibodies were ascertained by the originating institutions using clinically validated methods (e.g., immunoprecipitation, line blot, ELISA, or immunodiffusion) according to laboratory standards. Biopsies from individuals with other genetic or autoimmune diseases were included as comparator samples.

### Standard protocol approvals and patient consent

This study was approved by the Institutional Review Boards of the National Institutes of Health; Johns Hopkins University; Hospital Clinic of Barcelona; Vall d’Hebron University Hospital; Mayo Clinic; Strasbourg University Hospitals; Centre hospitalier de l’Université de Montréal (CHUM); Charité–Universitätsmedizin Berlin, and Western University. Written informed consent was obtained from each participant. All methods were performed in accordance with the relevant guidelines and regulations.

### Tissue processing

Muscle biopsies were processed according to standard clinical protocols. For immunofluorescence, 10 µm cryosections were prepared from frozen tissue. Skin biopsies (when present) were processed analogously from lesional or clinically involved sites, or from clinically normal skin when lesional skin was unavailable (details in Supplementary Methods).

### Direct immunofluorescence (DIF) for human IgG

DIF without fixation (fresh-frozen cryosections) for muscle and skin were prepared as follows: sections were blocked, incubated for either one hour or overnight at room temperature with fluorochrome-conjugated goat anti-human IgG (H+L, Invitrogen A11013), washed, counterstained with Hoechst 33258, and mounted. Images were acquired on a Leica SP8 confocal microscope.

DIF with fixation: 10 µm frozen sections were fixed in 95% ethanol, incubated 30 minutes at room temperature with FITC-conjugated rabbit anti-human IgG (Dako/Agilent F0202, 1:20 dilution), and cover-slipped in Vectashield antifade mounting medium with DAPI to stain nuclei. Images were acquired on an Axio Imager.M1 microscope with an Axiocam 820 color camera (Zeiss).

For skin, direct immunofluorescence (DIF) was performed on unfixed frozen sections. Cryosections (10 µm) were blocked and then incubated with fluorochrome-conjugated goat anti-human IgG (H+L) (Invitrogen A11013) for 1 h or overnight at room temperature, followed by washes, nuclear counterstaining with Hoechst 33258, and mounting. Images were acquired on a Leica SP8 confocal microscope.

### RNA extraction, library preparation, and sequencing

Bulk RNAseq of the 814 biopsies processed at the NIH was performed on frozen muscle biopsy specimens as previously described ^6,22–26^. Briefly, muscle biopsies were immediately flash-frozen and stored at −80°C across all contributing centers. Samples were then transported in dry ice to the NIH and processed uniformly to prepare the library and conduct the analysis. RNA was extracted with TRIzol. Libraries were either prepared with the NeoPrep system according to the TruSeq Stranded mRNA Library Prep protocol (Illumina, San Diego, CA) or with the NEBNext Poly(A) mRNA Magnetic Isolation Module and Ultra^™^ II Directional RNA Library Prep Kit for Illumina (New England BioLabs, ref. #E7490, and #E7760).

Bulk RNAseq of the 41 biopsies processed independently at the University of Strasbourg was performed on frozen muscle biopsy specimens. Briefly, muscle biopsies obtained in France and Canada were immediately flash-frozen and stored at −80 °C, then transported on dry ice to the GenomEast platform (IGBMC, Strasbourg) for uniform processing. RNA was extracted using TRIzol. Stranded total RNA-seq libraries were prepared following ribosomal RNA depletion with the riboPOOL kit (siTOOLs Biotech). Libraries were generated using the MGIEasy Fast RNA Library Prep Set, including RNA fragmentation, strand-specific cDNA synthesis, adapter ligation, and PCR amplification. Library quality and quantity were assessed using Bioanalyzer 2100 and Qubit fluorimetry. Libraries were sequenced as paired-end 100 bp reads on an MGI DNBSEQ-G400RS platform.

### Electroporation of antibodies into human muscle cells

Human immunoglobulin G was purified and concentrated from serum using protein G Agarose (Millipore, ref. 16-266) and the Amicon Pro Purification System (Millipore, ref. ACS500024) with a 30kDa molecular weight cutoff Amicon Ultra Centrifugal Filter (Millipore, ref. UFC503024).

Normal human skeletal muscle myoblasts were cultured in growth medium and nucleofected with purified immunoglobulins according to the protocol recommended by the supplier (Lonza) and using the P5 Primary Cell 4D-Nucleofector™ X Kit L (Lonza, ref. V4XP-5024). Nucleofected cells were plated in differentiation medium and harvested for RNA extraction and subsequent RNA sequencing 24 and 72 hours after.

### Spatial transcriptomics

Spatial transcriptomics was performed using the 10x Genomics Xenium Human Multi-Tissue panel (∼380 genes) supplemented with a 100-gene custom add-on panel, following the manufacturer’s protocols for tissue preparation, probe hybridization, amplification, and imaging with the Xenium Analyzer. 5µm cryosections were mounted on Xenium slides and processed with the Xenium in situ gene expression workflow, including cell-segmentation staining, exactly as recommended by 10x Genomics. Combined probe sets were hybridized overnight, amplified through rolling-circle methods, and decoded through iterative fluorescence imaging to generate subcellular transcript-level maps. After automated onboard processing, spatially resolved gene counts, cell segmentation masks, and transcript coordinates were exported and analyzed using Xenium Explorer v4.1.1, which was used for quality control, visualization, and downstream spatial interpretation.

### Statistical and bioinformatic analysis

For RNA-seq analysis of samples processed at the NIH, sequencing reads were demultiplexed using bcl2fastq v2.20.0 and preprocessed with fastp v0.21.0. Gene-level abundances were quantified using Salmon v1.5.2, against a transcriptome indexed from the GRCh38 human genome assembly with GENCODE release 39 annotations.

Samples processed at the University of Strasbourg were preprocessed to remove adapter sequences, low-quality reads, and rRNA contaminants, and subsequently aligned to the GRCh38 human reference genome. Gene expression was quantified using STAR with Ensembl release 114 gene annotations.

For datasets generated at both institutions, raw counts were normalized using the Trimmed Mean of M-values (TMM) method implemented in edgeR v3.34.1 for graphical analyses. Differential expression analysis was performed using limma v3.48.3. Where applicable, P values were adjusted for multiple testing using the Benjamini–Hochberg procedure, and genes with a false discovery rate (q < 0.05) were considered statistically significant.

## Contributorship

All authors contributed to study conception and design, data interpretation, substantive manuscript review, and approval of the final draft.

## Acknowledgments

We thank Julie Thompson for coordinating and maintaining the NIH Natural History Protocol, and we acknowledge the NIAMS Sequencing and Imaging Core and its members for their technical support. Sequencing was performed by the GenomEast platform, a member of the “France Genomique” consortium (ANR-10-INBS-0009).

## Ethical approval

All biopsies were obtained under IRB-approved protocols from the National Institutes of Health, Johns Hopkins, Hospital Clinic, Vall d’Hebron Hospital, Mayo Clinic, Strasbourg University Hospitals, Centre hospitalier de l’Université de Montréal (CHUM), Charité-Universitätsmedizin Berlin, and Western University.

## Data sharing

Anonymized data not published within the article are available upon reasonable request to qualified investigators.

