## Supplementary Figures for "Spatial transcriptomics reveals mechanism of autoimmunity driven by internalized autoantibodies"

**Supplementary Figure 1. Flowchart illustrating the samples used across the different sections of the study.** Sample counts are reported as n (%). IF: immunofluorescence; NT, histologically normal muscle; DM, dermatomyositis; AS, antisynthetase syndrome; IBM, inclusion body myositis; ACA, anti-centromere autoantibodies; IM, inflammatory myopathies.

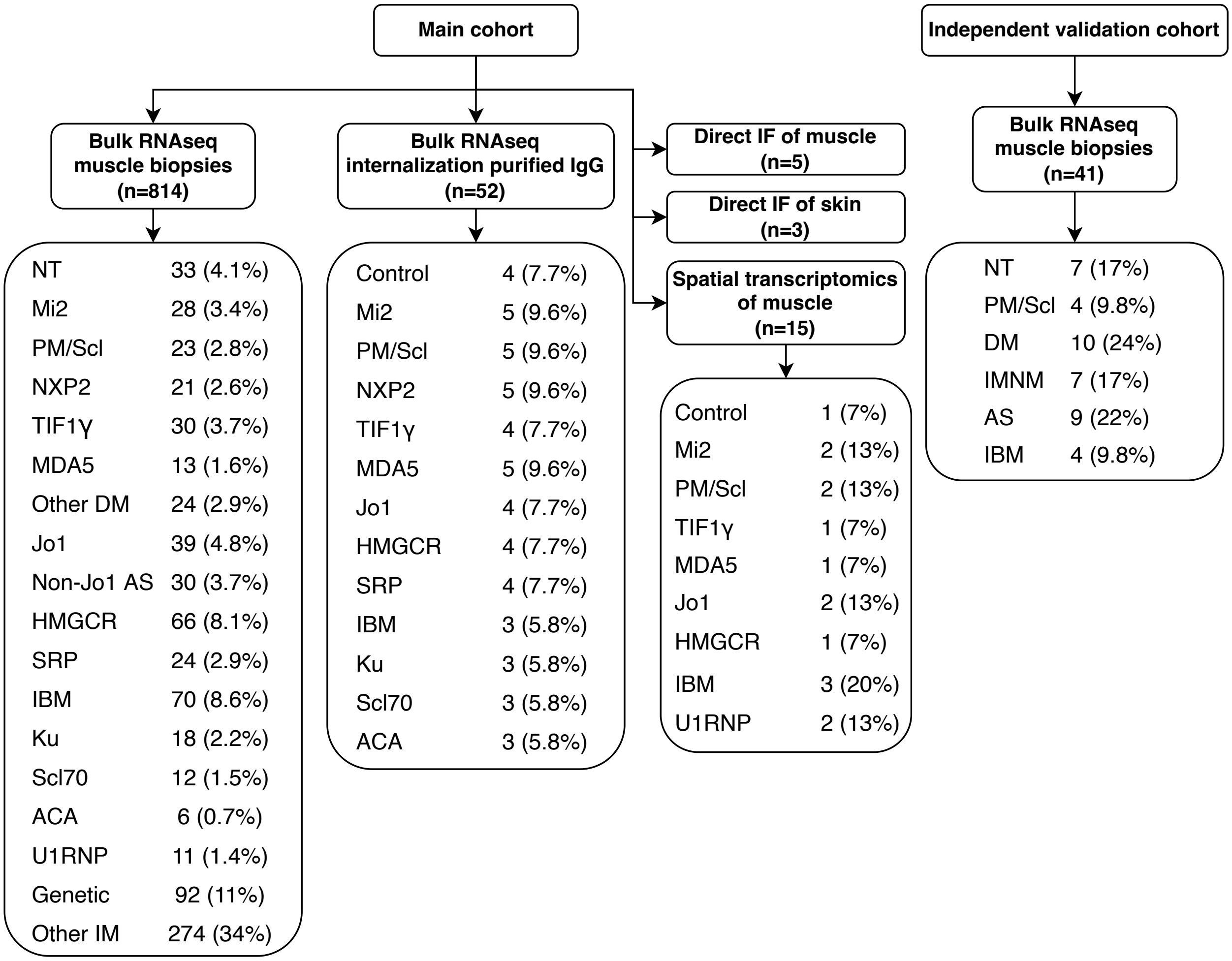

**Supplementary Figure 2.** Expression of anti-Mi2 (A) and anti-PM/Scl (B) associated gene signatures. Shown are the top 12 genes specifically overexpressed in muscle biopsies from anti-Mi2 (A) and anti-PM/Scl (B)-positive patients compared with all other groups included in the study, stratified by geographical biopsy location. Each dot represents the gene expression value from an individual patient. NT, histologically normal muscle; DM, dermatomyositis; AS, antisynthetase syndrome; IBM, inclusion body myositis; ACA, anti-centromere autoantibodies; IM, inflammatory myopathies.

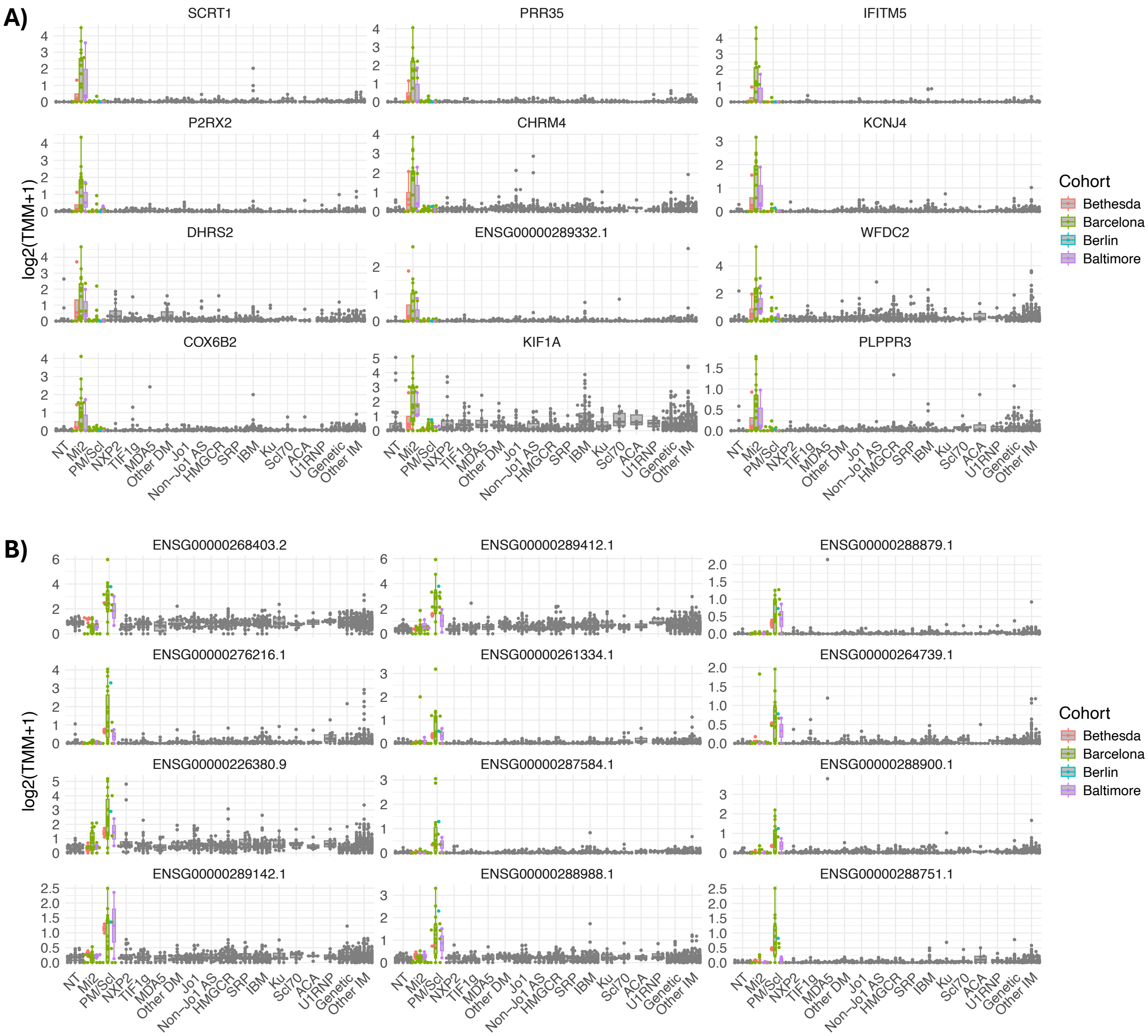

**Supplementary Figure 3.** Expression of anti-PM/Scl associated gene signatures across myositis subtypes and biopsy locations. Shown are the top 12 genes specifically overexpressed in muscle biopsies from anti-PM/Scl-positive patients compared with all other groups included in the study, stratified by geographical biopsy location. Each dot represents the gene expression value from an individual patient. NT, histologically normal muscle; DM, dermatomyositis; IMNM: immune-mediated necrotizing myositis; AS, antisynthetase syndrome; IBM, inclusion body myositis.

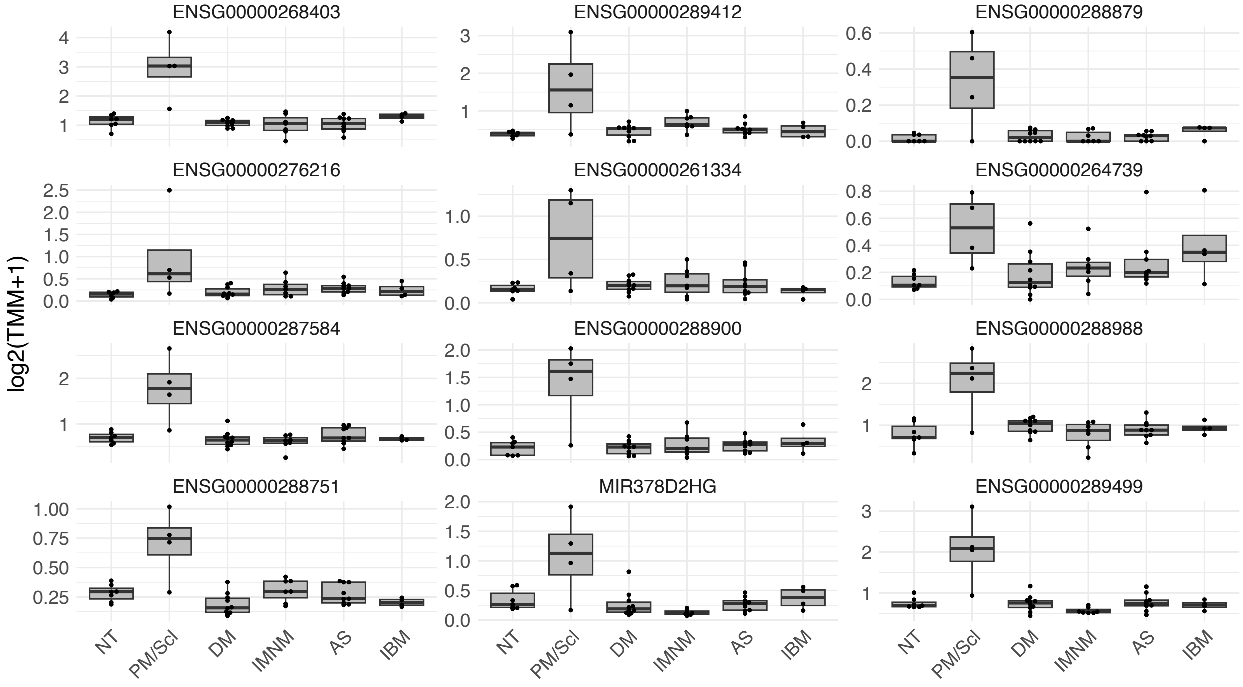

**Supplementary Figure 4. Anti-Mi2-specific gene expression in cultured muscle cells 24h after IgG electroporation.** Heatmap showing standardized expression levels (Z-scores) of the top 100 anti-Mi2-specific genes in human skeletal muscle cells 24h after electroporation with purified IgG from myositis patients and healthy controls. Each column represents IgG from an individual donor. Robust induction of anti-Mi2-specific gene expression is observed exclusively in cells electroporated with IgG from anti-Mi2-positive patients. IBM, inclusion body myositis; ACA, anti-centromere autoantibodies.

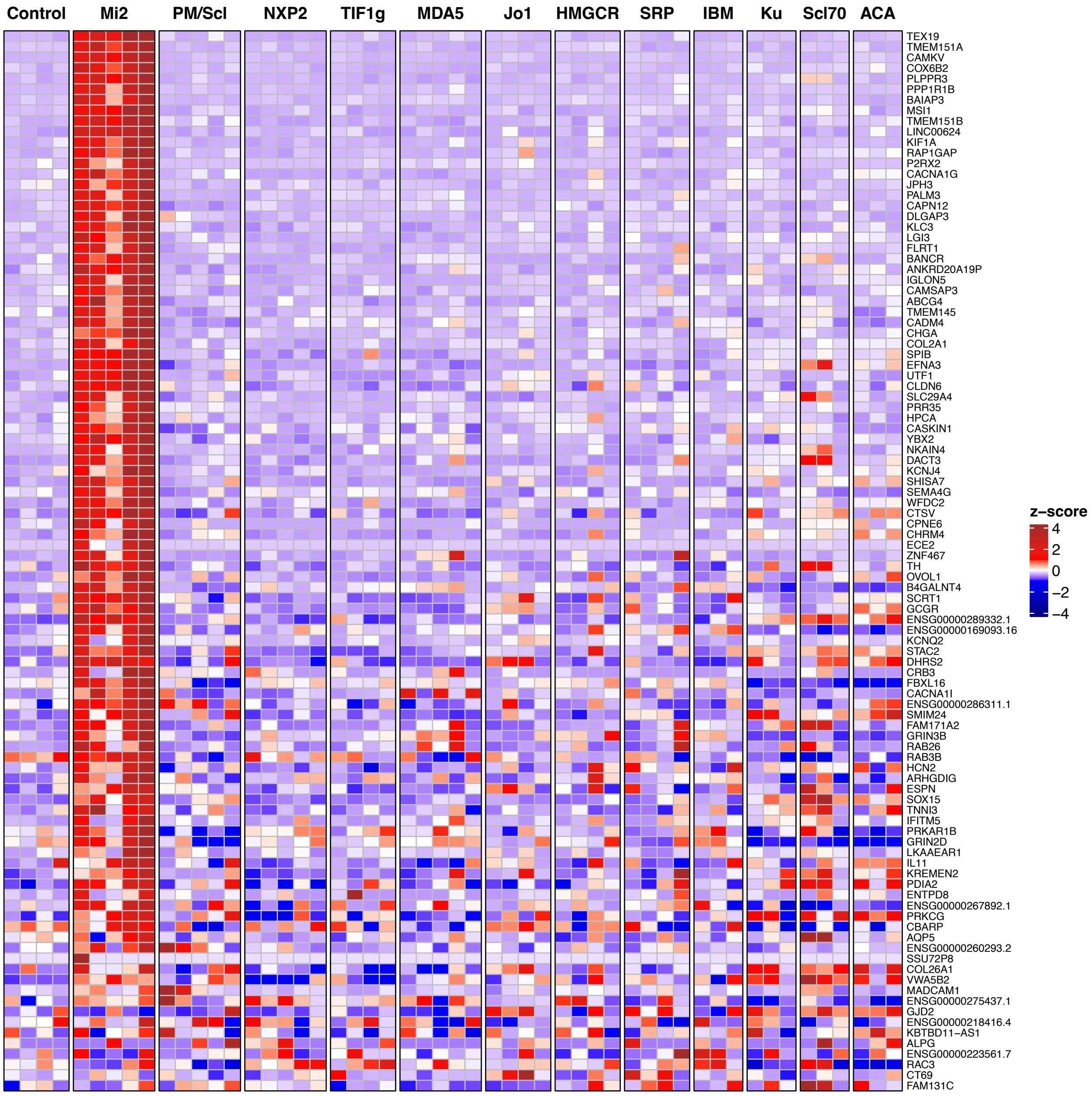

**Supplementary Figure 5. Anti-PM/Scl-specific gene expression in cultured muscle cells 72h after IgG electroporation.** Heatmap showing standardized expression levels (Z-scores) of the top 100 anti-PM/Scl-specific genes in human skeletal muscle cells 72h after electroporation with purified IgG from myositis patients and healthy controls. Each column represents IgG from an individual donor. Robust induction of anti-PM/Scl-specific gene expression is observed exclusively in cells electroporated with IgG from anti-PM/Scl-positive patients. IBM, inclusion body myositis; ACA, anti-centromere autoantibodies.

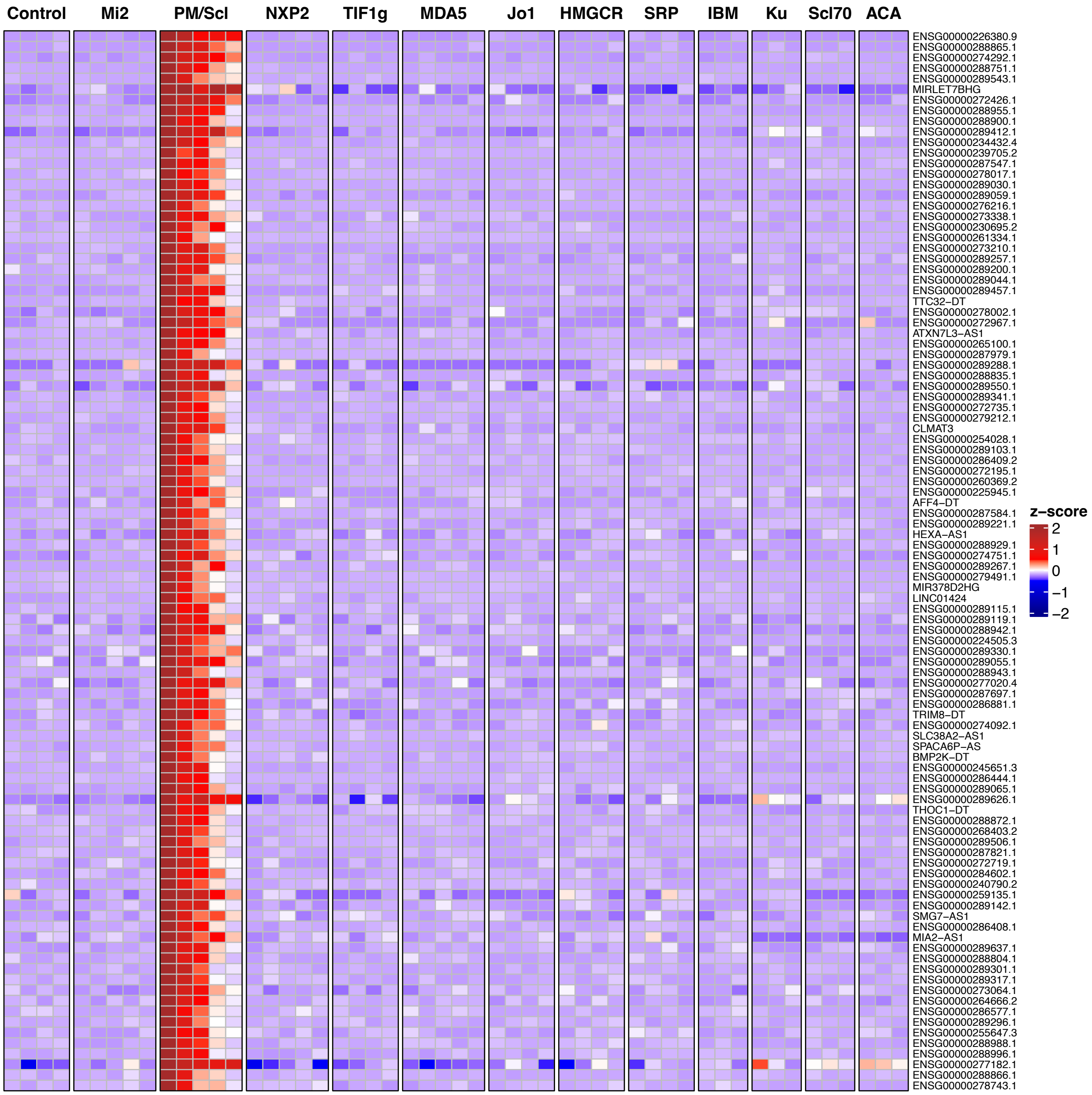

**Supplementary Figure 6.** Anti-Mi2-specific gene expression in cultured muscle cells 72h after IgG electroporation. Heatmap showing standardized expression levels (Z-scores) of the top 100 anti-Mi2-specific genes in human skeletal muscle cells 72h after electroporation with purified IgG from myositis patients and healthy controls. Each column represents IgG from an individual donor. Robust induction of anti-Mi2-specific gene expression is observed exclusively in cells electroporated with IgG from anti-Mi2-positive patients. IBM, inclusion body myositis; ACA, anti-centromere autoantibodies.

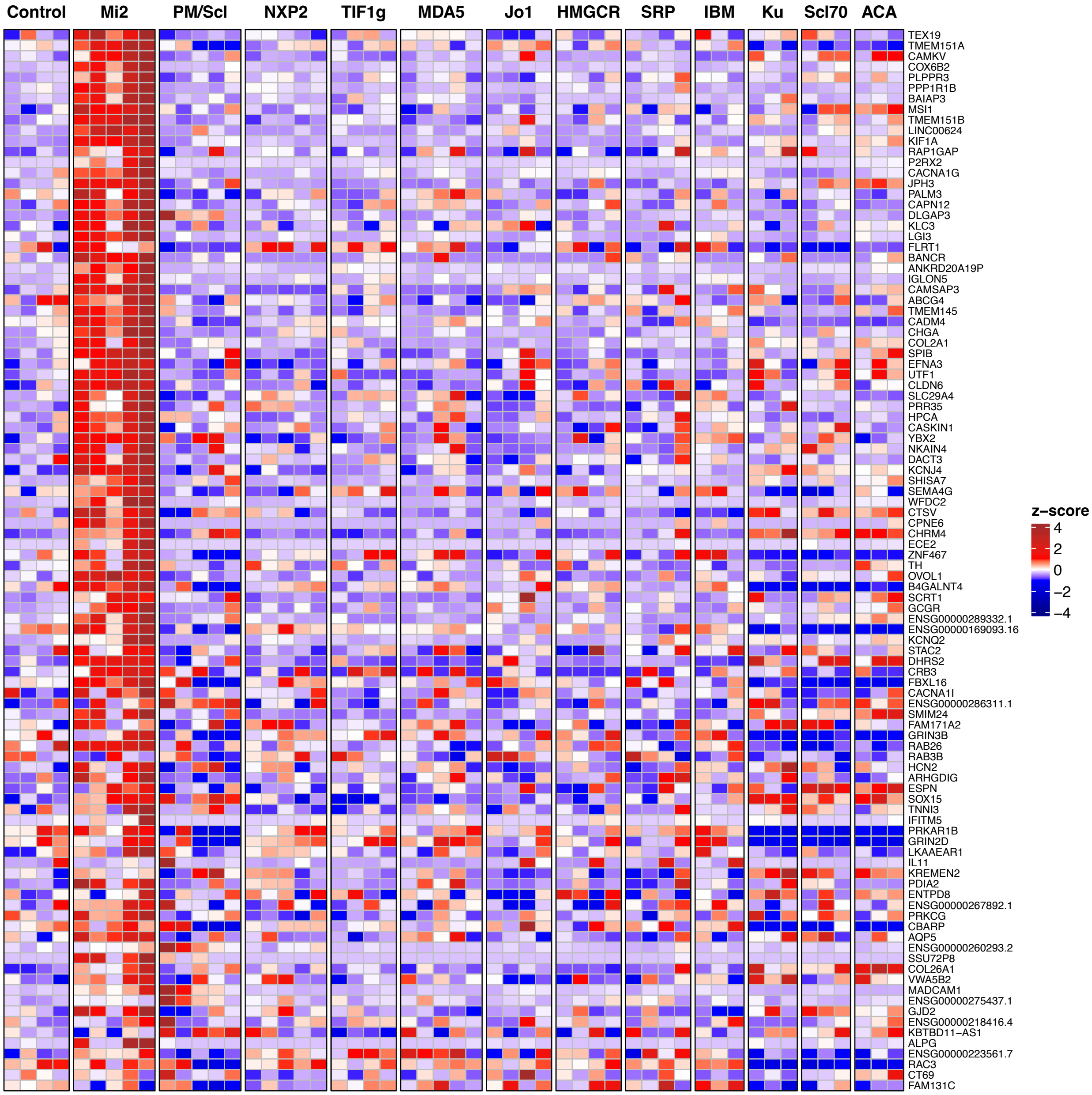

**Supplementary Figure 7.** Anti-PM/Scl-specific gene expression in cultured muscle cells 24h after IgG electroporation. Heatmap showing standardized expression levels (Z-scores) of the top 100 anti-PM/Scl-specific genes in human skeletal muscle cells 24h after electroporation with purified IgG from myositis patients and healthy controls. Each column represents IgG from an individual donor. Robust induction of anti-PM/Scl-specific gene expression is observed exclusively in cells electroporated with IgG from anti-PM/Scl-positive patients. IBM, inclusion body myositis; ACA, anti-centromere autoantibodies.

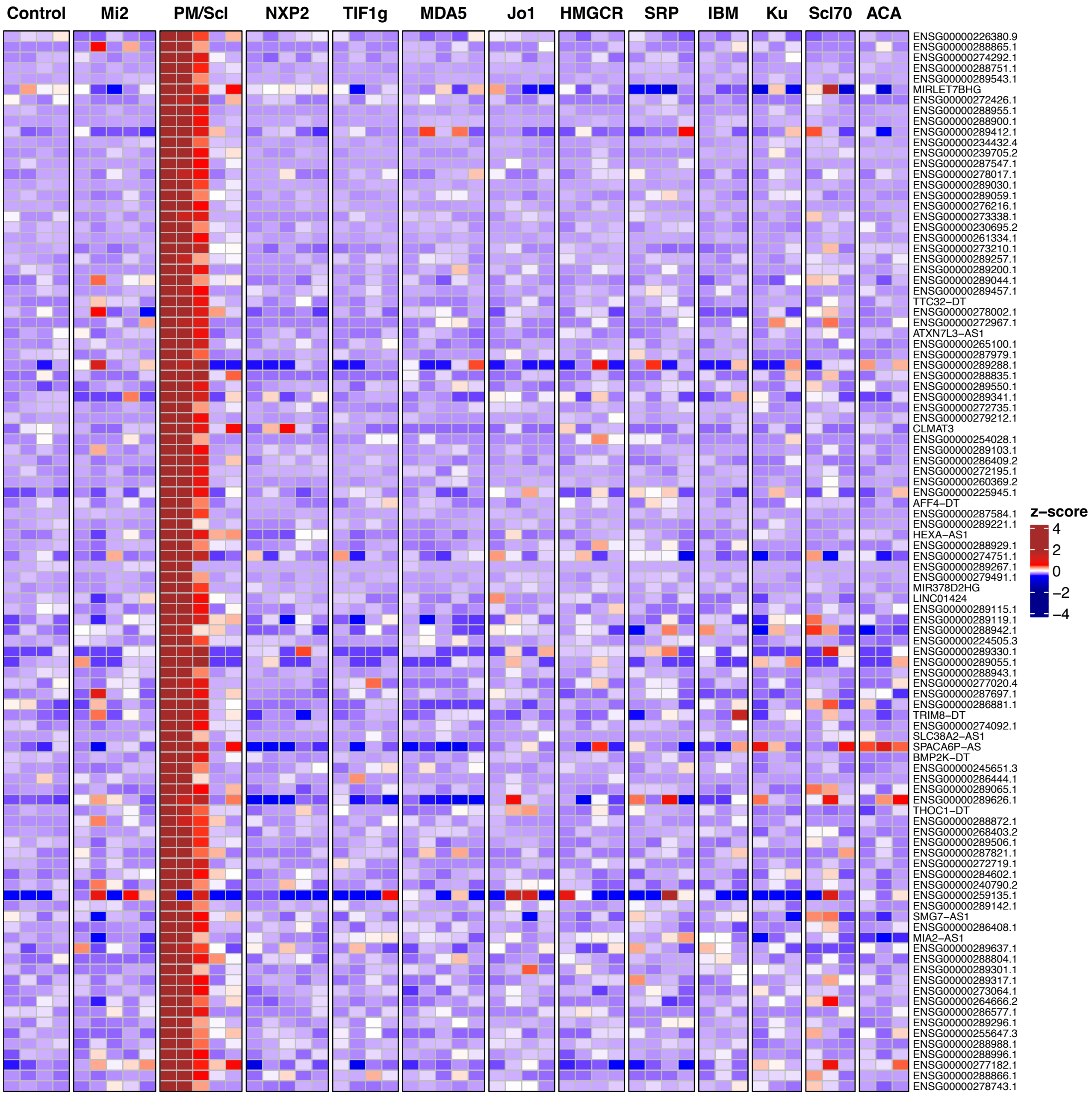

**Supplementary Figure 8.** Spatial transcriptomic analysis of a representative biopsy area from an anti-Mi2-positive patient, highlighting molecular signatures consistent with antibody internalization-associated tissue damage and inflammation and IgA-producing antibody-secreting cell externalization of immunoglobulin RNA. Colored dots indicate the spatial distribution of: (A) anti-Mi2-specific transcripts; (B) type I interferon-inducible genes (*ISG15*, *MX1*) and the interferon receptor *IFNAR1*; (C) transcripts associated with mature muscle fibers (*TTN*) and specific muscle fiber types (*MYH1*, type IIx; *MYH2*, type IIa; *MYH7*, type I); (D) *TGFB1* and *IL11*; (E) markers of muscle regeneration (*NCAM1*, *MYH3*, *MYH8*); and (F) plasma cell markers (*SDC1*), immunoglobulin A2 heavy-chain constant region (*IGHA2*), and immunoglobulin kappa constant region (*IGK*).

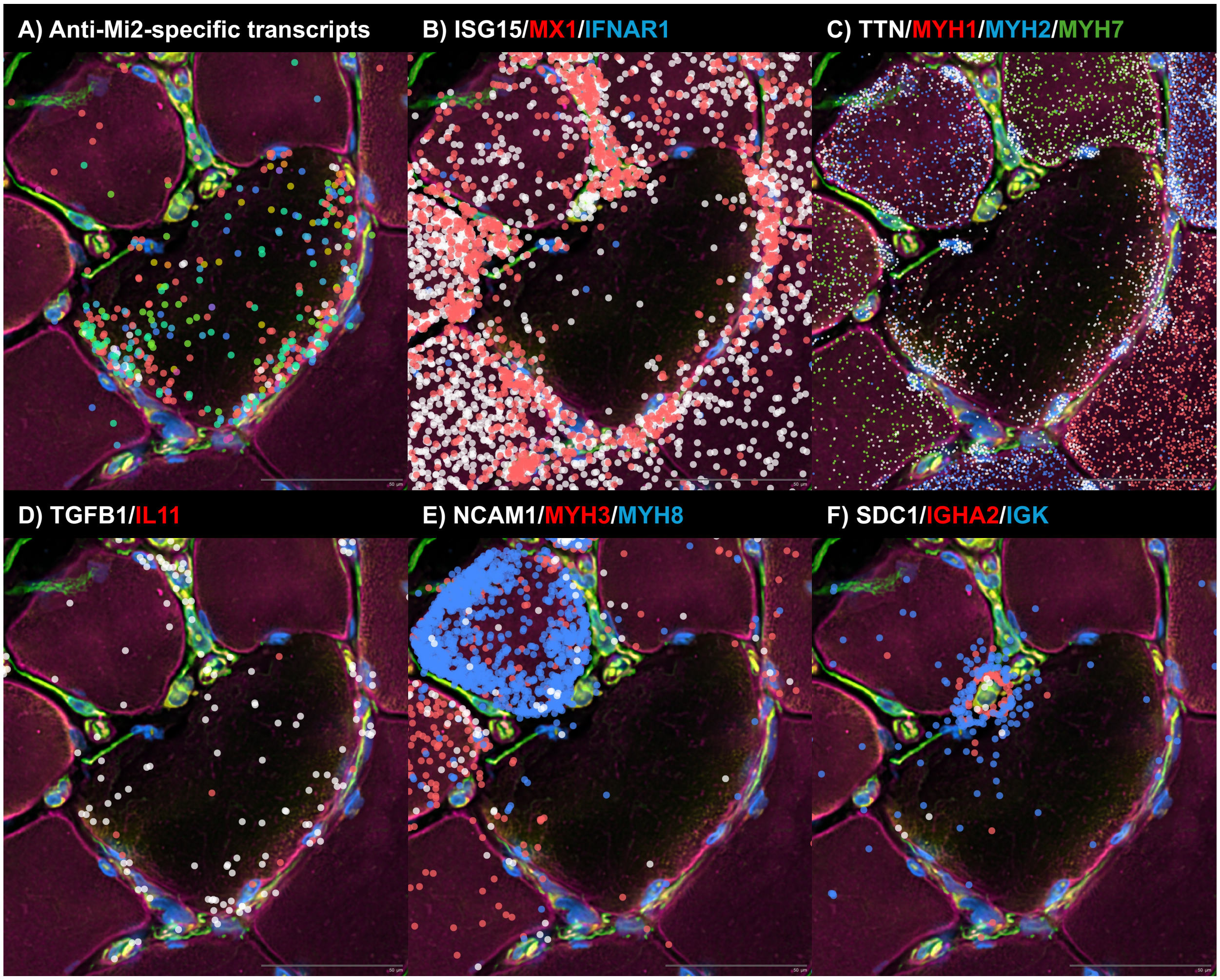

**Supplementary Figure 9.** Spatial transcriptomic analysis of a representative low-magnification region from an anti-Mi2-positive patient (white squares indicate biopsy regions highlighted in Figure 4 and Supplementary Figure 8), revealing molecular signatures consistent with antibody internalization-associated tissue damage, inflammation, and immunoglobulin RNA externalization by IgA-producing antibody-secreting cells. Colored dots, with size proportional to transcript abundance, depict the spatial distribution of transcripts.

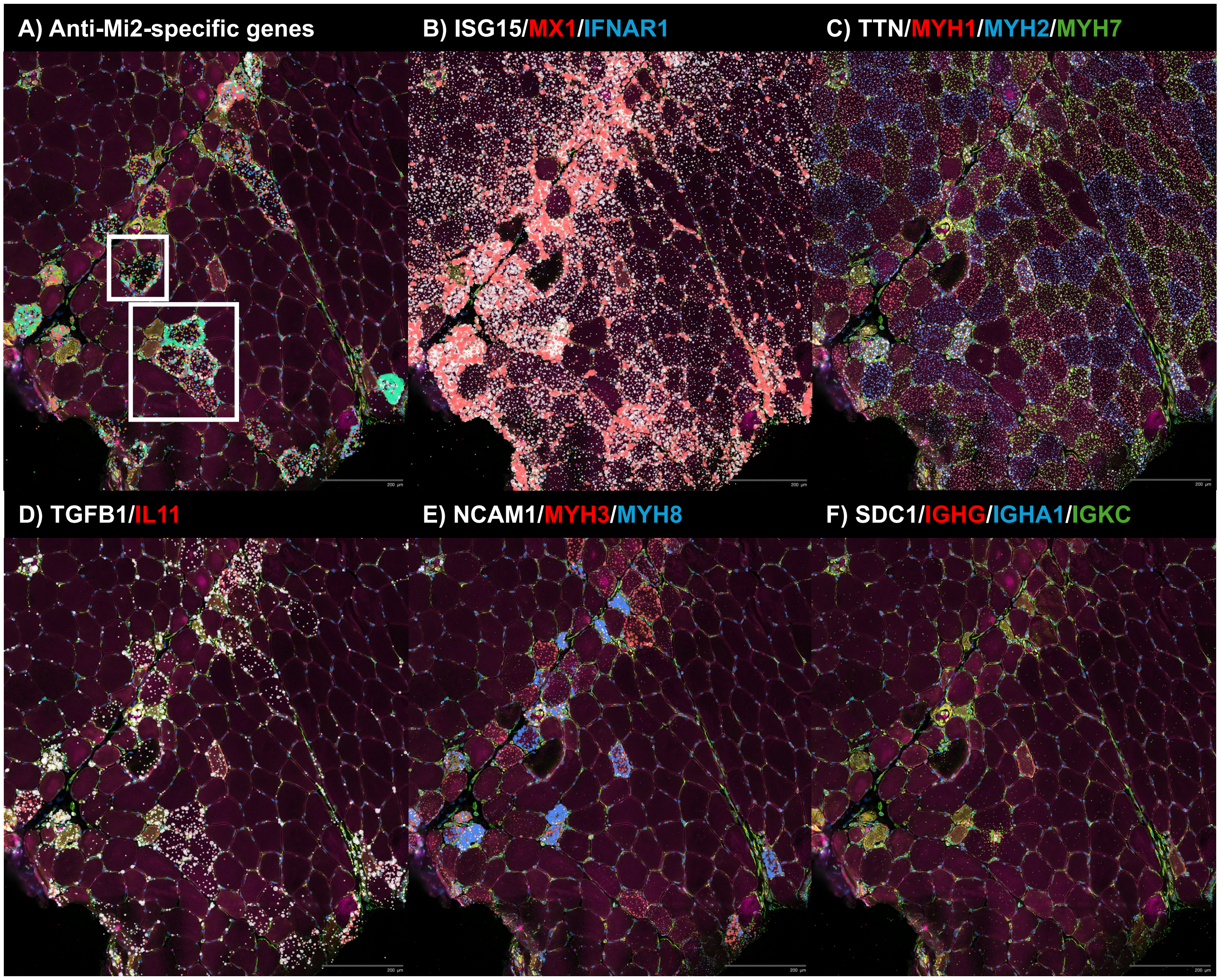

**Supplementary Figure 10.** Spatial transcriptomic analysis of representative low-magnification regions from an anti-Mi2-positive patient (A, B), an anti-PM/Scl-positive patient (C, D), and a healthy comparator (E, F). Selective expression of anti-Mi2-specific genes is observed in the anti-Mi2 biopsy (A), whereas anti-PM/Scl-specific transcripts are selectively detected in the anti-PM/Scl biopsy (D). No relevant expression of either signature is observed in the healthy comparator (E, F), and minimal expression of the alternative gene signature is detected in each disease biopsy.

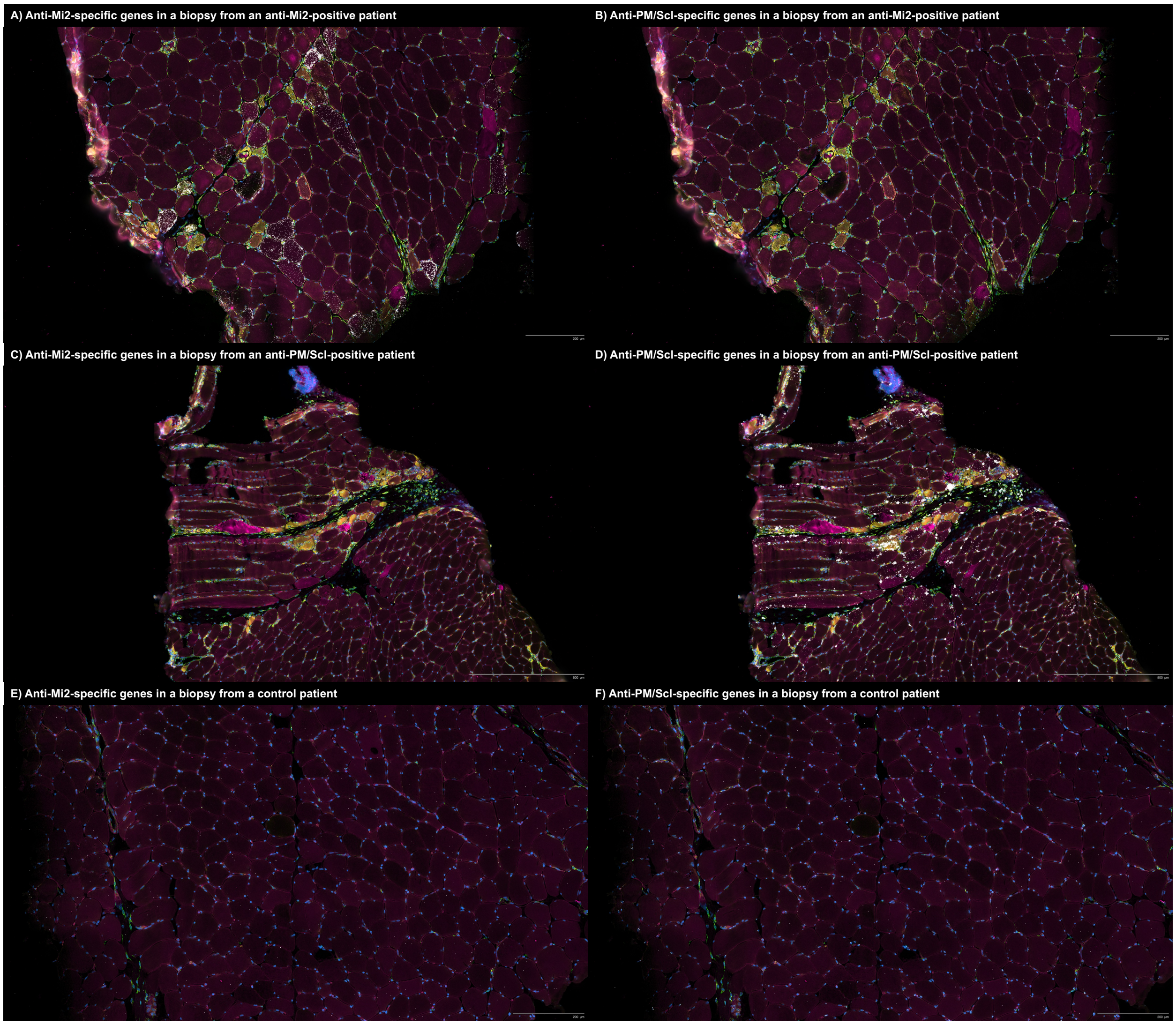

**Supplementary Figure 11.** Spatial transcriptomic analysis of a representative biopsy region from an anti-PM/Scl-positive patient reveals molecular signatures consistent with antibody internalization-associated tissue damage, type II interferon-mediated inflammation, and immunoglobulin RNA externalization in IgG-producing antibody-secreting cells. Colored dots denote the spatial distribution of: (A) anti-PM/Scl-specific transcripts; (B) transcripts marking mature muscle fibers (*TTN*) and muscle fiber subtypes (*MYH1*, type IIx; *MYH2*, type IIa; *MYH7*, type I); (C) type II interferon-inducible genes (*GBP2*, *CXCL9*); (D) markers of fibroblasts (*COL1A2*), macrophages (*CD163*), and T cells (*CD3D*); (E) B-cell markers immunoglobulin M heavy-chain constant region (*IGHM*) and *CD19*; and (F) antibody-secreting cell markers, including syndecan-1 (*SDC1*) and the immunoglobulin G heavy-chain constant region (*IGHG*).

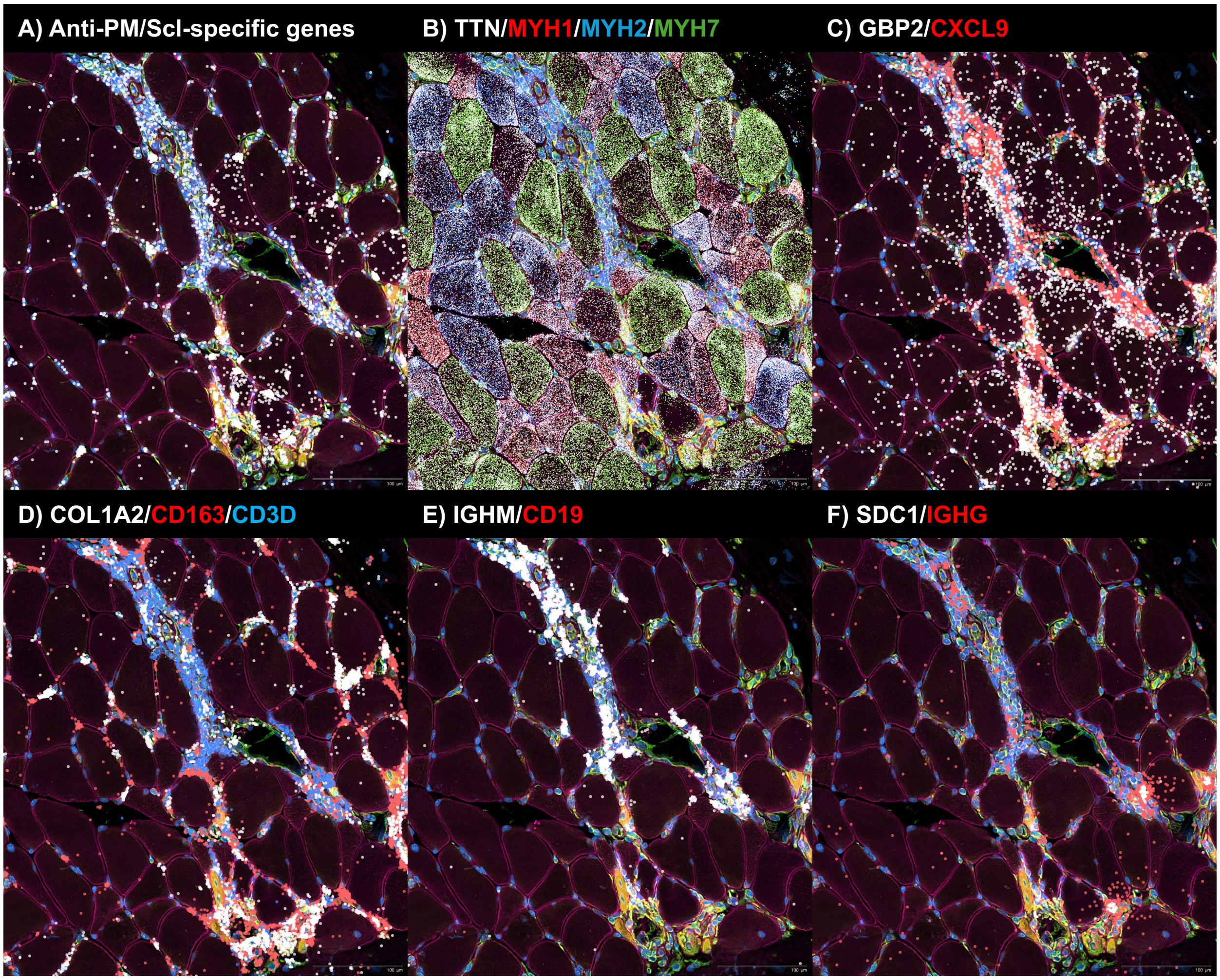

**Supplementary Figure 12.** Expression of IFNγ-inducible genes GBP2 and CXCL9 in different types of muscle diseases. The expression of the IFNγ-inducible genes GBP2 and CXCL9 is higher in anti-PM/Scl scleromyositis than in anti-Scl70 or anti-centromere systemic sclerosis. Each dot represents the gene expression value from an individual patient. NT, histologically normal muscle; DM, dermatomyositis; AS, antisynthetase syndrome; IBM, inclusion body myositis; ACA, anti-centromere autoantibodies; IM, inflammatory myopathies.

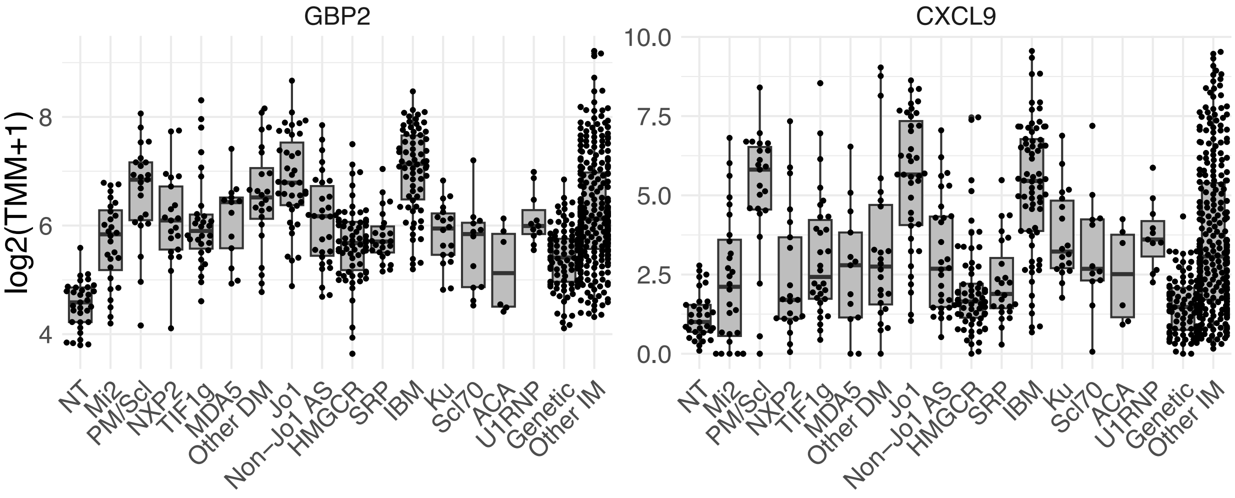

**Supplementary Figure 13.** Spatial transcriptomic analysis of representative biopsy regions from patients with anti-Mi2 and anti-PM/Scl autoantibodies. In an anti-Mi2-positive patient (A-C), anti-Mi2-specific genes (A) are overexpressed in IFNB1-expressing macrophages (B, C). In an anti-PM/Scl-positive patient (D, E), anti-PM/Scl-specific genes (D) are enriched in endothelial cells and fibroblasts (E).

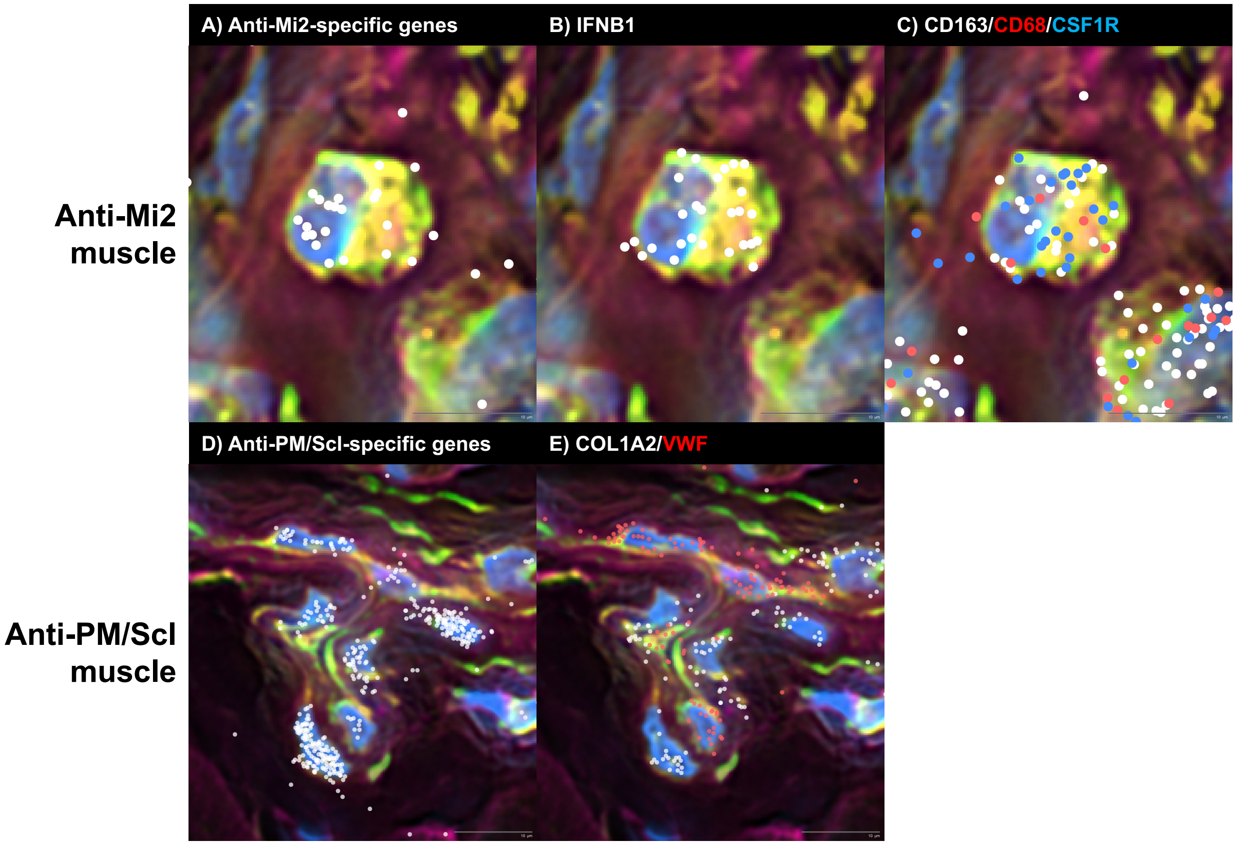

**Supplementary Figure 14.** Spatial transcriptomic analysis of a representative muscle biopsy from an anti-Mi2-positive patient demonstrates precise delineation of individual myofiber boundaries with minimal to no detectable transcript spillover into adjacent cells. Expression of *MYH1* (white; type 2X fibers), *MYH2* (red; type 2A fibers), and *MYH7* (blue; type 1 fibers) highlights fiber-type-specific localization. The magnified region shown in the lower left corresponds to the boxed area in the main image.

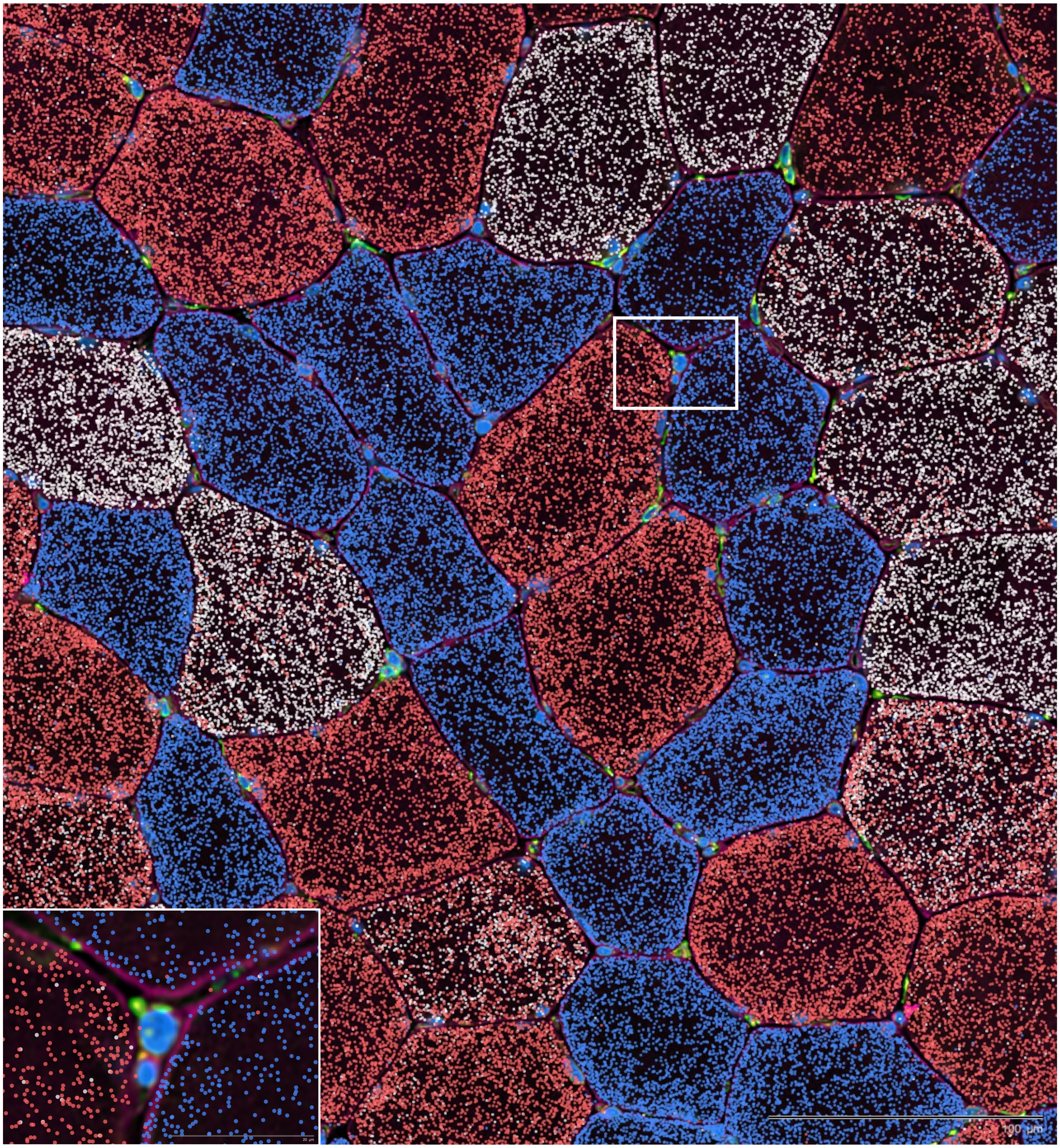

**Supplementary Figure 15. Independent validation of direct immunofluorescence with fixation from Canada demonstrates nuclear immunoglobulin deposition in muscle from patients with anti-U1RNP and anti-Ku autoantibodies.** Immunofluorescence of human IgG shows antibody deposition in the nuclei of muscle fibers in anti-Ku (A) and anti-U1RNP myositis (C-D), but not in normal muscle (B). In D, DAPI-stained nuclei (arrows) appear in blue and confirm nuclear localization of IgG deposition. Box in C shows speckled pattern of IgG deposits in a large, internalized muscle fiber nucleus. Scale bars: A–B, 100 µm; C-D, 25 24µm.

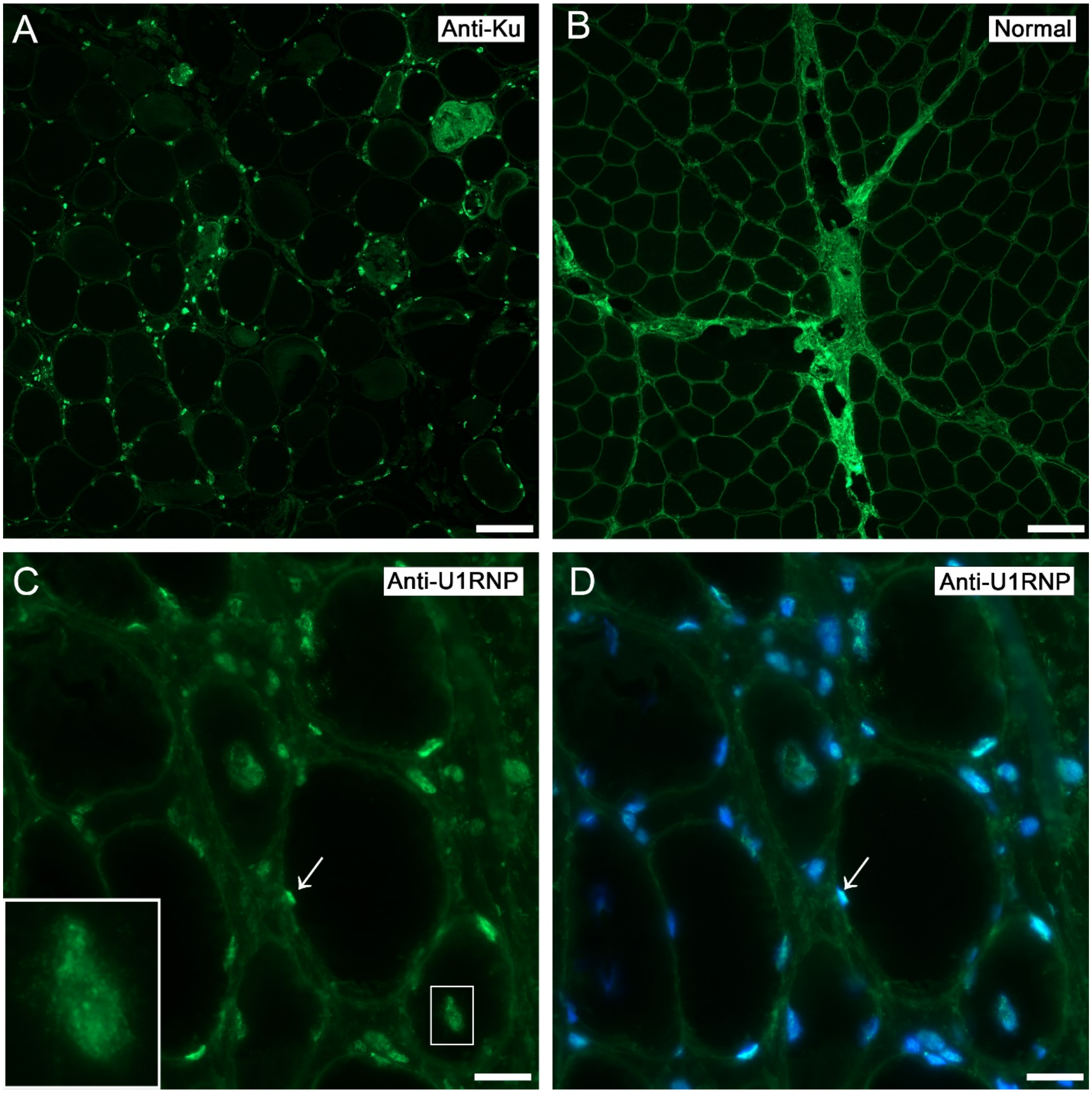
