## Supplementary Tables for "Spatial transcriptomics reveals mechanism of autoimmunity driven by internalized autoantibodies"

**Supplementary Table 1. Differential expression of anti-Mi2-specific genes in muscle biopsies of anti-Mi2-positive patients and anti-PM/Scl-positive patients relative to all the other samples.** Pos, position in the complete differential expression table; logFC, $log_2$ fold change; adj.P.Val, Benjamini-Hochberg-adjusted P value.

| | Anti-Mi2 vs. all | | | Anti-PM/Scl vs. all | | |
|---|---|---|---|---|---|---|
| **Gene** | **Pos** | **logFC** | **adj.P.Val** | **Pos** | **logFC** | **adj.P.Val** |
| PRR35 | 1 | 3.9 | 2.5e-32 | 20,640 | 0.4 | 4.3e-01 |
| IFITM5 | 2 | 3.8 | 2.9e-32 | 36,321 | -0.1 | 8.4e-01 |
| SCRT1 | 3 | 4.1 | 5.0e-32 | 33,329 | 0.2 | 7.7e-01 |
| P2RX2 | 5 | 3.8 | 6.9e-30 | 19,464 | 0.5 | 3.9e-01 |
| DHRS2 | 11 | 3.4 | 2.8e-20 | 43,901 | 0.0 | 9.7e-01 |
| CHRM4 | 13 | 2.9 | 5.8e-20 | 42,428 | -0.1 | 9.4e-01 |
| KCNJ4 | 16 | 3.2 | 1.8e-18 | 42,665 | 0.1 | 9.5e-01 |
| COX6B2 | 18 | 3.0 | 2.3e-17 | 35,506 | 0.2 | 8.2e-01 |
| RAB3B | 23 | 3.0 | 1.0e-16 | 8,263 | 1.1 | 7.5e-02 |
| ENSG00000289332.1 | 28 | 2.7 | 9.9e-15 | 41,843 | 0.1 | 9.3e-01 |
| PLPPR3 | 31 | 2.7 | 3.8e-14 | 41,517 | -0.1 | 9.3e-01 |
| WFDC2 | 38 | 2.5 | 8.9e-13 | 27,373 | -0.4 | 6.2e-01 |
| TMEM151A | 48 | 2.4 | 6.1e-12 | 33,711 | 0.2 | 7.8e-01 |
| KIF1A | 49 | 2.7 | 6.3e-12 | 23,004 | -0.6 | 5.0e-01 |
| ENSG00000255375.3 | 56 | 2.4 | 2.3e-11 | 36,872 | 0.1 | 8.5e-01 |
| KREMEN2 | 69 | 2.8 | 3.4e-11 | 41,385 | -0.1 | 9.3e-01 |
| ANKRD20A19P | 95 | 2.4 | 2.6e-10 | 20,493 | 0.5 | 4.2e-01 |
| COL2A1 | 149 | 2.2 | 3.6e-09 | 42,905 | 0.1 | 9.5e-01 |
| ECE2 | 161 | 2.1 | 4.5e-09 | 26,432 | 0.3 | 6.0e-01 |
| IGLON5 | 220 | 2.2 | 1.5e-08 | 6,184 | 1.0 | 3.9e-02 |
| FAM171A2 | 230 | 1.7 | 1.6e-08 | 20,374 | 0.4 | 4.2e-01 |
| CAMKV | 243 | 2.1 | 1.8e-08 | 10,673 | 0.8 | 1.3e-01 |
| CLDN6 | 365 | 2.1 | 8.9e-08 | 44,856 | -0.0 | 9.8e-01 |
| ENSG00000286311.1 | 407 | 1.9 | 1.2e-07 | 35,885 | 0.2 | 8.3e-01 |
| FBXL16 | 416 | 2.0 | 1.3e-07 | 34,888 | -0.2 | 8.1e-01 |
| B4GALNT4 | 476 | 1.8 | 2.5e-07 | 40,237 | -0.1 | 9.1e-01 |
| TMEM151B | 522 | 1.9 | 3.9e-07 | 7,772 | 0.9 | 6.5e-02 |
| ENSG00000251511.1 | 533 | 1.5 | 4.2e-07 | 31,396 | 0.2 | 7.3e-01 |
| PTH2 | 560 | 1.4 | 4.9e-07 | 41,308 | 0.1 | 9.3e-01 |
| CPNE6 | 575 | 2.1 | 5.5e-07 | 24,094 | 0.4 | 5.3e-01 |
| SHISA7 | 755 | 2.0 | 1.5e-06 | 27,415 | 0.3 | 6.2e-01 |
| LKAAEAR1 | 764 | 1.7 | 1.6e-06 | 17,248 | 0.5 | 3.2e-01 |
| GIMD1 | 789 | 1.4 | 1.9e-06 | 9,779 | 0.6 | 1.1e-01 |
| NKAIN4 | 853 | 2.1 | 2.5e-06 | 37,213 | -0.2 | 8.5e-01 |
| GJD2 | 902 | 2.0 | 3.1e-06 | 18,502 | -0.6 | 3.6e-01 |
| GCGR | 931 | 2.1 | 3.5e-06 | 22,377 | 0.5 | 4.8e-01 |
| PRKCG | 968 | 1.8 | 4.1e-06 | 16,296 | 0.6 | 2.9e-01 |
| ENSG00000256481.1 | 1,002 | 1.6 | 4.8e-06 | 33,689 | 0.2 | 7.8e-01 |
| BANCR | 1,053 | 1.6 | 6.1e-06 | 30,304 | 0.2 | 7.0e-01 |
| MMEL1-AS1 | 1,148 | 1.3 | 8.4e-06 | 22,848 | 0.3 | 4.9e-01 |
| OVOL1 | 1,330 | 1.7 | 1.5e-05 | 33,390 | 0.2 | 7.7e-01 |
| SMIM24 | 1,379 | 1.7 | 1.7e-05 | 29,643 | 0.3 | 6.8e-01 |
| TNNI3 | 1,646 | 1.4 | 3.4e-05 | 20,297 | -0.5 | 4.2e-01 |
| HCN2 | 1,763 | 1.5 | 4.5e-05 | 9,100 | 0.8 | 9.2e-02 |
| ALPG | 1,851 | 1.8 | 5.5e-05 | 1,302 | 1.8 | 4.2e-04 |
| CT69 | 1,918 | 1.5 | 6.4e-05 | 27,704 | -0.3 | 6.3e-01 |
| ENSG00000142539.9 | 1,966 | 2.0 | 6.8e-05 | 4,367 | 1.5 | 1.6e-02 |
| C5ORF47 | 2,152 | 1.4 | 9.8e-05 | 39,707 | -0.1 | 9.0e-01 |
| CACNA1G | 2,183 | 1.3 | 1.1e-04 | 24,912 | -0.4 | 5.5e-01 |
| TEX19 | 2,207 | 1.5 | 1.1e-04 | 38,342 | 0.1 | 8.7e-01 |
| IL11 | 2,233 | 1.5 | 1.2e-04 | 22,338 | 0.4 | 4.8e-01 |
| COL26A1 | 2,238 | 1.5 | 1.2e-04 | 9,059 | 0.9 | 9.1e-02 |
| FAM131C | 2,304 | 1.5 | 1.4e-04 | 25,062 | -0.4 | 5.6e-01 |
| ESPN | 2,461 | 1.5 | 1.8e-04 | 16,727 | 0.6 | 3.0e-01 |
| CHGA | 2,679 | 1.5 | 2.6e-04 | 26,193 | 0.3 | 5.9e-01 |
| LINC02066 | 2,714 | 1.2 | 2.7e-04 | 16,800 | 0.5 | 3.1e-01 |
| ENSG00000251076.1 | 2,778 | 1.7 | 3.0e-04 | 26,879 | -0.4 | 6.1e-01 |
| UOX | 2,811 | 1.3 | 3.1e-04 | 31,473 | -0.2 | 7.3e-01 |

| | Anti-Mi2 vs. all | | | Anti-PM/Scl vs. all | | |
|---|---|---|---|---|---|---|
| **Gene** | **Pos** | **logFC** | **adj.P.Val** | **Pos** | **logFC** | **adj.P.Val** |
| **LINC00624** | 3,044 | 1.3 | 4.3e-04 | 37,463 | -0.1 | 8.6e-01 |
| **ENTPD8** | 3,237 | 1.4 | 5.7e-04 | 24,051 | 0.4 | 5.3e-01 |
| **TH** | 3,347 | 1.3 | 6.5e-04 | 36,567 | 0.1 | 8.4e-01 |
| **SSU72P8** | 3,383 | 1.1 | 6.8e-04 | 1,539 | 1.3 | 7.9e-04 |
| **RAC3** | 3,573 | 1.1 | 8.5e-04 | 20,171 | 0.4 | 4.2e-01 |
| **RAP1GAP** | 3,660 | 1.2 | 9.3e-04 | 13,683 | -0.7 | 2.1e-01 |
| **KLC3** | 3,767 | 1.3 | 1.0e-03 | 31,908 | 0.3 | 7.4e-01 |
| **CAPN12** | 3,929 | 1.1 | 1.2e-03 | 43,666 | -0.0 | 9.6e-01 |
| **C1QTNF8** | 4,174 | 1.3 | 1.5e-03 | 23,133 | 0.4 | 5.0e-01 |
| **GRIN2D** | 4,331 | 1.1 | 1.7e-03 | 34,095 | -0.2 | 7.9e-01 |
| **KCNQ2** | 5,061 | 1.3 | 3.2e-03 | 38,536 | -0.1 | 8.8e-01 |
| **CTSV** | 5,241 | 0.7 | 3.6e-03 | 14,994 | -0.4 | 2.5e-01 |
| **GRIN3B** | 5,546 | 1.2 | 4.6e-03 | 3,309 | 1.3 | 7.7e-03 |
| **FLRT1** | 5,935 | 1.1 | 6.1e-03 | 45,852 | 0.0 | 1.0e+00 |
| **PALM3** | 5,979 | 1.0 | 6.3e-03 | 26,955 | -0.3 | 6.1e-01 |
| **JPH3** | 6,304 | 1.0 | 7.9e-03 | 42,715 | -0.1 | 9.5e-01 |
| **SPIB** | 6,580 | 0.8 | 9.4e-03 | 4,060 | 0.9 | 1.4e-02 |
| **CAMSAP3** | 6,687 | 1.1 | 1.0e-02 | 38,196 | 0.1 | 8.7e-01 |
| **UTF1** | 6,805 | 0.9 | 1.1e-02 | 37,411 | 0.1 | 8.6e-01 |
| **PPP1R1B** | 6,881 | 1.4 | 1.1e-02 | 11,735 | -1.2 | 1.6e-01 |
| **CBARP** | 6,888 | 0.7 | 1.1e-02 | 10,642 | 0.6 | 1.3e-01 |
| **CASKIN1** | 7,086 | 0.9 | 1.3e-02 | 23,155 | -0.4 | 5.0e-01 |
| **HPCA** | 7,287 | 0.9 | 1.4e-02 | 17,836 | 0.5 | 3.4e-01 |
| **SEMA4G** | 7,448 | 0.6 | 1.5e-02 | 4,317 | -0.8 | 1.6e-02 |
| **LGI3** | 8,003 | 1.0 | 2.0e-02 | 19,788 | -0.5 | 4.0e-01 |
| **ENSG00000218416.4** | 8,066 | 0.9 | 2.0e-02 | 45,289 | 0.0 | 9.9e-01 |
| **AQP5** | 8,729 | 0.9 | 2.7e-02 | 21,512 | 0.4 | 4.5e-01 |
| **SMPD4P1** | 8,854 | 1.0 | 2.8e-02 | 34,461 | 0.2 | 8.0e-01 |
| **CACNA1I** | 8,931 | 0.9 | 2.9e-02 | 38,365 | 0.1 | 8.7e-01 |
| **ENSG00000260293.2** | 9,144 | 0.8 | 3.2e-02 | 15,413 | 0.5 | 2.7e-01 |
| **CRB3** | 9,914 | 0.8 | 4.4e-02 | 45,033 | -0.0 | 9.9e-01 |
| **MSI1** | 10,015 | 0.8 | 4.5e-02 | 29,875 | 0.3 | 6.9e-01 |
| **PDIA2** | 10,063 | 0.8 | 4.6e-02 | 15,196 | -0.6 | 2.6e-01 |
| **MADCAM1** | 10,489 | 0.6 | 5.3e-02 | 10,038 | 0.6 | 1.2e-01 |
| **VWA5B2** | 10,720 | 0.8 | 5.7e-02 | 19,911 | 0.5 | 4.1e-01 |
| **STAC2** | 12,426 | 0.7 | 9.5e-02 | 8,663 | 0.8 | 8.3e-02 |
| **ARHGDIG** | 12,727 | 0.8 | 1.0e-01 | 10,228 | -0.9 | 1.2e-01 |
| **SLC29A4** | 13,376 | 0.6 | 1.2e-01 | 4,157 | -1.3 | 1.5e-02 |
| **ENSG00000275437.1** | 13,527 | 0.6 | 1.2e-01 | 7,970 | 0.8 | 6.9e-02 |
| **PRKAR1B** | 15,145 | 0.3 | 1.7e-01 | 13,091 | 0.3 | 2.0e-01 |
| **KBTBD11-AS1** | 16,067 | 0.5 | 2.1e-01 | 41,669 | 0.1 | 9.3e-01 |
| **ABCG4** | 18,233 | 0.4 | 2.9e-01 | 21,429 | 0.4 | 4.5e-01 |
| **EFNA3** | 19,086 | -0.4 | 3.3e-01 | 27,852 | -0.2 | 6.3e-01 |
| **TMEM145** | 19,372 | 0.4 | 3.4e-01 | 9,193 | -0.7 | 9.5e-02 |
| **BAIAP3** | 25,949 | 0.2 | 6.3e-01 | 5,807 | -0.7 | 3.4e-02 |
| **ENSG00000267892.1** | 26,756 | 0.2 | 6.6e-01 | 20,577 | 0.3 | 4.3e-01 |
| **ENSG00000223561.7** | 29,623 | 0.2 | 7.7e-01 | 7,522 | -0.9 | 6.1e-02 |
| **RAB26** | 31,400 | -0.2 | 8.2e-01 | 26,383 | -0.3 | 6.0e-01 |
| **DACT3** | 31,684 | -0.1 | 8.3e-01 | 19,613 | 0.3 | 4.0e-01 |
| **YBX2** | 35,516 | 0.1 | 9.2e-01 | 33,614 | -0.1 | 7.8e-01 |
| **SOX15** | 38,072 | -0.1 | 9.5e-01 | 4,889 | -1.0 | 2.2e-02 |
| **ENSG00000169093.16** | 38,229 | 0.0 | 9.5e-01 | 16,785 | -0.3 | 3.1e-01 |
| **DLGAP3** | 39,181 | 0.1 | 9.6e-01 | 19,453 | 0.4 | 3.9e-01 |
| **ZNF467** | 40,148 | -0.0 | 9.7e-01 | 45,843 | 0.0 | 1.0e+00 |
| **CADM4** | 40,341 | 0.0 | 9.7e-01 | 8,798 | -0.5 | 8.5e-02 |

**Supplementary Table 2. Differential expression of anti-PM/Scl-specific genes (PMID 38902010) in muscle biopsies of anti-PM/Scl-positive patients and anti-Mi2-positive patients relative to all the other samples.** Pos, position in the complete differential expression table; logFC, $log_2$ fold change; adj.P.Val, Benjamini-Hochberg-adjusted P value.

| | Anti-PM/Scl vs. all | | | Anti-Mi2 vs. all | | |
|---|---|---|---|---|---|---|
| **Gene** | **Pos** | **logFC** | **adj.P.Val** | **Pos** | **logFC** | **adj.P.Val** |
| **ENSG00000268403.2** | 1 | 2.6 | 2.2e-33 | 740 | -1.5 | 1.4e-06 |
| **ENSG00000276216.1** | 2 | 4.5 | 1.3e-31 | 12,611 | -0.8 | 9.9e-02 |
| **ENSG00000289412.1** | 3 | 2.7 | 4.4e-31 | 2,557 | -1.3 | 2.1e-04 |
| **ENSG00000288879.1** | 4 | 3.6 | 7.6e-26 | 43,206 | -0.0 | 9.9e-01 |
| **ENSG00000288751.1** | 5 | 3.7 | 3.3e-25 | 25,338 | -0.3 | 6.1e-01 |
| **ENSG00000287584.1** | 6 | 3.5 | 2.1e-24 | 19,495 | -0.4 | 3.4e-01 |
| **ENSG00000226380.9** | 7 | 2.7 | 2.6e-23 | 36,171 | -0.1 | 9.3e-01 |
| **ENSG00000289296.1** | 8 | 3.4 | 2.4e-22 | 10,028 | -0.7 | 4.6e-02 |
| **MIR378D2HG** | 9 | 3.3 | 1.7e-21 | 20,814 | -0.4 | 4.0e-01 |
| **ENSG00000288988.1** | 10 | 2.7 | 2.6e-21 | 14,972 | -0.6 | 1.7e-01 |
| **ENSG00000261334.1** | 11 | 3.5 | 1.8e-20 | 44,744 | 0.0 | 9.9e-01 |
| **ENSG00000289030.1** | 12 | 3.3 | 4.9e-20 | 12,825 | -0.6 | 1.0e-01 |
| **ENSG00000264739.1** | 13 | 3.4 | 1.1e-18 | 32,691 | -0.2 | 8.5e-01 |
| **ENSG00000273210.1** | 14 | 2.4 | 1.8e-18 | 24,175 | 0.3 | 5.5e-01 |
| **ENSG00000289341.1** | 15 | 3.3 | 2.3e-18 | 16,718 | -0.6 | 2.3e-01 |
| **ENSG00000288943.1** | 16 | 3.5 | 6.1e-18 | 14,566 | -0.6 | 1.6e-01 |
| **ENSG00000289103.1** | 17 | 3.1 | 1.6e-17 | 4,136 | -1.1 | 1.5e-03 |
| **ENSG00000289499.1** | 18 | 3.0 | 2.7e-17 | 12,500 | -0.6 | 9.6e-02 |
| **ENSG00000288900.1** | 19 | 3.3 | 5.3e-17 | 17,660 | -0.5 | 2.7e-01 |
| **ENSG00000234432.4** | 20 | 2.5 | 5.5e-17 | 13,187 | -0.7 | 1.1e-01 |
| **ENSG00000288865.1** | 21 | 2.9 | 1.1e-16 | 28,333 | -0.2 | 7.2e-01 |
| **ENSG00000287979.1** | 23 | 3.4 | 1.1e-16 | 17,226 | -0.7 | 2.5e-01 |
| **ENSG00000239705.2** | 25 | 2.8 | 1.6e-16 | 25,954 | -0.2 | 6.3e-01 |
| **ENSG00000289221.1** | 26 | 2.9 | 2.0e-16 | 24,269 | -0.3 | 5.6e-01 |
| **ENSG00000289115.1** | 27 | 2.8 | 2.2e-16 | 26,280 | -0.2 | 6.4e-01 |
| **ENSG00000272967.1** | 28 | 2.8 | 8.0e-16 | 17,163 | -0.5 | 2.5e-01 |
| **TRIM8-DT** | 29 | 2.0 | 1.2e-15 | 4,792 | -1.0 | 2.6e-03 |
| **BMP2K-DT** | 30 | 2.8 | 7.8e-15 | 5,770 | -1.2 | 5.4e-03 |
| **ENSG00000288919.1** | 31 | 2.7 | 9.5e-15 | 2,131 | -1.7 | 9.5e-05 |
| **LINC01126** | 32 | 2.4 | 1.6e-14 | 9,080 | -0.8 | 3.1e-02 |
| **TIMMDC1-DT** | 33 | 2.7 | 1.6e-14 | 28,667 | -0.2 | 7.3e-01 |
| **ENSG00000260369.2** | 34 | 2.7 | 2.4e-14 | 14,839 | -0.6 | 1.7e-01 |
| **ENSG00000272719.1** | 35 | 2.7 | 4.7e-14 | 19,904 | -0.4 | 3.6e-01 |
| **ENSG00000286409.2** | 36 | 2.7 | 4.7e-14 | 13,122 | -0.5 | 1.1e-01 |
| **ENSG00000279259.1** | 37 | 2.4 | 5.0e-14 | 7,544 | -0.9 | 1.6e-02 |
| **ENSG00000288955.1** | 38 | 2.7 | 5.7e-14 | 20,880 | -0.3 | 4.0e-01 |
| **ENSG00000287654.1** | 39 | 2.6 | 6.3e-14 | 25,537 | -0.3 | 6.1e-01 |
| **ENSG00000289200.1** | 40 | 2.7 | 4.2e-13 | 40,881 | -0.0 | 9.7e-01 |
| **ENSG00000265100.1** | 41 | 2.6 | 1.3e-12 | 15,645 | -0.6 | 1.9e-01 |
| **UAP1-DT** | 42 | 2.4 | 1.3e-12 | 27,735 | -0.2 | 7.0e-01 |
| **ENSG00000289202.1** | 43 | 2.7 | 2.0e-12 | 19,159 | -0.4 | 3.3e-01 |
| **ENSG00000288804.1** | 45 | 2.5 | 2.4e-12 | 16,181 | -0.5 | 2.1e-01 |
| **ENSG00000289044.1** | 46 | 2.7 | 2.6e-12 | 14,130 | -0.5 | 1.4e-01 |
| **ENSG00000289142.1** | 47 | 2.2 | 3.8e-12 | 23,540 | -0.3 | 5.2e-01 |
| **ENSG00000288996.1** | 48 | 2.4 | 3.9e-12 | 31,325 | -0.2 | 8.2e-01 |
| **ENSG00000289288.1** | 49 | 2.6 | 4.2e-12 | 32,002 | 0.2 | 8.4e-01 |
| **MIR23AHG** | 50 | 1.2 | 4.5e-12 | 11,078 | -0.4 | 6.4e-02 |
| **ENSG00000289152.1** | 52 | 2.5 | 6.1e-12 | 34,381 | -0.1 | 9.0e-01 |
| **ENSG00000286408.1** | 53 | 1.9 | 6.2e-12 | 3,172 | -1.1 | 5.2e-04 |
| **TTC32-DT** | 54 | 2.5 | 6.3e-12 | 17,194 | -0.4 | 2.5e-01 |
| **ENSG00000289379.1** | 55 | 2.6 | 7.5e-12 | 31,827 | -0.1 | 8.3e-01 |

| | Anti-PM/Scl vs. all | | | Anti-Mi2 vs. all | | |
|---|---|---|---|---|---|---|
| **Gene** | **Pos** | **logFC** | **adj.P.Val** | **Pos** | **logFC** | **adj.P.Val** |
| **ENSG00000288963.1** | 56 | 2.5 | 1.1e-11 | 22,085 | -0.3 | 4.6e-01 |
| **ENSG00000279212.1** | 57 | 2.3 | 1.2e-11 | 11,772 | -0.7 | 7.9e-02 |
| **ENSG00000289159.1** | 58 | 2.3 | 1.4e-11 | 2,449 | -1.5 | 1.7e-04 |
| **ENSG00000289154.1** | 59 | 1.7 | 1.6e-11 | 3,454 | -1.0 | 7.4e-04 |
| **ENSG00000288872.1** | 61 | 2.4 | 1.9e-11 | 21,618 | -0.3 | 4.4e-01 |
| **ENSG00000289551.1** | 62 | 2.4 | 3.2e-11 | 31,639 | -0.1 | 8.3e-01 |
| **LINC00677** | 63 | 2.3 | 3.8e-11 | 44,449 | 0.0 | 9.9e-01 |
| **TNFAIP8L1** | 64 | 1.0 | 3.8e-11 | 26,390 | 0.1 | 6.5e-01 |
| **ENSG00000270019.1** | 66 | 2.6 | 5.9e-11 | 37,236 | -0.1 | 9.4e-01 |
| **ENSG00000289055.1** | 68 | 2.5 | 1.4e-10 | 21,962 | -0.3 | 4.5e-01 |
| **ENSG00000289303.1** | 70 | 2.4 | 1.8e-10 | 32,361 | -0.1 | 8.5e-01 |
| **SLC38A2-AS1** | 71 | 2.4 | 1.8e-10 | 40,653 | -0.0 | 9.7e-01 |
| **ENSG00000261242.1** | 72 | 2.3 | 2.4e-10 | 28,825 | -0.2 | 7.4e-01 |
| **THOC1-DT** | 73 | 1.9 | 2.4e-10 | 2,959 | -1.1 | 3.8e-04 |
| **TERC** | 74 | 2.7 | 3.9e-10 | 33,904 | -0.1 | 8.9e-01 |
| **ENSG00000289235.1** | 75 | 2.3 | 5.0e-10 | 34,172 | -0.1 | 8.9e-01 |
| **SPACA6P-AS** | 76 | 2.5 | 6.3e-10 | 11,328 | -0.7 | 6.9e-02 |
| **ENSG00000289478.1** | 77 | 2.6 | 7.8e-10 | 28,259 | -0.2 | 7.2e-01 |
| **ENSG00000287821.1** | 79 | 2.1 | 9.7e-10 | 35,055 | 0.1 | 9.1e-01 |
| **ENSG00000245651.3** | 80 | 2.3 | 9.9e-10 | 14,183 | -0.6 | 1.4e-01 |
| **ENSG00000241666.2** | 82 | 2.2 | 1.6e-09 | 20,379 | -0.4 | 3.8e-01 |
| **ENSG00000286113.1** | 85 | 2.4 | 2.2e-09 | 41,761 | -0.0 | 9.8e-01 |
| **ENSG00000288939.1** | 87 | 2.3 | 2.3e-09 | 16,391 | -0.5 | 2.2e-01 |
| **ENSG00000289550.1** | 88 | 2.3 | 2.6e-09 | 16,091 | -0.6 | 2.1e-01 |
| **ENSG00000224505.3** | 89 | 1.4 | 2.7e-09 | 609 | -1.4 | 6.9e-07 |
| **ENSG00000284484.1** | 90 | 2.7 | 3.0e-09 | 12,957 | 0.8 | 1.1e-01 |
| **ENSG00000288744.1** | 92 | 2.3 | 3.5e-09 | 21,588 | -0.3 | 4.3e-01 |
| **RNVU1-14** | 93 | 2.4 | 3.9e-09 | 36,863 | -0.1 | 9.4e-01 |
| **ENSG00000255089.1** | 97 | 2.3 | 4.3e-09 | 18,671 | -0.4 | 3.1e-01 |
| **MSRA-DT** | 101 | 2.3 | 7.5e-09 | 13,762 | -0.6 | 1.3e-01 |
| **ENSG00000286577.1** | 104 | 2.1 | 9.6e-09 | 32,344 | -0.1 | 8.5e-01 |
| **ENSG00000289317.1** | 106 | 2.0 | 1.0e-08 | 27,881 | -0.2 | 7.0e-01 |
| **ENSG00000272768.1** | 107 | 2.0 | 1.1e-08 | 17,274 | 0.5 | 2.5e-01 |
| **ENSG00000289506.1** | 109 | 2.0 | 1.2e-08 | 25,578 | -0.3 | 6.2e-01 |
| **ENSG00000284602.1** | 110 | 1.4 | 1.2e-08 | 4,042 | -0.9 | 1.4e-03 |
| **ENSG00000272426.1** | 111 | 2.2 | 1.3e-08 | 16,711 | -0.5 | 2.3e-01 |
| **ENSG00000274213.1** | 112 | 2.1 | 1.3e-08 | 14,195 | -0.7 | 1.4e-01 |
| **ENSG00000282936.2** | 113 | 2.3 | 1.5e-08 | 8,511 | -0.9 | 2.4e-02 |
| **TRIM51BP** | 114 | 2.6 | 1.6e-08 | 21,559 | 0.4 | 4.3e-01 |
| **ENSG00000255647.3** | 115 | 2.2 | 1.6e-08 | 41,156 | -0.0 | 9.7e-01 |
| **ENSG00000288842.1** | 119 | 2.1 | 1.9e-08 | 19,144 | -0.4 | 3.3e-01 |
| **ENSG00000272953.1** | 121 | 1.6 | 3.0e-08 | 1,290 | -1.4 | 1.3e-05 |
| **MIR5188** | 122 | 1.9 | 3.1e-08 | 45,085 | -0.0 | 1.0e+00 |
| **ENSG00000273338.1** | 125 | 2.6 | 3.6e-08 | 19,582 | -0.5 | 3.4e-01 |
| **ENSG00000253838.1** | 127 | 2.1 | 3.6e-08 | 29,758 | 0.2 | 7.7e-01 |
| **ENSG00000259135.1** | 129 | 2.2 | 3.8e-08 | 9,358 | -0.8 | 3.5e-02 |
| **SMG7-AS1** | 131 | 2.0 | 3.9e-08 | 13,052 | -0.6 | 1.1e-01 |
| **PRAMEF13** | 133 | 2.4 | 4.1e-08 | 26,978 | 0.3 | 6.7e-01 |
| **ENSG00000288929.1** | 134 | 2.4 | 4.6e-08 | 11,545 | -0.7 | 7.3e-02 |
| **FSCN1** | 139 | 1.3 | 5.3e-08 | 369 | 1.2 | 9.0e-08 |
| **RABEP2** | 140 | 0.8 | 6.1e-08 | 42,537 | 0.0 | 9.8e-01 |
| **DDX39B-AS1** | 141 | 2.0 | 6.3e-08 | 19,955 | -0.4 | 3.6e-01 |
| **ENSG00000278743.1** | 142 | 1.9 | 6.4e-08 | 23,365 | -0.4 | 5.1e-01 |
| **LINC01089** | 153 | 0.8 | 1.1e-07 | 19,113 | -0.2 | 3.3e-01 |
| **MIRLET7BHG** | 164 | 1.2 | 1.4e-07 | 2,798 | -1.0 | 3.1e-04 |
| **ENSG00000287070.1** | 166 | 2.3 | 1.5e-07 | 11,854 | -0.8 | 8.1e-02 |
| **ENSG00000288866.1** | 167 | 2.1 | 1.5e-07 | 30,386 | -0.2 | 7.9e-01 |

| | Anti-PM/Scl vs. all | | | Anti-Mi2 vs. all | | |
|---|---|---|---|---|---|---|
| **Gene** | **Pos** | **logFC** | **adj.P.Val** | **Pos** | **logFC** | **adj.P.Val** |
| **ENSG00000286444.1** | 168 | 2.0 | 1.6e-07 | 35,673 | 0.1 | 9.2e-01 |
| **ENSG00000274751.1** | 173 | 1.7 | 1.9e-07 | 6,863 | -0.9 | 1.1e-02 |
| **ENSG00000289177.1** | 177 | 2.1 | 2.0e-07 | 35,133 | -0.1 | 9.1e-01 |
| **ENSG00000289229.1** | 179 | 2.0 | 2.0e-07 | 40,050 | -0.1 | 9.7e-01 |
| **ENSG00000289182.1** | 180 | 1.5 | 2.1e-07 | 16,272 | -0.5 | 2.1e-01 |
| **ENSG00000288896.1** | 181 | 2.0 | 2.1e-07 | 21,197 | -0.4 | 4.2e-01 |
| **ENSG00000276524.1** | 182 | 1.9 | 2.2e-07 | 12,585 | -0.6 | 9.9e-02 |
| **LINC01424** | 184 | 1.7 | 2.4e-07 | 12,310 | -0.6 | 9.2e-02 |
| **ENSG00000268670.1** | 190 | 2.0 | 2.5e-07 | 11,800 | -0.7 | 8.0e-02 |
| **ENSG00000254028.1** | 191 | 2.0 | 2.6e-07 | 16,406 | -0.4 | 2.2e-01 |
| **CAPN10-DT** | 196 | 1.3 | 2.8e-07 | 661 | -1.4 | 8.9e-07 |
| **RENO1** | 198 | 1.1 | 2.8e-07 | 492 | -1.3 | 2.8e-07 |
| **ENSG00000289257.1** | 207 | 1.9 | 3.5e-07 | 40,249 | 0.0 | 9.7e-01 |
| **AFF4-DT** | 211 | 1.9 | 3.6e-07 | 35,793 | 0.1 | 9.2e-01 |
| **PRND** | 212 | 2.4 | 3.6e-07 | 7,192 | -1.4 | 1.3e-02 |
| **ENSG00000286482.1** | 222 | 1.7 | 4.6e-07 | 7,773 | -0.8 | 1.7e-02 |
| **LINC00896** | 224 | 2.0 | 4.6e-07 | 18,577 | -0.4 | 3.0e-01 |
| **ENSG00000289457.1** | 229 | 2.0 | 5.0e-07 | 5,073 | -1.0 | 3.2e-03 |
| **ENSG00000279491.1** | 231 | 1.7 | 5.1e-07 | 1,916 | -1.4 | 6.3e-05 |
| **ENSG00000273064.1** | 232 | 1.6 | 5.2e-07 | 12,614 | -0.6 | 9.9e-02 |
| **ENSG00000287697.1** | 236 | 1.8 | 5.7e-07 | 18,920 | -0.4 | 3.2e-01 |
| **CPEB2-DT** | 253 | 2.0 | 8.1e-07 | 40,730 | -0.0 | 9.7e-01 |
| **ENSG00000289005.1** | 259 | 1.6 | 9.3e-07 | 4,528 | -1.0 | 2.0e-03 |
| **ENSG00000288813.1** | 271 | 1.9 | 1.2e-06 | 33,664 | -0.1 | 8.8e-01 |
| **LINC02776** | 272 | 1.9 | 1.2e-06 | 44,036 | 0.0 | 9.9e-01 |
| **ENSG00000278158.1** | 276 | 1.9 | 1.4e-06 | 16,987 | -0.4 | 2.4e-01 |
| **SDR42E2** | 281 | 2.0 | 1.5e-06 | 25,980 | -0.3 | 6.3e-01 |
| **ENSG00000286881.1** | 285 | 1.9 | 1.6e-06 | 22,759 | -0.3 | 4.9e-01 |
| **ENSG00000283959.2** | 295 | 1.7 | 1.9e-06 | 17,958 | -0.5 | 2.8e-01 |
| **ANKH-DT** | 296 | 1.8 | 2.0e-06 | 8,322 | -0.8 | 2.2e-02 |
| **COL18A1** | 310 | 0.9 | 2.5e-06 | 21,567 | 0.2 | 4.3e-01 |
| **KLF2-DT** | 311 | 1.9 | 2.6e-06 | 34,117 | -0.1 | 8.9e-01 |
| **CAGE1** | 315 | 2.0 | 2.6e-06 | 26,017 | -0.3 | 6.3e-01 |
| **ENSG00000272948.2** | 320 | 1.6 | 2.7e-06 | 14,695 | -0.6 | 1.6e-01 |
| **ENSG00000274292.1** | 333 | 1.7 | 3.2e-06 | 16,350 | -0.6 | 2.2e-01 |
| **ATXN7L3-AS1** | 334 | 1.9 | 3.2e-06 | 25,528 | -0.3 | 6.1e-01 |
| **ENSG00000276744.1** | 336 | 1.7 | 3.2e-06 | 12,715 | -0.6 | 1.0e-01 |
| **ENSG00000289626.1** | 355 | 1.2 | 4.3e-06 | 4,481 | -0.9 | 1.9e-03 |
| **CLMAT3** | 369 | 1.8 | 5.1e-06 | 16,032 | -0.4 | 2.1e-01 |
| **ENSG00000257258.2** | 374 | 1.7 | 5.5e-06 | 24,579 | 0.3 | 5.7e-01 |
| **ENSG00000278002.1** | 379 | 1.6 | 6.0e-06 | 1,909 | -1.4 | 6.2e-05 |
| **ENSG00000273363.1** | 382 | 1.7 | 6.1e-06 | 19,579 | -0.4 | 3.4e-01 |
| **ENSG00000269399.2** | 384 | 1.4 | 6.2e-06 | 1,338 | -1.4 | 1.5e-05 |
| **ENSG00000287547.1** | 393 | 1.6 | 7.2e-06 | 33,432 | -0.1 | 8.7e-01 |
| **ENSG00000289637.1** | 403 | 2.0 | 7.7e-06 | 12,180 | -0.6 | 8.8e-02 |
| **ENSG00000289301.1** | 431 | 1.7 | 9.4e-06 | 18,184 | -0.4 | 2.9e-01 |
| **ENSG00000289059.1** | 436 | 1.7 | 9.6e-06 | 14,603 | -0.6 | 1.6e-01 |
| **ENSG00000260651.1** | 453 | 1.8 | 1.2e-05 | 14,899 | -0.5 | 1.7e-01 |
| **ENSG00000288835.1** | 484 | 1.7 | 1.5e-05 | 33,600 | -0.1 | 8.8e-01 |
| **ENSG00000289547.1** | 488 | 1.9 | 1.6e-05 | 27,793 | 0.3 | 7.0e-01 |
| **ENSG00000275709.1** | 502 | 1.7 | 1.8e-05 | 39,268 | 0.1 | 9.6e-01 |
| **MIR1915HG** | 503 | 1.3 | 1.8e-05 | 11,042 | -0.7 | 6.3e-02 |
| **ENSG00000289031.1** | 504 | 1.7 | 1.8e-05 | 17,651 | -0.4 | 2.7e-01 |
| **ENSG00000225945.1** | 509 | 1.6 | 1.8e-05 | 39,336 | 0.1 | 9.6e-01 |
| **ENSG00000278017.1** | 545 | 1.6 | 2.3e-05 | 4,406 | -1.1 | 1.8e-03 |
| **ENSG00000289230.1** | 550 | 1.8 | 2.4e-05 | 15,081 | -0.6 | 1.7e-01 |
| **ENSG00000230695.2** | 564 | 1.7 | 2.6e-05 | 35,010 | -0.1 | 9.1e-01 |

| | Anti-PM/Scl vs. all | | | Anti-Mi2 vs. all | | |
|---|---|---|---|---|---|---|
| **Gene** | **Pos** | **logFC** | **adj.P.Val** | **Pos** | **logFC** | **adj.P.Val** |
| **ENSG00000280035.1** | 635 | 1.7 | 3.9e-05 | 22,528 | -0.3 | 4.8e-01 |
| **ENSG00000288746.1** | 647 | 1.4 | 4.2e-05 | 24,187 | -0.3 | 5.5e-01 |
| **EGFL7** | 662 | 0.8 | 4.5e-05 | 35,119 | 0.1 | 9.1e-01 |
| **ENSG00000274251.1** | 663 | 1.7 | 4.5e-05 | 25,787 | -0.2 | 6.2e-01 |
| **ENSG00000279140.1** | 667 | 1.4 | 4.6e-05 | 7,790 | -0.9 | 1.8e-02 |
| **ENSG00000278932.5** | 676 | 1.2 | 4.8e-05 | 1,648 | -1.4 | 3.5e-05 |
| **ENSG00000277020.4** | 710 | 1.4 | 5.5e-05 | 5,238 | -1.0 | 3.6e-03 |
| **ENSG00000274737.1** | 719 | 1.6 | 5.7e-05 | 17,000 | -0.5 | 2.4e-01 |
| **ENSG00000270012.1** | 720 | 1.0 | 5.8e-05 | 3,427 | -0.9 | 7.2e-04 |
| **RN7SL521P** | 723 | 1.6 | 6.0e-05 | 12,768 | -0.6 | 1.0e-01 |
| **ENSG00000288927.1** | 728 | 1.6 | 6.3e-05 | 17,550 | -0.5 | 2.6e-01 |
| **ENSG00000289543.1** | 736 | 1.4 | 6.5e-05 | 30,214 | -0.1 | 7.8e-01 |
| **ENSG00000289253.1** | 756 | 1.6 | 7.1e-05 | 20,414 | -0.4 | 3.8e-01 |
| **CPNE2-DT** | 761 | 1.4 | 7.5e-05 | 32,728 | -0.1 | 8.5e-01 |
| **ENSG00000264666.2** | 769 | 1.3 | 7.7e-05 | 22,291 | -0.3 | 4.6e-01 |
| **ENSG00000289119.1** | 771 | 1.4 | 7.7e-05 | 5,887 | -1.0 | 5.9e-03 |
| **ENSG00000289067.1** | 778 | 1.4 | 8.1e-05 | 4,128 | -1.1 | 1.5e-03 |
| **ENSG00000288942.1** | 810 | 1.5 | 9.6e-05 | 13,957 | -0.6 | 1.3e-01 |
| **DYNLL2-DT** | 813 | 1.7 | 9.8e-05 | 21,871 | -0.4 | 4.5e-01 |
| **MIR3188** | 817 | 1.5 | 9.8e-05 | 22,922 | -0.3 | 4.9e-01 |
| **ENSG00000289267.1** | 822 | 1.8 | 1.0e-04 | 40,240 | -0.1 | 9.7e-01 |
| **ENSG00000274092.1** | 845 | 1.6 | 1.1e-04 | 26,818 | -0.2 | 6.6e-01 |
| **ENSG00000267882.2** | 958 | 1.6 | 1.7e-04 | 31,209 | -0.2 | 8.1e-01 |
| **SMG1-DT** | 973 | 1.5 | 1.7e-04 | 12,940 | -0.6 | 1.1e-01 |
| **MAP3K11** | 1,001 | 0.5 | 1.9e-04 | 22,995 | -0.1 | 5.0e-01 |
| **ENSG00000289241.1** | 1,005 | 1.4 | 1.9e-04 | 40,799 | 0.0 | 9.7e-01 |
| **ZFX-AS1** | 1,016 | 1.4 | 2.0e-04 | 35,026 | -0.1 | 9.1e-01 |
| **ENSG00000272735.1** | 1,025 | 1.4 | 2.1e-04 | 12,399 | -0.7 | 9.4e-02 |
| **ENSG00000273335.1** | 1,026 | 1.5 | 2.1e-04 | 25,461 | 0.3 | 6.1e-01 |
| **ENSG00000287420.1** | 1,037 | 1.3 | 2.2e-04 | 39,718 | 0.0 | 9.7e-01 |
| **CETP** | 1,061 | 1.4 | 2.3e-04 | 27,248 | -0.3 | 6.8e-01 |
| **ENSG00000279198.1** | 1,104 | 1.2 | 2.5e-04 | 25,432 | -0.3 | 6.1e-01 |
| **ENSG00000261519.3** | 1,114 | 1.5 | 2.6e-04 | 24,261 | -0.3 | 5.6e-01 |
| **ENSG00000288772.1** | 1,122 | 1.5 | 2.7e-04 | 5,381 | -0.9 | 4.1e-03 |
| **ARHGEF2-AS2** | 1,144 | 1.2 | 2.9e-04 | 11,463 | -0.7 | 7.2e-02 |
| **ENSG00000288737.1** | 1,145 | 1.5 | 2.9e-04 | 17,756 | -0.4 | 2.7e-01 |
| **ENSG00000267212.1** | 1,178 | 1.3 | 3.3e-04 | 44,531 | 0.0 | 9.9e-01 |
| **ENSG00000279691.1** | 1,210 | 1.5 | 3.5e-04 | 27,034 | -0.2 | 6.7e-01 |
| **ENSG00000289330.1** | 1,223 | 1.5 | 3.6e-04 | 30,618 | -0.2 | 8.0e-01 |
| **ENSG00000289222.1** | 1,235 | 1.4 | 3.7e-04 | 35,135 | -0.1 | 9.1e-01 |
| **OIT3** | 1,249 | 1.7 | 3.8e-04 | 20,562 | -0.5 | 3.9e-01 |
| **ENSG00000273141.1** | 1,257 | 1.4 | 3.9e-04 | 678 | -2.0 | 9.6e-07 |
| **ENSG00000272195.1** | 1,320 | 1.0 | 4.5e-04 | 719 | -1.5 | 1.2e-06 |
| **LINC00115** | 1,321 | 0.9 | 4.6e-04 | 2,002 | -1.0 | 7.2e-05 |
| **ENSG00000278546.1** | 1,327 | 1.5 | 4.6e-04 | 24,328 | -0.3 | 5.6e-01 |
| **C10ORF95** | 1,377 | 1.4 | 5.3e-04 | 24,139 | -0.3 | 5.5e-01 |
| **ENSG00000289518.1** | 1,392 | 1.3 | 5.6e-04 | 36,356 | -0.1 | 9.3e-01 |
| **HEXA-AS1** | 1,424 | 1.4 | 6.1e-04 | 38,433 | -0.1 | 9.6e-01 |
| **HIGD2B** | 1,461 | 1.3 | 6.5e-04 | 30,695 | 0.2 | 8.0e-01 |
| **ENSG00000273248.1** | 1,462 | 1.5 | 6.6e-04 | 8,365 | -0.8 | 2.3e-02 |
| **ITPKB** | 1,507 | 0.4 | 7.4e-04 | 33,932 | -0.0 | 8.9e-01 |
| **ENSG00000262412.1** | 1,554 | 1.3 | 8.1e-04 | 17,569 | -0.4 | 2.6e-01 |
| **ENSG00000240790.2** | 1,763 | 1.3 | 1.2e-03 | 30,806 | -0.2 | 8.0e-01 |
| **ENSG00000289334.1** | 2,102 | 1.1 | 2.1e-03 | 8,873 | -0.8 | 2.9e-02 |
| **MIA2-AS1** | 2,224 | 1.2 | 2.4e-03 | 28,956 | -0.2 | 7.4e-01 |
| **OTULIN-DT** | 2,300 | 1.0 | 2.6e-03 | 8,470 | -0.7 | 2.4e-02 |
| **ENSG00000272969.1** | 2,525 | 1.3 | 3.5e-03 | 32,644 | 0.1 | 8.5e-01 |

| | Anti-PM/Scl vs. all | | | Anti-Mi2 vs. all | | |
|---|---|---|---|---|---|---|
| **Gene** | **Pos** | **logFC** | **adj.P.Val** | **Pos** | **logFC** | **adj.P.Val** |
| **ENSG00000277383.1** | 2,662 | 1.1 | 4.1e-03 | 3,356 | -1.3 | 6.6e-04 |
| **ENSG00000272906.1** | 2,789 | 1.0 | 4.8e-03 | 12,652 | -0.6 | 1.0e-01 |
| **ENSG00000283341.3** | 2,820 | 0.8 | 4.9e-03 | 2,689 | -1.1 | 2.6e-04 |
| **ENSG00000273289.1** | 2,852 | 1.1 | 5.1e-03 | 10,618 | -0.7 | 5.5e-02 |
| **ENSG00000279529.1** | 3,039 | 0.8 | 6.2e-03 | 2,288 | -1.0 | 1.3e-04 |
| **EXOC3L2** | 3,333 | 0.9 | 7.8e-03 | 6,175 | -0.9 | 7.3e-03 |
| **ENSG00000289065.1** | 3,437 | 0.8 | 8.5e-03 | 1,028 | -1.4 | 5.5e-06 |
| **ZNF252P-AS1** | 4,460 | 0.9 | 1.7e-02 | 17,388 | -0.5 | 2.6e-01 |
| **TNFRSF4** | 5,061 | 0.9 | 2.4e-02 | 22,220 | -0.4 | 4.6e-01 |
| **ENSG00000288061.2** | 5,690 | 0.7 | 3.2e-02 | 1,745 | -1.2 | 4.2e-05 |
| **ENSG00000277182.1** | 5,838 | 0.8 | 3.4e-02 | 30,769 | -0.2 | 8.0e-01 |
| **MHENCR** | 15,746 | 0.3 | 2.8e-01 | 4,107 | -0.6 | 1.5e-03 |
| **ENSG00000280152.1** | 16,487 | 0.5 | 3.0e-01 | 718 | -1.7 | 1.1e-06 |
| **ENSG00000276570.1** | 39,493 | 0.1 | 8.9e-01 | 3,518 | -1.0 | 8.0e-04 |

**Supplementary Table 3. Differential expression of anti-PM/Scl-specific genes (PMID 38902010) in muscle biopsies from anti-PM/Scl-positive patients, relative to all other samples, in an independent external validation cohort.** Pos, position in the complete differential expression table; logFC, $log_2$ fold change; adj.P.Val, Benjamini-Hochberg-adjusted P value.

| Gene | Pos | logFC | adj.P.Val |
|---|---|---|---|
| ENSG00000265100 | 1 | 2.9 | 1.8e-10 |
| ENSG00000268403 | 6 | 2.6 | 1.1e-08 |
| ENSG00000289296 | 7 | 2.4 | 1.2e-08 |
| ENSG00000260369 | 8 | 3.4 | 1.2e-08 |
| ENSG00000272735 | 13 | 2.7 | 2.2e-08 |
| ENSG00000239705 | 17 | 4.0 | 2.5e-08 |
| ENSG00000289499 | 20 | 2.3 | 3.8e-08 |
| ENSG00000289637 | 22 | 3.3 | 3.8e-08 |
| ENSG00000287821 | 29 | 3.6 | 4.0e-08 |
| CAPN10-DT | 30 | 1.6 | 5.8e-08 |
| BMP2K-DT | 33 | 3.2 | 6.8e-08 |
| ENSG00000289030 | 37 | 2.4 | 9.6e-08 |
| ENSG00000288804 | 41 | 3.2 | 1.9e-07 |
| ENSG00000279212 | 52 | 3.0 | 2.8e-07 |
| ENSG00000287584 | 53 | 1.9 | 3.1e-07 |
| ENSG00000286408 | 59 | 2.5 | 3.5e-07 |
| ENSG00000273210 | 60 | 2.7 | 3.5e-07 |
| MIR5188 | 61 | 4.2 | 3.5e-07 |
| ENSG00000287547 | 71 | 3.3 | 4.7e-07 |
| TTC32-DT | 84 | 2.9 | 7.4e-07 |
| DDX39B-AS1 | 100 | 2.8 | 1.0e-06 |
| HIGD2B | 105 | 3.5 | 1.1e-06 |
| TRIM51BP | 108 | 4.7 | 1.1e-06 |
| ENSG00000289119 | 116 | 2.5 | 1.3e-06 |
| ENSG00000288996 | 121 | 3.2 | 1.5e-06 |
| ENSG00000289115 | 122 | 3.4 | 1.5e-06 |
| ENSG00000288900 | 147 | 3.0 | 2.3e-06 |
| ENSG00000286409 | 149 | 2.7 | 2.4e-06 |
| ENSG00000273141 | 158 | 2.3 | 2.8e-06 |
| ENSG00000289253 | 159 | 2.1 | 2.8e-06 |
| TIMMDC1-DT | 169 | 2.9 | 3.5e-06 |
| ENSG00000261242 | 171 | 2.5 | 3.7e-06 |
| ENSG00000273338 | 182 | 4.6 | 5.0e-06 |
| ENSG00000272967 | 186 | 2.9 | 5.3e-06 |
| ENSG00000289303 | 187 | 1.7 | 5.3e-06 |
| ENSG00000255647 | 205 | 2.0 | 7.5e-06 |
| ENSG00000272768 | 223 | 2.1 | 9.8e-06 |
| ENSG00000269399 | 244 | 1.8 | 1.3e-05 |
| ENSG00000276524 | 256 | 2.0 | 1.5e-05 |
| ENSG00000240790 | 258 | 2.3 | 1.5e-05 |
| ENSG00000278017 | 267 | 1.8 | 1.7e-05 |
| ENSG00000284484 | 283 | 4.2 | 2.0e-05 |
| ENSG00000286113 | 296 | 3.0 | 2.2e-05 |
| ENSG00000274292 | 297 | 1.6 | 2.2e-05 |
| ENSG00000234432 | 300 | 1.6 | 2.4e-05 |
| AFF4-DT | 301 | 2.3 | 2.4e-05 |
| ENSG00000288872 | 316 | 3.0 | 3.0e-05 |
| ENSG00000288865 | 322 | 2.5 | 3.2e-05 |
| ENSG00000288751 | 323 | 1.7 | 3.2e-05 |
| THOC1-DT | 324 | 1.4 | 3.2e-05 |
| UAP1-DT | 331 | 2.0 | 3.3e-05 |

| Gene | Pos | logFC | adj.P.Val |
|---|---|---|---|
| **SLC38A2-AS1** | 349 | 2.5 | 3.8e-05 |
| **ENSG00000288988** | 357 | 1.8 | 3.9e-05 |
| **ENSG00000277383** | 364 | 1.8 | 4.2e-05 |
| **ENSG00000289229** | 369 | 2.5 | 4.4e-05 |
| **ENSG00000286444** | 384 | 3.0 | 5.1e-05 |
| **LINC02776** | 388 | 2.7 | 5.4e-05 |
| **ENSG00000272906** | 397 | 1.6 | 6.3e-05 |
| **ENSG00000287070** | 404 | 1.6 | 6.6e-05 |
| **ENSG00000287697** | 423 | 1.3 | 9.2e-05 |
| **ENSG00000289412** | 435 | 2.1 | 1.1e-04 |
| **ENSG00000274213** | 444 | 2.4 | 1.1e-04 |
| **ENSG00000273335** | 454 | 2.7 | 1.3e-04 |
| **ENSG00000289230** | 457 | 2.4 | 1.3e-04 |
| **MIR378D2HG** | 458 | 2.3 | 1.3e-04 |
| **PRAMEF13** | 465 | 2.4 | 1.4e-04 |
| **ENSG00000289341** | 474 | 2.9 | 1.4e-04 |
| **LINC01424** | 485 | 1.6 | 1.7e-04 |
| **ENSG00000286881** | 489 | 2.2 | 1.7e-04 |
| **ENSG00000288744** | 490 | 1.2 | 1.7e-04 |
| **ENSG00000289200** | 501 | 3.0 | 1.9e-04 |
| **ENSG00000289202** | 536 | 2.8 | 2.5e-04 |
| **ENSG00000272195** | 546 | 1.4 | 2.7e-04 |
| **RN7SL521P** | 549 | 2.1 | 2.8e-04 |
| **LINC00677** | 550 | 2.3 | 2.9e-04 |
| **ENSG00000245651** | 556 | 2.0 | 3.0e-04 |
| **ENSG00000225945** | 558 | 1.9 | 3.0e-04 |
| **CPEB2-DT** | 570 | 1.5 | 3.2e-04 |
| **ARHGEF2-AS2** | 571 | 1.3 | 3.3e-04 |
| **ENSG00000289031** | 622 | 1.1 | 4.7e-04 |
| **SMG1-DT** | 624 | 1.4 | 4.8e-04 |
| **ENSG00000288879** | 632 | 2.7 | 5.0e-04 |
| **ENSG00000289317** | 645 | 2.1 | 5.3e-04 |
| **ENSG00000272426** | 658 | 2.0 | 5.7e-04 |
| **ENSG00000289379** | 664 | 2.0 | 5.9e-04 |
| **ENSG00000289152** | 687 | 2.1 | 7.5e-04 |
| **ENSG00000278743** | 692 | 1.4 | 7.9e-04 |
| **ENSG00000287979** | 702 | 2.8 | 8.5e-04 |
| **ENSG00000279140** | 717 | 1.4 | 9.5e-04 |
| **ENSG00000272969** | 729 | 2.3 | 1.0e-03 |
| **DYNLL2-DT** | 734 | 2.3 | 1.0e-03 |
| **ENSG00000287654** | 746 | 2.1 | 1.1e-03 |
| **ENSG00000289334** | 759 | 1.7 | 1.2e-03 |
| **ENSG00000275709** | 763 | 1.7 | 1.3e-03 |
| **ENSG00000276744** | 764 | 1.6 | 1.3e-03 |
| **ENSG00000289301** | 768 | 2.4 | 1.4e-03 |
| **ENSG00000289055** | 772 | 1.8 | 1.4e-03 |
| **ENSG00000264666** | 790 | 1.7 | 1.6e-03 |
| **ENSG00000289257** | 810 | 2.2 | 1.9e-03 |
| **ENSG00000276216** | 816 | 2.1 | 1.9e-03 |
| **ENSG00000289221** | 822 | 2.6 | 2.0e-03 |
| **ENSG00000261334** | 829 | 1.8 | 2.1e-03 |
| **ENSG00000289065** | 831 | 1.6 | 2.2e-03 |
| **ENSG00000277020** | 842 | 1.6 | 2.3e-03 |
| **ZFX-AS1** | 870 | 2.1 | 2.6e-03 |
| **ENSG00000267882** | 911 | 1.7 | 2.9e-03 |
| **ENSG00000288746** | 935 | 1.3 | 3.4e-03 |
| **ENSG00000288919** | 948 | 1.2 | 3.6e-03 |
| **LINC01126** | 979 | 0.8 | 4.1e-03 |

| Gene | Pos | logFC | adj.P.Val |
|---|---|---|---|
| **ENSG00000289154** | 1,006 | 0.8 | 4.7e-03 |
| **ENSG00000289267** | 1,022 | 2.5 | 5.0e-03 |
| **ENSG00000286577** | 1,025 | 1.6 | 5.1e-03 |
| **ENSG00000289288** | 1,047 | 1.7 | 5.8e-03 |
| **ENSG00000288835** | 1,067 | 1.6 | 6.3e-03 |
| **ENSG00000288955** | 1,086 | 1.8 | 6.9e-03 |
| **ENSG00000260651** | 1,113 | 1.9 | 8.1e-03 |
| **ENSG00000289457** | 1,120 | 1.0 | 8.2e-03 |
| **SMG7-AS1** | 1,139 | 1.3 | 9.2e-03 |
| **CPNE2-DT** | 1,150 | 2.0 | 9.8e-03 |
| **ENSG00000286482** | 1,153 | 0.8 | 9.9e-03 |
| **ENSG00000289550** | 1,157 | 2.6 | 1.0e-02 |
| **TRIM8-DT** | 1,198 | 1.3 | 1.2e-02 |
| **ENSG00000288942** | 1,255 | 0.9 | 1.5e-02 |
| **MSRA-DT** | 1,267 | 1.0 | 1.6e-02 |
| **ENSG00000268670** | 1,271 | 0.9 | 1.6e-02 |
| **ENSG00000224505** | 1,310 | 0.7 | 1.8e-02 |
| **ENSG00000288927** | 1,347 | 1.0 | 2.1e-02 |
| **CAGE1** | 1,350 | 2.1 | 2.1e-02 |
| **ENSG00000274751** | 1,392 | 1.4 | 2.5e-02 |
| **HEXA-AS1** | 1,430 | 1.2 | 2.8e-02 |
| **ENSG00000279259** | 1,432 | 1.8 | 2.9e-02 |
| **ENSG00000289222** | 1,444 | 2.0 | 3.0e-02 |
| **ENSG00000289551** | 1,463 | 0.8 | 3.0e-02 |
| **MIA2-AS1** | 1,469 | 1.2 | 3.1e-02 |
| **ENSG00000288842** | 1,529 | 1.6 | 3.6e-02 |
| **ENSG00000273064** | 1,542 | 0.9 | 3.7e-02 |
| **ENSG00000289506** | 1,564 | 1.2 | 4.0e-02 |
| **ENSG00000289103** | 1,610 | 0.7 | 4.4e-02 |
| **ENSG00000241666** | 1,621 | 1.0 | 4.5e-02 |
| **ATXN7L3-AS1** | 1,652 | 0.6 | 4.9e-02 |
| **OTULIN-DT** | 1,678 | 1.0 | 5.2e-02 |
| **SDR42E2** | 1,681 | 1.3 | 5.2e-02 |
| **ENSG00000259135** | 1,687 | 1.3 | 5.3e-02 |
| **ENSG00000278002** | 1,716 | 0.8 | 5.6e-02 |
| **ENSG00000264739** | 1,844 | 1.4 | 7.4e-02 |
| **CLMAT3** | 1,864 | 1.5 | 7.6e-02 |
| **ENSG00000289182** | 1,873 | 0.9 | 7.8e-02 |
| **ENSG00000279491** | 1,929 | 1.2 | 8.8e-02 |
| **KLF2-DT** | 1,940 | 0.9 | 8.9e-02 |
| **ENSG00000288061** | 1,986 | 0.7 | 9.4e-02 |
| **MIR23AHG** | 2,142 | 0.9 | 1.1e-01 |
| **ENSG00000279529** | 2,154 | 0.4 | 1.1e-01 |
| **ENSG00000289067** | 2,206 | 0.9 | 1.2e-01 |
| **ITPKB** | 2,216 | -0.5 | 1.2e-01 |
| **ENSG00000288939** | 2,232 | 1.2 | 1.2e-01 |
| **ENSG00000270012** | 2,236 | 0.5 | 1.2e-01 |
| **ENSG00000289543** | 2,363 | 1.7 | 1.4e-01 |
| **ENSG00000257258** | 2,575 | 1.5 | 1.7e-01 |
| **RENO1** | 2,655 | 0.4 | 1.9e-01 |
| **ANKH-DT** | 2,705 | 1.4 | 1.9e-01 |
| **ENSG00000267212** | 2,771 | 0.8 | 2.0e-01 |
| **MHENCR** | 2,805 | 0.6 | 2.0e-01 |
| **ENSG00000279691** | 3,011 | 0.7 | 2.3e-01 |
| **LINC01089** | 3,242 | 0.5 | 2.6e-01 |
| **TERC** | 3,387 | 0.6 | 2.7e-01 |
| **ENSG00000289330** | 3,641 | 1.3 | 3.0e-01 |
| **ENSG00000277182** | 3,893 | 0.8 | 3.2e-01 |

| Gene | Pos | logFC | adj.P.Val |
|---|---|---|---|
| **ENSG00000283959** | 3,907 | 0.8 | 3.2e-01 |
| **ENSG00000289059** | 3,988 | 0.7 | 3.3e-01 |
| **ENSG00000262412** | 4,355 | 0.7 | 3.6e-01 |
| **ENSG00000289547** | 4,514 | 0.8 | 3.7e-01 |
| **SPACA6P-AS** | 4,803 | 0.9 | 3.9e-01 |
| **COL18A1** | 5,251 | 0.6 | 4.2e-01 |
| **ENSG00000288813** | 5,262 | 1.3 | 4.2e-01 |
| **EGFL7** | 5,757 | 0.4 | 4.5e-01 |
| **ENSG00000289159** | 5,878 | 1.1 | 4.6e-01 |
| **ZNF252P-AS1** | 6,237 | 0.4 | 4.8e-01 |
| **ENSG00000288929** | 7,175 | 0.6 | 5.2e-01 |
| **ENSG00000230695** | 7,235 | 1.0 | 5.3e-01 |
| **EXOC3L2** | 7,589 | 0.7 | 5.4e-01 |
| **ENSG00000274251** | 9,262 | 0.3 | 6.1e-01 |
| **MIRLET7BHG** | 9,396 | 0.2 | 6.1e-01 |
| **ENSG00000288866** | 9,854 | 0.4 | 6.3e-01 |
| **LINC00896** | 9,985 | 0.5 | 6.3e-01 |
| **ENSG00000289177** | 10,001 | 0.2 | 6.3e-01 |
| **ENSG00000280152** | 10,935 | 0.2 | 6.6e-01 |
| **ENSG00000278932** | 11,098 | 0.4 | 6.6e-01 |
| **TNFRSF4** | 15,102 | 0.5 | 7.5e-01 |
| **TNFAIP8L1** | 17,598 | 0.2 | 8.0e-01 |
| **ENSG00000278546** | 18,146 | 0.4 | 8.1e-01 |
| **ENSG00000289235** | 19,171 | 0.4 | 8.2e-01 |
| **OIT3** | 19,402 | 0.5 | 8.3e-01 |
| **ENSG00000274737** | 20,140 | 0.2 | 8.4e-01 |
| **MIR1915HG** | 21,234 | 0.3 | 8.6e-01 |
| **ENSG00000287420** | 21,675 | 0.4 | 8.6e-01 |
| **FSCN1** | 22,834 | 0.3 | 8.8e-01 |
| **ENSG00000274092** | 23,199 | 0.3 | 8.8e-01 |
| **PRND** | 23,378 | 0.7 | 8.9e-01 |
| **ENSG00000282936** | 25,015 | 0.2 | 9.1e-01 |
| **ENSG00000289044** | 25,415 | 0.1 | 9.1e-01 |
| **ENSG00000280035** | 26,813 | -0.1 | 9.3e-01 |
| **MAP3K11** | 26,947 | 0.1 | 9.3e-01 |
| **ENSG00000289518** | 26,948 | -0.2 | 9.3e-01 |
| **ENSG00000273248** | 27,246 | 0.1 | 9.3e-01 |
| **ENSG00000276570** | 28,986 | -0.1 | 9.5e-01 |
| **ENSG00000253838** | 29,287 | -0.2 | 9.5e-01 |
| **LINC00115** | 33,981 | 0.0 | 1.0e+00 |
| **RABEP2** | 34,012 | -0.0 | 1.0e+00 |
| **CETP** | 34,071 | -0.0 | 1.0e+00 |

**Supplementary Table 4. Association between a previously defined set of anti-Mi2- and anti-PM/Scl-specific genes (PMID 38902010) and genes differentially expressed (q-value < 0.001) at 24h and 72h following electroporation of purified immunoglobulins into primary muscle cell cultures, relative to other electroporated samples.** Anti-Mi2-specific genes are preferentially enriched at 24h post-electroporation, whereas anti-PM/Scl-specific genes are preferentially enriched at 72h post-electroporation. IBM, inclusion body myositis; ACA, anti-centromere autoantibodies.

### 24h after electroporation

| Group | Mi2 | Not Mi2 | p-value |
|---|---|---|---|
| Control | 0% (0) | 0% (0) | 1 |
| Mi2 | 66% (75) | 2% (1196) | <2e-16 |
| PM/Scl | 1% (1) | 2% (937) | 1 |
| NXP2 | 0% (0) | 0% (0) | 1 |
| TIF1 | 0% (0) | 0% (2) | 1 |
| MDA5 | 0% (0) | 0% (46) | 1 |
| Jo1 | 0% (0) | 0% (0) | 1 |
| HMGCR | 0% (0) | 0% (0) | 1 |
| SRP | 0% (0) | 0% (3) | 1 |
| IBM | 0% (0) | 0% (2) | 1 |
| Ku | 0% (0) | 0% (10) | 1 |
| Scl70 | 1% (1) | 1% (572) | 1 |
| ACA | 0% (0) | 0% (11) | 1 |

| Group | PM/Scl | Not PM/Scl | p-value |
|---|---|---|---|
| Control | 0% (0) | 0% (0) | 1 |
| Mi2 | 1% (2) | 2% (1269) | 0.25 |
| PM/Scl | 69% (163) | 1% (775) | <2e-16 |
| NXP2 | 0% (0) | 0% (0) | 1 |
| TIF1 | 0% (0) | 0% (2) | 1 |
| MDA5 | 0% (0) | 0% (46) | 1 |
| Jo1 | 0% (0) | 0% (0) | 1 |
| HMGCR | 0% (0) | 0% (0) | 1 |
| SRP | 0% (0) | 0% (3) | 1 |
| IBM | 0% (0) | 0% (2) | 1 |
| Ku | 0% (0) | 0% (10) | 1 |
| Scl70 | 0% (0) | 1% (573) | 0.18 |
| ACA | 0% (0) | 0% (11) | 1 |

### 72h after electroporation

| Group | Mi2 | Not Mi2 | p-value |
|---|---|---|---|
| Control | 0% (0) | 0% (0) | 1 |
| Mi2 | 49% (55) | 1% (435) | <2e-16 |
| PM/Scl | 3% (3) | 3% (1903) | 1 |
| NXP2 | 0% (0) | 0% (0) | 1 |
| TIF1 | 0% (0) | 0% (12) | 1 |
| MDA5 | 0% (0) | 0% (0) | 1 |
| Jo1 | 0% (0) | 0% (0) | 1 |
| HMGCR | 0% (0) | 0% (1) | 1 |
| SRP | 0% (0) | 0% (0) | 1 |
| IBM | 0% (0) | 0% (0) | 1 |
| Ku | 0% (0) | 0% (3) | 1 |
| Scl70 | 0% (0) | 0% (87) | 1 |
| ACA | 0% (0) | 0% (63) | 1 |

| Group | PM/Scl | Not PM/Scl | p-value |
|---|---|---|---|
| Control | 0% (0) | 0% (0) | 1 |
| Mi2 | 2% (4) | 1% (486) | 0.12 |
| PM/Scl | 86% (202) | 3% (1704) | <2e-16 |
| NXP2 | 0% (0) | 0% (0) | 1 |
| TIF1 | 0% (0) | 0% (12) | 1 |
| MDA5 | 0% (0) | 0% (0) | 1 |
| Jo1 | 0% (0) | 0% (0) | 1 |
| HMGCR | 0% (0) | 0% (1) | 1 |
| SRP | 0% (0) | 0% (0) | 1 |
| IBM | 0% (0) | 0% (0) | 1 |
| Ku | 0% (0) | 0% (3) | 1 |
| Scl70 | 0% (0) | 0% (87) | 1 |
| ACA | 0% (0) | 0% (63) | 1 |

**Supplementary Table 5. Differential expression of anti-Mi2-specific genes (PMID 38902010) at 24h and 72h following electroporation of purified immunoglobulins from anti-Mi2-positive patients into primary muscle cell cultures, relative to other electroporated conditions.** Pos, position in the complete differential expression table; logFC, $\log_2$ fold change; adj.P.Val, Benjamini-Hochberg-adjusted P value.

| | Mi2-specific genes at 24h | | | Mi2-specific genes at 72h | | |
|---|---|---|---|---|---|---|
| **Gene** | **Pos** | **logFC** | **adj.P.Val** | **Pos** | **logFC** | **adj.P.Val** |
| **TEX19** | 2 | 4.5 | 3.6e-24 | 317 | 2.5 | 4.3e-05 |
| **TMEM151A** | 3 | 3.5 | 3.6e-24 | 278 | 1.6 | 1.4e-05 |
| **CAMKV** | 4 | 6.1 | 6.0e-23 | 324 | 2.6 | 5.5e-05 |
| **COX6B2** | 9 | 4.2 | 4.4e-22 | 32 | 3.3 | 1.9e-15 |
| **PLPPR3** | 10 | 3.4 | 8.8e-22 | 262 | 1.5 | 6.8e-06 |
| **PPP1R1B** | 12 | 5.8 | 1.7e-21 | 53 | 3.6 | 4.0e-13 |
| **BAIAP3** | 16 | 3.4 | 7.2e-21 | 70 | 1.8 | 1.4e-11 |
| **MSI1** | 17 | 3.0 | 7.2e-21 | 259 | 1.3 | 5.9e-06 |
| **TMEM151B** | 25 | 4.0 | 1.9e-19 | 207 | 2.6 | 5.0e-07 |
| **LINC00624** | 27 | 4.8 | 2.2e-19 | 55 | 3.5 | 6.7e-13 |
| **KIF1A** | 32 | 3.5 | 6.7e-19 | 54 | 2.4 | 4.1e-13 |
| **RAP1GAP** | 34 | 2.9 | 1.7e-18 | 1,688 | 0.7 | 1.5e-01 |
| **P2RX2** | 39 | 4.5 | 8.6e-18 | 108 | 3.6 | 5.9e-10 |
| **CACNA1G** | 43 | 3.0 | 1.8e-17 | 89 | 2.4 | 8.1e-11 |
| **JPH3** | 44 | 3.4 | 1.8e-17 | 200 | 2.5 | 3.2e-07 |
| **PALM3** | 47 | 3.7 | 2.2e-17 | 335 | 1.5 | 7.1e-05 |
| **CAPN12** | 55 | 3.0 | 5.6e-17 | 183 | 1.6 | 1.5e-07 |
| **DLGAP3** | 57 | 3.6 | 8.2e-17 | 161 | 2.5 | 4.2e-08 |
| **KLC3** | 62 | 2.9 | 1.3e-16 | 468 | 1.5 | 7.1e-04 |
| **LGI3** | 67 | 3.4 | 2.0e-16 | 43 | 3.1 | 2.1e-14 |
| **FLRT1** | 70 | 3.0 | 3.0e-16 | 4,037 | 0.9 | 4.4e-01 |
| **BANCR** | 74 | 4.1 | 8.9e-16 | 123 | 2.7 | 2.5e-09 |
| **ANKRD20A19P** | 80 | 3.7 | 1.8e-15 | 99 | 3.4 | 2.2e-10 |
| **IGLON5** | 82 | 3.6 | 2.4e-15 | 62 | 3.5 | 2.7e-12 |
| **CAMSAP3** | 86 | 3.4 | 4.7e-15 | 189 | 2.6 | 2.2e-07 |
| **ABCG4** | 95 | 2.9 | 1.1e-14 | 598 | 1.1 | 3.7e-03 |
| **TMEM145** | 96 | 2.6 | 1.1e-14 | 218 | 1.7 | 8.0e-07 |
| **CADM4** | 97 | 1.8 | 1.3e-14 | 143 | 1.4 | 1.3e-08 |
| **CHGA** | 108 | 3.9 | 7.2e-14 | 128 | 2.8 | 4.0e-09 |
| **COL2A1** | 109 | 2.9 | 9.2e-14 | 211 | 1.9 | 6.1e-07 |
| **SPIB** | 120 | 3.0 | 1.9e-13 | 164 | 2.1 | 4.6e-08 |
| **EFNA3** | 126 | 1.9 | 2.9e-13 | 567 | 0.9 | 2.6e-03 |
| **UTF1** | 127 | 4.0 | 3.8e-13 | 273 | 2.5 | 1.1e-05 |
| **CLDN6** | 138 | 2.4 | 7.8e-13 | 254 | 1.4 | 3.7e-06 |
| **SLC29A4** | 156 | 2.0 | 3.5e-12 | 432 | 0.9 | 4.9e-04 |
| **PRR35** | 173 | 3.5 | 7.3e-12 | 472 | 1.7 | 7.7e-04 |
| **HPCA** | 179 | 2.6 | 9.3e-12 | 210 | 1.9 | 6.0e-07 |
| **CASKIN1** | 180 | 1.7 | 1.0e-11 | 371 | 0.8 | 1.8e-04 |
| **YBX2** | 181 | 2.6 | 1.1e-11 | 599 | 1.3 | 3.8e-03 |
| **NKAIN4** | 187 | 3.0 | 1.2e-11 | 479 | 1.8 | 9.2e-04 |
| **DACT3** | 200 | 2.1 | 2.8e-11 | 167 | 1.1 | 5.4e-08 |
| **KCNJ4** | 201 | 2.6 | 2.9e-11 | 229 | 1.8 | 1.8e-06 |
| **SHISA7** | 229 | 4.3 | 1.1e-10 | 180 | 3.3 | 1.1e-07 |
| **SEMA4G** | 237 | 1.8 | 1.7e-10 | 808 | 0.4 | 2.0e-02 |
| **WFDC2** | 239 | 3.7 | 1.9e-10 | 50 | 4.2 | 3.3e-13 |
| **CTSV** | 257 | 1.4 | 4.7e-10 | 348 | 1.2 | 8.5e-05 |
| **CPNE6** | 258 | 3.3 | 4.9e-10 | 45 | 2.7 | 8.4e-14 |
| **CHRM4** | 273 | 2.2 | 8.0e-10 | 893 | 1.3 | 2.9e-02 |
| **ECE2** | 284 | 4.9 | 1.4e-09 | 119 | 4.1 | 1.7e-09 |
| **ZNF467** | 290 | 3.0 | 1.6e-09 | 746 | 1.8 | 1.2e-02 |

| | Mi2-specific genes at 24h | | | Mi2-specific genes at 72h | | |
|---|---|---|---|---|---|---|
| **Gene** | **Pos** | **logFC** | **adj.P.Val** | **Pos** | **logFC** | **adj.P.Val** |
| **TH** | 299 | 2.3 | 2.1e-09 | 430 | 1.5 | 4.7e-04 |
| **OVOL1** | 371 | 2.9 | 1.4e-08 | 170 | 2.6 | 6.1e-08 |
| **B4GALNT4** | 388 | 1.9 | 2.3e-08 | 483 | 1.6 | 9.4e-04 |
| **SCRT1** | 456 | 2.4 | 1.3e-07 | 744 | 1.4 | 1.2e-02 |
| **GCGR** | 483 | 2.5 | 2.2e-07 | 391 | 2.4 | 2.3e-04 |
| **ENSG00000289332.1** | 511 | 2.7 | 4.0e-07 | 185 | 3.0 | 1.6e-07 |
| **ENSG00000169093.16** | 530 | 0.8 | 6.7e-07 | 454 | 0.6 | 6.3e-04 |
| **KCNQ2** | 539 | 2.8 | 7.5e-07 | 303 | 2.8 | 2.5e-05 |
| **STAC2** | 545 | 1.5 | 7.9e-07 | 2,934 | 0.6 | 3.2e-01 |
| **DHRS2** | 574 | 1.9 | 1.5e-06 | 413 | 2.1 | 3.6e-04 |
| **CRB3** | 605 | 2.7 | 2.7e-06 | 805 | 1.6 | 1.9e-02 |
| **FBXL16** | 662 | 1.7 | 6.8e-06 | 588 | 0.9 | 3.3e-03 |
| **CACNA1I** | 728 | 2.3 | 1.5e-05 | 761 | 1.7 | 1.4e-02 |
| **ENSG00000286311.1** | 732 | 1.5 | 1.6e-05 | 14,714 | 0.3 | 8.4e-01 |
| **SMIM24** | 783 | 1.8 | 2.6e-05 | 384 | 2.6 | 2.0e-04 |
| **FAM171A2** | 853 | 1.2 | 5.1e-05 | 6,716 | 0.5 | 6.2e-01 |
| **GRIN3B** | 964 | 1.6 | 1.4e-04 | 3,901 | 0.8 | 4.3e-01 |
| **RAB26** | 968 | 1.9 | 1.5e-04 | 310 | 1.1 | 3.3e-05 |
| **RAB3B** | 986 | 0.9 | 1.7e-04 | 17,260 | -0.1 | 8.9e-01 |
| **HCN2** | 1,025 | 0.6 | 2.2e-04 | 1,536 | 0.5 | 1.2e-01 |
| **ARHGDIG** | 1,036 | 1.4 | 2.5e-04 | 529 | 1.4 | 1.7e-03 |
| **ESPN** | 1,065 | 1.1 | 3.2e-04 | 3,736 | 0.7 | 4.1e-01 |
| **SOX15** | 1,175 | 1.1 | 6.3e-04 | 2,241 | 0.5 | 2.3e-01 |
| **TNNI3** | 1,176 | 1.6 | 6.3e-04 | 1,134 | 1.1 | 6.2e-02 |
| **IFITM5** | 1,209 | 2.4 | 7.4e-04 | 707 | 1.6 | 9.8e-03 |
| **PRKAR1B** | 1,284 | 0.7 | 1.1e-03 | 2,082 | 0.4 | 2.1e-01 |
| **GRIN2D** | 1,287 | 1.2 | 1.1e-03 | 6,809 | 0.5 | 6.3e-01 |
| **LKAAEAR1** | 1,291 | 2.1 | 1.1e-03 | 1,168 | 1.5 | 6.8e-02 |
| **IL11** | 1,424 | 1.3 | 2.3e-03 | 23,689 | -0.1 | 9.8e-01 |
| **KREMEN2** | 1,643 | 1.2 | 5.3e-03 | 11,034 | 0.5 | 7.6e-01 |
| **PDIA2** | 1,693 | 0.8 | 6.3e-03 | 398 | 1.8 | 2.6e-04 |
| **ENTPD8** | 1,736 | 1.9 | 7.1e-03 | 17,744 | 0.2 | 9.0e-01 |
| **ENSG00000267892.1** | 1,874 | 1.2 | 1.0e-02 | 1,853 | 0.6 | 1.8e-01 |
| **PRKCG** | 2,093 | 0.6 | 1.6e-02 | 778 | 1.0 | 1.6e-02 |
| **CBARP** | 2,747 | 0.5 | 4.2e-02 | 5,638 | 0.2 | 5.6e-01 |
| **AQP5** | 2,816 | 1.2 | 4.6e-02 | 650 | 1.1 | 5.9e-03 |
| **ENSG00000260293.2** | 2,842 | 1.0 | 4.7e-02 | 6,219 | 0.5 | 6.0e-01 |
| **SSU72P8** | 3,168 | 0.7 | 6.7e-02 | 81 | 2.7 | 3.1e-11 |
| **COL26A1** | 3,545 | 1.0 | 8.8e-02 | 2,092 | 1.3 | 2.1e-01 |
| **VWA5B2** | 5,107 | 0.9 | 2.1e-01 | 1,849 | 0.9 | 1.8e-01 |
| **MADCAM1** | 7,365 | 0.5 | 3.9e-01 | 571 | 1.3 | 2.6e-03 |
| **ENSG00000275437.1** | 8,813 | 0.3 | 4.8e-01 | 22,714 | 0.1 | 9.7e-01 |
| **GJD2** | 9,051 | -0.6 | 4.9e-01 | 509 | 1.3 | 1.3e-03 |
| **ENSG00000218416.4** | 9,411 | 0.5 | 5.1e-01 | 11,984 | 0.5 | 7.8e-01 |
| **KBTBD11-AS1** | 10,062 | 0.6 | 5.5e-01 | 17,934 | -0.3 | 9.0e-01 |
| **ALPG** | 20,050 | 0.2 | 9.0e-01 | 268 | 3.4 | 7.6e-06 |
| **ENSG00000223561.7** | 20,277 | 0.2 | 9.0e-01 | 6,002 | 0.7 | 5.8e-01 |
| **RAC3** | 23,126 | 0.0 | 9.6e-01 | 7,238 | 0.3 | 6.4e-01 |
| **CT69** | 23,950 | -0.1 | 9.7e-01 | 13,098 | 0.4 | 8.1e-01 |
| **FAM131C** | 24,144 | 0.0 | 9.8e-01 | 13,940 | -0.2 | 8.3e-01 |

**Supplementary Table 6. Differential expression of anti-PM/Scl-specific genes (PMID 38902010) at 72h and 24h following electroporation of purified immunoglobulins from anti-PM/Scl-positive patients into primary muscle cell cultures, relative to other electroporated conditions.** Pos, position in the complete differential expression table; logFC, $log_2$ fold change; adj.P.Val, Benjamini-Hochberg-adjusted P value.

| | PM/Scl-specific genes at 72h | | | PM/Scl-specific genes at 24h | | |
|---|---|---|---|---|---|---|
| **Gene** | **Pos** | **logFC** | **adj.P.Val** | **Pos** | **logFC** | **adj.P.Val** |
| **ENSG00000226380.9** | 2 | 5.0 | 3.4e-25 | 128 | 2.6 | 2.4e-07 |
| **ENSG00000288865.1** | 3 | 5.2 | 5.0e-21 | 549 | 2.8 | 7.9e-05 |
| **ENSG00000274292.1** | 6 | 4.7 | 8.1e-21 | 52 | 2.6 | 1.9e-08 |
| **ENSG00000288751.1** | 8 | 6.1 | 5.2e-20 | 22 | 4.2 | 2.0e-09 |
| **ENSG00000289543.1** | 9 | 6.3 | 7.9e-20 | 33 | 4.6 | 9.6e-09 |
| **MIRLET7BHG** | 11 | 2.3 | 2.1e-19 | 362 | 0.8 | 1.4e-05 |
| **ENSG00000272426.1** | 12 | 4.8 | 1.5e-18 | 176 | 3.4 | 7.7e-07 |
| **ENSG00000288955.1** | 14 | 5.9 | 2.5e-18 | 138 | 4.3 | 3.0e-07 |
| **ENSG00000288900.1** | 20 | 6.4 | 1.6e-17 | 62 | 4.4 | 2.5e-08 |
| **ENSG00000289412.1** | 21 | 3.1 | 4.0e-17 | 212 | 2.5 | 1.6e-06 |
| **ENSG00000234432.4** | 23 | 4.1 | 4.1e-17 | 26 | 3.3 | 5.5e-09 |
| **ENSG00000239705.2** | 24 | 6.3 | 4.6e-17 | 4 | 4.3 | 1.3e-10 |
| **ENSG00000287547.1** | 26 | 5.0 | 1.2e-16 | 35 | 3.5 | 1.1e-08 |
| **ENSG00000278017.1** | 28 | 5.1 | 1.4e-16 | 783 | 2.5 | 3.5e-04 |
| **ENSG00000289030.1** | 30 | 6.0 | 1.7e-16 | 17 | 4.7 | 1.0e-09 |
| **ENSG00000289059.1** | 36 | 4.4 | 3.3e-16 | 47 | 3.3 | 1.6e-08 |
| **ENSG00000276216.1** | 38 | 6.3 | 4.7e-16 | 32 | 4.2 | 9.6e-09 |
| **ENSG00000273338.1** | 39 | 4.6 | 5.2e-16 | 175 | 3.6 | 7.7e-07 |
| **ENSG00000230695.2** | 40 | 4.6 | 5.5e-16 | 100 | 3.5 | 8.5e-08 |
| **ENSG00000261334.1** | 41 | 6.4 | 5.8e-16 | 8 | 4.5 | 3.0e-10 |
| **ENSG00000273210.1** | 43 | 4.9 | 5.8e-16 | 13 | 3.5 | 7.5e-10 |
| **ENSG00000289257.1** | 44 | 4.6 | 7.0e-16 | 92 | 3.3 | 8.1e-08 |
| **ENSG00000289200.1** | 47 | 5.2 | 7.5e-16 | 434 | 3.1 | 2.3e-05 |
| **ENSG00000289044.1** | 48 | 5.2 | 8.2e-16 | 121 | 2.7 | 1.7e-07 |
| **ENSG00000289457.1** | 56 | 4.9 | 1.4e-15 | 658 | 3.0 | 1.6e-04 |
| **TTC32-DT** | 58 | 5.1 | 1.5e-15 | 126 | 3.4 | 2.2e-07 |
| **ENSG00000278002.1** | 60 | 3.1 | 1.6e-15 | 74 | 2.4 | 4.5e-08 |
| **ENSG00000272967.1** | 62 | 4.2 | 1.7e-15 | 604 | 2.5 | 1.1e-04 |
| **ATXN7L3-AS1** | 67 | 4.3 | 2.9e-15 | 929 | 2.4 | 9.1e-04 |
| **ENSG00000265100.1** | 74 | 4.9 | 6.1e-15 | 75 | 3.5 | 4.5e-08 |
| **ENSG00000287979.1** | 76 | 5.9 | 6.8e-15 | 990 | 2.5 | 1.3e-03 |
| **ENSG00000289288.1** | 78 | 3.8 | 7.0e-15 | 1,569 | 1.9 | 1.8e-02 |
| **ENSG00000288835.1** | 79 | 4.4 | 7.2e-15 | 10 | 3.8 | 5.3e-10 |
| **ENSG00000289550.1** | 80 | 3.2 | 9.0e-15 | 285 | 2.5 | 5.2e-06 |
| **ENSG00000289341.1** | 81 | 4.8 | 1.1e-14 | 715 | 2.5 | 2.4e-04 |
| **ENSG00000272735.1** | 82 | 5.4 | 1.2e-14 | 116 | 3.5 | 1.1e-07 |
| **ENSG00000279212.1** | 83 | 5.2 | 1.6e-14 | 258 | 3.7 | 4.2e-06 |
| **CLMAT3** | 85 | 4.7 | 2.2e-14 | 55 | 3.4 | 2.1e-08 |
| **ENSG00000254028.1** | 89 | 4.2 | 3.1e-14 | 535 | 2.4 | 7.0e-05 |
| **ENSG00000289103.1** | 93 | 4.7 | 3.4e-14 | 229 | 3.5 | 2.3e-06 |
| **ENSG00000286409.2** | 95 | 4.5 | 3.9e-14 | 27 | 3.7 | 6.3e-09 |
| **ENSG00000272195.1** | 96 | 3.9 | 4.1e-14 | 85 | 2.9 | 6.1e-08 |
| **ENSG00000260369.2** | 97 | 5.5 | 4.1e-14 | 34 | 4.3 | 1.0e-08 |
| **ENSG00000225945.1** | 98 | 4.3 | 4.6e-14 | 683 | 2.5 | 2.0e-04 |
| **AFF4-DT** | 102 | 4.5 | 5.8e-14 | 718 | 2.5 | 2.5e-04 |
| **ENSG00000287584.1** | 104 | 4.4 | 5.9e-14 | 251 | 3.3 | 3.5e-06 |
| **ENSG00000289221.1** | 108 | 4.5 | 7.3e-14 | 143 | 3.8 | 3.2e-07 |
| **HEXA-AS1** | 109 | 3.1 | 7.3e-14 | 2 | 3.2 | 1.0e-10 |
| **ENSG00000288929.1** | 111 | 4.8 | 7.4e-14 | 159 | 3.4 | 4.6e-07 |
| **ENSG00000274751.1** | 115 | 2.8 | 8.1e-14 | 820 | 1.6 | 4.5e-04 |
| **ENSG00000289267.1** | 116 | 4.5 | 8.3e-14 | 418 | 3.1 | 2.1e-05 |
| **ENSG00000279491.1** | 121 | 3.9 | 1.1e-13 | 179 | 2.9 | 8.2e-07 |
| **MIR378D2HG** | 122 | 5.0 | 1.2e-13 | 202 | 3.5 | 1.4e-06 |
| **LINC01424** | 124 | 3.7 | 1.4e-13 | 95 | 2.5 | 8.4e-08 |
| **ENSG00000289115.1** | 125 | 5.1 | 1.6e-13 | 480 | 2.9 | 4.1e-05 |
| **ENSG00000289119.1** | 127 | 2.7 | 1.7e-13 | 67 | 2.1 | 3.5e-08 |

| | PM/Scl-specific genes at 72h | | | PM/Scl-specific genes at 24h | | |
|---|---|---|---|---|---|---|
| **Gene** | **Pos** | **logFC** | **adj.P.Val** | **Pos** | **logFC** | **adj.P.Val** |
| **ENSG00000288942.1** | 128 | 3.4 | 1.7e-13 | 740 | 2.2 | 2.8e-04 |
| **ENSG00000224505.3** | 129 | 3.2 | 1.7e-13 | 97 | 2.6 | 8.5e-08 |
| **ENSG00000289330.1** | 130 | 3.7 | 1.7e-13 | 1,196 | 2.0 | 4.0e-03 |
| **ENSG00000289055.1** | 136 | 4.0 | 2.3e-13 | 1,035 | 2.2 | 1.6e-03 |
| **ENSG00000288943.1** | 138 | 5.2 | 2.3e-13 | 288 | 3.2 | 5.5e-06 |
| **ENSG00000277020.4** | 140 | 3.9 | 2.4e-13 | 336 | 2.9 | 1.0e-05 |
| **ENSG00000287697.1** | 145 | 3.9 | 2.6e-13 | 135 | 2.4 | 2.9e-07 |
| **ENSG00000286881.1** | 146 | 4.2 | 2.7e-13 | 149 | 3.1 | 3.7e-07 |
| **TRIM8-DT** | 147 | 3.1 | 3.4e-13 | 201 | 2.2 | 1.3e-06 |
| **ENSG00000274092.1** | 149 | 4.1 | 3.9e-13 | 29 | 3.3 | 7.4e-09 |
| **SLC38A2-AS1** | 151 | 4.5 | 4.1e-13 | 152 | 3.4 | 4.1e-07 |
| **SPACA6P-AS** | 153 | 3.2 | 5.4e-13 | 331 | 2.0 | 9.8e-06 |
| **BMP2K-DT** | 154 | 3.9 | 5.5e-13 | 61 | 2.9 | 2.5e-08 |
| **ENSG00000245651.3** | 157 | 4.8 | 6.3e-13 | 537 | 2.6 | 7.3e-05 |
| **ENSG00000286444.1** | 163 | 4.7 | 6.8e-13 | 368 | 2.8 | 1.5e-05 |
| **ENSG00000289065.1** | 165 | 3.4 | 6.9e-13 | 111 | 2.2 | 1.1e-07 |
| **ENSG00000289626.1** | 169 | 2.6 | 7.8e-13 | 478 | 1.5 | 4.0e-05 |
| **THOC1-DT** | 174 | 3.2 | 9.9e-13 | 270 | 2.4 | 4.8e-06 |
| **ENSG00000288872.1** | 175 | 4.9 | 1.0e-12 | 190 | 3.3 | 1.1e-06 |
| **ENSG00000268403.2** | 178 | 3.4 | 1.2e-12 | 132 | 2.3 | 2.6e-07 |
| **ENSG00000289506.1** | 184 | 3.9 | 1.6e-12 | 88 | 3.1 | 6.7e-08 |
| **ENSG00000287821.1** | 185 | 4.6 | 1.6e-12 | 241 | 2.9 | 2.8e-06 |
| **ENSG00000272719.1** | 192 | 4.1 | 2.3e-12 | 187 | 3.3 | 1.0e-06 |
| **ENSG00000284602.1** | 195 | 2.4 | 2.4e-12 | 39 | 2.0 | 1.1e-08 |
| **ENSG00000240790.2** | 197 | 4.4 | 2.6e-12 | 697 | 2.5 | 2.1e-04 |
| **ENSG00000259135.1** | 200 | 4.2 | 2.7e-12 | 14,630 | 0.7 | 7.3e-01 |
| **ENSG00000289142.1** | 203 | 4.1 | 3.2e-12 | 257 | 2.5 | 4.1e-06 |
| **SMG7-AS1** | 215 | 2.9 | 4.4e-12 | 284 | 2.4 | 5.2e-06 |
| **ENSG00000286408.1** | 225 | 3.3 | 5.5e-12 | 155 | 2.2 | 4.3e-07 |
| **MIA2-AS1** | 229 | 2.6 | 5.7e-12 | 807 | 1.6 | 3.9e-04 |
| **ENSG00000289637.1** | 234 | 4.2 | 6.2e-12 | 1,085 | 2.1 | 2.1e-03 |
| **ENSG00000288804.1** | 241 | 4.5 | 7.4e-12 | 38 | 3.4 | 1.1e-08 |
| **ENSG00000289301.1** | 245 | 4.4 | 8.4e-12 | 620 | 2.8 | 1.2e-04 |
| **ENSG00000289317.1** | 248 | 4.4 | 8.7e-12 | 701 | 2.8 | 2.1e-04 |
| **ENSG00000273064.1** | 255 | 2.5 | 1.0e-11 | 86 | 2.3 | 6.2e-08 |
| **ENSG00000264666.2** | 258 | 2.8 | 1.1e-11 | 145 | 2.1 | 3.3e-07 |
| **ENSG00000286577.1** | 259 | 4.0 | 1.1e-11 | 539 | 2.5 | 7.3e-05 |
| **ENSG00000289296.1** | 268 | 3.9 | 1.4e-11 | 139 | 2.9 | 3.1e-07 |
| **ENSG00000255647.3** | 270 | 4.0 | 1.4e-11 | 106 | 3.3 | 9.8e-08 |
| **ENSG00000288988.1** | 272 | 3.2 | 1.6e-11 | 188 | 2.2 | 1.0e-06 |
| **ENSG00000288996.1** | 275 | 4.2 | 1.6e-11 | 865 | 2.3 | 6.9e-04 |
| **ENSG00000277182.1** | 276 | 2.0 | 1.7e-11 | 127 | 1.7 | 2.2e-07 |
| **ENSG00000288866.1** | 282 | 4.0 | 1.8e-11 | 240 | 3.1 | 2.8e-06 |
| **ENSG00000278743.1** | 289 | 3.4 | 2.2e-11 | 580 | 2.3 | 9.4e-05 |
| **MHENCR** | 297 | 1.3 | 2.8e-11 | 460 | 0.9 | 3.2e-05 |
| **ENSG00000283341.3** | 299 | 1.9 | 2.9e-11 | 696 | 1.0 | 2.1e-04 |
| **ENSG00000288061.2** | 302 | 2.2 | 3.2e-11 | 338 | 1.7 | 1.1e-05 |
| **ENSG00000289303.1** | 304 | 3.4 | 3.5e-11 | 428 | 2.5 | 2.2e-05 |
| **ENSG00000289499.1** | 321 | 4.3 | 5.1e-11 | 136 | 3.1 | 2.9e-07 |
| **ENSG00000288927.1** | 328 | 3.5 | 5.3e-11 | 198 | 3.4 | 1.3e-06 |
| **ENSG00000286482.1** | 329 | 2.0 | 5.3e-11 | 405 | 2.0 | 1.9e-05 |
| **ENSG00000289518.1** | 332 | 3.0 | 5.7e-11 | 530 | 2.3 | 6.8e-05 |
| **ZNF252P-AS1** | 333 | 2.0 | 5.9e-11 | 772 | 1.7 | 3.3e-04 |
| **ENSG00000272953.1** | 334 | 2.9 | 6.3e-11 | 72 | 2.5 | 4.0e-08 |
| **ENSG00000288896.1** | 338 | 3.6 | 6.8e-11 | 158 | 2.8 | 4.6e-07 |
| **ZFX-AS1** | 339 | 4.1 | 6.8e-11 | 928 | 2.1 | 9.1e-04 |
| **ENSG00000261519.3** | 341 | 3.2 | 7.5e-11 | 1,721 | 1.5 | 2.7e-02 |
| **ENSG00000261242.1** | 343 | 3.0 | 7.7e-11 | 688 | 2.1 | 2.0e-04 |
| **ENSG00000276524.1** | 346 | 3.4 | 8.3e-11 | 383 | 2.6 | 1.7e-05 |
| **ENSG00000278932.5** | 348 | 1.9 | 8.8e-11 | 45 | 1.3 | 1.5e-08 |
| **ENSG00000267882.2** | 355 | 4.0 | 1.0e-10 | 162 | 3.0 | 5.4e-07 |
| **MSRA-DT** | 365 | 3.3 | 1.2e-10 | 246 | 3.2 | 3.1e-06 |
| **ENSG00000289031.1** | 379 | 3.4 | 1.5e-10 | 442 | 2.8 | 2.5e-05 |
| **ENSG00000269399.2** | 380 | 2.0 | 1.5e-10 | 365 | 1.3 | 1.4e-05 |
| **ARHGEF2-AS2** | 383 | 2.2 | 1.6e-10 | 134 | 1.6 | 2.8e-07 |

| | PM/Scl-specific genes at 72h | | | PM/Scl-specific genes at 24h | | |
|---|---|---|---|---|---|---|
| Gene | Pos | logFC | adj.P.Val | Pos | logFC | adj.P.Val |
| **ENSG00000289222.1** | 389 | 3.3 | 1.8e-10 | 160 | 2.9 | 4.7e-07 |
| **DDX39B-AS1** | 399 | 3.8 | 2.3e-10 | 204 | 3.0 | 1.4e-06 |
| **CAGE1** | 406 | 3.2 | 2.6e-10 | 1,868 | 1.5 | 4.0e-02 |
| **ENSG00000289154.1** | 408 | 2.0 | 2.6e-10 | 350 | 1.4 | 1.2e-05 |
| **TERC** | 411 | 4.8 | 2.6e-10 | 977 | 2.1 | 1.2e-03 |
| **CAPN10-DT** | 422 | 1.8 | 3.2e-10 | 231 | 1.2 | 2.4e-06 |
| **LINC02776** | 423 | 3.5 | 3.2e-10 | 958 | 2.2 | 1.1e-03 |
| **HIGD2B** | 427 | 3.8 | 3.2e-10 | 652 | 2.6 | 1.6e-04 |
| **ENSG00000289159.1** | 435 | 3.9 | 3.8e-10 | 1,056 | 2.2 | 1.8e-03 |
| **ENSG00000264739.1** | 438 | 3.7 | 3.9e-10 | 193 | 3.0 | 1.1e-06 |
| **ENSG00000267212.1** | 447 | 2.9 | 4.3e-10 | 717 | 2.0 | 2.5e-04 |
| **ENSG00000273141.1** | 458 | 3.2 | 5.6e-10 | 344 | 2.3 | 1.1e-05 |
| **ENSG00000279198.1** | 461 | 2.1 | 5.8e-10 | 489 | 1.4 | 4.7e-05 |
| **ENSG00000287070.1** | 472 | 3.7 | 7.6e-10 | 555 | 2.1 | 8.5e-05 |
| **ENSG00000289067.1** | 473 | 2.3 | 7.6e-10 | 684 | 1.8 | 2.0e-04 |
| **LINC01126** | 478 | 2.2 | 8.2e-10 | 985 | 1.5 | 1.3e-03 |
| **LINC01089** | 484 | 1.2 | 9.0e-10 | 901 | 0.8 | 8.3e-04 |
| **UAP1-DT** | 487 | 4.7 | 9.5e-10 | 397 | 3.3 | 1.8e-05 |
| **ENSG00000288919.1** | 491 | 2.6 | 1.0e-09 | 1,461 | 1.1 | 1.3e-02 |
| **ENSG00000272906.1** | 497 | 2.6 | 1.1e-09 | 564 | 1.5 | 9.1e-05 |
| **ENSG00000287654.1** | 498 | 3.3 | 1.1e-09 | 543 | 2.5 | 7.5e-05 |
| **ENSG00000289334.1** | 500 | 2.4 | 1.1e-09 | 9 | 2.5 | 3.6e-10 |
| **ENSG00000276744.1** | 509 | 3.1 | 1.2e-09 | 181 | 2.9 | 8.5e-07 |
| **ENSG00000289253.1** | 515 | 2.7 | 1.3e-09 | 239 | 2.6 | 2.7e-06 |
| **ENSG00000279529.1** | 517 | 1.1 | 1.3e-09 | 1,305 | 0.8 | 6.7e-03 |
| **ENSG00000288737.1** | 532 | 3.4 | 1.5e-09 | 1,153 | 2.2 | 3.0e-03 |
| **ENSG00000255089.1** | 542 | 2.9 | 1.8e-09 | 600 | 2.7 | 1.1e-04 |
| **C10ORF95** | 551 | 3.1 | 2.0e-09 | 1,678 | 1.2 | 2.5e-02 |
| **ENSG00000274251.1** | 552 | 3.4 | 2.1e-09 | 310 | 2.7 | 6.5e-06 |
| **ENSG00000270012.1** | 560 | 1.5 | 2.4e-09 | 558 | 0.9 | 8.7e-05 |
| **ENSG00000274213.1** | 615 | 2.6 | 6.2e-09 | 1,908 | 1.2 | 4.3e-02 |
| **MIR5188** | 618 | 4.1 | 6.2e-09 | 484 | 3.1 | 4.4e-05 |
| **ENSG00000288744.1** | 630 | 3.2 | 7.5e-09 | 818 | 1.9 | 4.3e-04 |
| **ENSG00000272768.1** | 663 | 1.3 | 1.1e-08 | 585 | 1.0 | 9.7e-05 |
| **ENSG00000289182.1** | 664 | 1.7 | 1.1e-08 | 1,386 | 1.1 | 9.3e-03 |
| **ENSG00000288939.1** | 677 | 2.7 | 1.4e-08 | 646 | 2.1 | 1.5e-04 |
| **ENSG00000260651.1** | 683 | 3.5 | 1.4e-08 | 1,304 | 2.1 | 6.7e-03 |
| **ENSG00000272948.2** | 693 | 2.3 | 1.5e-08 | 194 | 2.1 | 1.2e-06 |
| **ENSG00000279259.1** | 698 | 2.1 | 1.6e-08 | 952 | 1.3 | 1.1e-03 |
| **OTULIN-DT** | 723 | 1.8 | 2.5e-08 | 619 | 1.7 | 1.2e-04 |
| **LINC00677** | 726 | 3.1 | 2.7e-08 | 1,273 | 1.7 | 5.9e-03 |
| **MIR23AHG** | 730 | 1.7 | 2.8e-08 | 517 | 1.1 | 6.1e-05 |
| **TIMMDC1-DT** | 733 | 3.1 | 2.9e-08 | 298 | 2.7 | 5.9e-06 |
| **ENSG00000275709.1** | 736 | 3.0 | 3.2e-08 | 447 | 2.4 | 2.8e-05 |
| **ENSG00000268670.1** | 737 | 2.0 | 3.2e-08 | 1,448 | 0.9 | 1.2e-02 |
| **ENSG00000289379.1** | 762 | 3.0 | 4.3e-08 | 242 | 2.5 | 2.9e-06 |
| **ENSG00000288746.1** | 779 | 2.3 | 5.6e-08 | 147 | 1.9 | 3.7e-07 |
| **ENSG00000289551.1** | 783 | 3.1 | 5.8e-08 | 250 | 2.9 | 3.5e-06 |
| **RENO1** | 791 | 1.1 | 6.4e-08 | 403 | 1.1 | 1.9e-05 |
| **ENSG00000289202.1** | 797 | 3.7 | 7.0e-08 | 292 | 3.1 | 5.5e-06 |
| **ENSG00000270019.1** | 809 | 2.4 | 8.1e-08 | 68 | 2.6 | 3.5e-08 |
| **SDR42E2** | 873 | 2.6 | 2.0e-07 | 657 | 2.1 | 1.6e-04 |
| **RN7SL521P** | 877 | 2.3 | 2.0e-07 | 945 | 1.7 | 1.1e-03 |
| **ENSG00000273289.1** | 914 | 2.6 | 2.8e-07 | 236 | 2.2 | 2.6e-06 |
| **ENSG00000289177.1** | 918 | 2.0 | 2.9e-07 | 1,019 | 1.2 | 1.5e-03 |
| **ENSG00000272969.1** | 927 | 2.9 | 3.3e-07 | 824 | 2.4 | 4.6e-04 |
| **CPEB2-DT** | 934 | 3.0 | 3.5e-07 | 1,066 | 2.3 | 1.9e-03 |
| **ENSG00000273363.1** | 1,013 | 2.1 | 7.5e-07 | 777 | 2.1 | 3.4e-04 |
| **LINC00115** | 1,015 | 1.6 | 7.6e-07 | 380 | 1.4 | 1.7e-05 |
| **ENSG00000288772.1** | 1,020 | 2.1 | 8.2e-07 | 574 | 1.9 | 9.3e-05 |
| **ENSG00000280152.1** | 1,037 | 1.6 | 1.0e-06 | 589 | 1.0 | 9.9e-05 |
| **ENSG00000278546.1** | 1,046 | 1.8 | 1.1e-06 | 1,104 | 1.4 | 2.3e-03 |
| **ENSG00000288813.1** | 1,121 | 2.3 | 2.3e-06 | 1,285 | 1.8 | 6.3e-03 |
| **ENSG00000277383.1** | 1,126 | 2.2 | 2.4e-06 | 562 | 1.7 | 9.0e-05 |
| **ENSG00000279140.1** | 1,183 | 2.1 | 4.1e-06 | 1,978 | 1.1 | 5.1e-02 |

| | PM/Scl-specific genes at 72h | | | PM/Scl-specific genes at 24h | | |
|---|---|---|---|---|---|---|
| **Gene** | **Pos** | **logFC** | **adj.P.Val** | **Pos** | **logFC** | **adj.P.Val** |
| **ENSG00000283959.2** | 1,215 | 1.4 | 5.5e-06 | 359 | 1.4 | 1.4e-05 |
| **ENSG00000288879.1** | 1,226 | 3.8 | 6.2e-06 | 1,064 | 2.8 | 1.9e-03 |
| **CPNE2-DT** | 1,228 | 2.0 | 6.3e-06 | 1,003 | 1.5 | 1.4e-03 |
| **ENSG00000279691.1** | 1,243 | 2.7 | 6.8e-06 | 1,120 | 2.2 | 2.5e-03 |
| **ENSG00000289229.1** | 1,259 | 2.6 | 7.5e-06 | 690 | 2.4 | 2.0e-04 |
| **ENSG00000273335.1** | 1,271 | 2.1 | 8.6e-06 | 1,046 | 1.5 | 1.7e-03 |
| **MIR3188** | 1,298 | 2.8 | 1.1e-05 | 2,197 | 1.5 | 7.3e-02 |
| **ANKH-DT** | 1,365 | 2.3 | 1.8e-05 | 2,750 | 1.5 | 1.3e-01 |
| **ENSG00000289152.1** | 1,417 | 2.5 | 3.0e-05 | 2,711 | 1.4 | 1.3e-01 |
| **ENSG00000288963.1** | 1,427 | 1.8 | 3.2e-05 | 8,647 | 0.8 | 4.9e-01 |
| **ENSG00000274737.1** | 1,438 | 1.9 | 3.4e-05 | 1,291 | 1.1 | 6.4e-03 |
| **ENSG00000289005.1** | 1,456 | 2.5 | 3.9e-05 | 997 | 2.1 | 1.4e-03 |
| **DYNLL2-DT** | 1,510 | 2.7 | 6.2e-05 | 431 | 2.3 | 2.3e-05 |
| **ENSG00000289241.1** | 1,594 | 2.3 | 1.2e-04 | 664 | 2.3 | 1.7e-04 |
| **ENSG00000262412.1** | 1,880 | 2.1 | 8.1e-04 | 1,117 | 2.1 | 2.5e-03 |
| **ENSG00000286113.1** | 1,896 | 1.7 | 8.9e-04 | 5,335 | 0.8 | 3.3e-01 |
| **SMG1-DT** | 1,915 | 2.0 | 1.0e-03 | 1,626 | 1.3 | 2.1e-02 |
| **ENSG00000289478.1** | 2,008 | 1.9 | 1.5e-03 | 1,935 | 1.6 | 4.6e-02 |
| **ENSG00000282936.2** | 2,325 | 0.8 | 5.9e-03 | 10,801 | 0.2 | 5.9e-01 |
| **KLF2-DT** | 2,450 | 1.6 | 8.2e-03 | 9,096 | 0.7 | 5.1e-01 |
| **LINC00896** | 2,798 | 1.6 | 1.8e-02 | 6,855 | 0.9 | 4.1e-01 |
| **ENSG00000241666.2** | 2,813 | 1.5 | 1.9e-02 | 2,525 | 1.3 | 1.1e-01 |
| **ENSG00000289235.1** | 3,427 | 1.5 | 4.6e-02 | 1,208 | 1.9 | 4.3e-03 |
| **ENSG00000288842.1** | 3,515 | 1.0 | 5.1e-02 | 1,633 | 1.4 | 2.1e-02 |
| **ENSG00000257258.2** | 5,979 | 0.9 | 1.8e-01 | 815 | 1.6 | 4.3e-04 |
| **EGFL7** | 5,984 | -0.6 | 1.8e-01 | 12,197 | -0.3 | 6.4e-01 |
| **FSCN1** | 6,851 | -0.3 | 2.2e-01 | 7,539 | -0.2 | 4.4e-01 |
| **ENSG00000273248.1** | 7,276 | 1.0 | 2.4e-01 | 3,532 | 1.2 | 2.1e-01 |
| **MAP3K11** | 7,914 | -0.2 | 2.8e-01 | 11,507 | -0.1 | 6.2e-01 |
| **CETP** | 8,760 | 0.7 | 3.2e-01 | 6,756 | 0.6 | 4.0e-01 |
| **EXOC3L2** | 10,122 | -0.8 | 3.9e-01 | 13,950 | -0.7 | 7.1e-01 |
| **RABEP2** | 10,627 | -0.2 | 4.1e-01 | 11,798 | -0.1 | 6.3e-01 |
| **COL18A1** | 12,710 | -0.3 | 5.2e-01 | 21,116 | -0.1 | 9.1e-01 |
| **ENSG00000278158.1** | 12,937 | 0.6 | 5.3e-01 | 6,661 | 0.8 | 4.0e-01 |
| **TNFAIP8L1** | 13,679 | -0.1 | 5.7e-01 | 19,181 | -0.1 | 8.7e-01 |
| **ITPKB** | 14,815 | 0.2 | 6.3e-01 | 8,470 | 0.3 | 4.8e-01 |
| **ENSG00000276570.1** | 15,050 | 0.1 | 6.3e-01 | 4,558 | 0.3 | 2.8e-01 |
| **MIR1915HG** | 19,066 | -0.2 | 8.0e-01 | 16,649 | -0.1 | 8.0e-01 |
| **OIT3** | 19,075 | -0.2 | 8.0e-01 | 25,019 | -0.0 | 9.9e-01 |
| **TNFRSF4** | 21,602 | -0.1 | 8.9e-01 | 17,070 | -0.3 | 8.1e-01 |
| **ENSG00000284484.1** | 22,602 | -0.1 | 9.2e-01 | 23,718 | 0.1 | 9.6e-01 |
| **PRAMEF13** | 25,349 | 0.0 | 9.9e-01 | 15,696 | 0.2 | 7.6e-01 |
| **TRIM51BP** | 25,706 | -0.0 | 1.0e+00 | 2,296 | 0.9 | 8.4e-02 |
